# Five years of Hospital at Home adoption in Catalonia: impact and challenges

**DOI:** 10.1101/2023.01.25.23284997

**Authors:** Rubèn González-Colom, Gerard Carot-Sans, Emili Vela, Mireia Espallargues, Carme Hernández, Francesc Xavier Jiménez, David Nicolás, Montserrat Suárez, Elvira Torné, Eulalia Villegas-Bruguera, Fernando Ozores, Isaac Cano, Jordi Piera-Jiménez, Josep Roca, the JADECARE consortium

## Abstract

**Background:** Hospital at home (HaH), either admission avoidance (AA) or early supported discharge (ESD), was increasingly implemented in Catalonia (7.7 M, Spain) for selected patients, achieving regional adoption within the 2011-2015 Health Plan. This study aimed to assess population-wide HaH outcomes over five years (2015-2019) in a consolidated regional HaH program and provide context-independent recommendations for service quality assurance.

**Methods:** A mixed-methods approach was adopted, combining population-based retrospective analyses of registry information with qualitative research. AA and ESD were separately compared with conventional hospitalization groups using propensity score matching techniques. In the analysis, we evaluated the 12-month period before the acute episode, the admission, and use of healthcare resources at 30 and 90 days after discharge. A panel of experts discussed the results and provided recommendations for monitoring HaH services.

**Results:** The adoption of AA steadily increased from 5,185 to 8,086 episodes/year (total episodes 31,901; mean age 73 (SD 17) years; 79% high-risk patients), whereas ESD remained stable over the study period, averaging 5,329 episodes per year (total episodes 26,646; mean age 68 (SD 16) years; 71% high-risk patients). Mortality rates were similar in HaH and conventional hospitalization within the episode (AA: 0.31% vs. 0.45%; ESD: 0.18% vs. 0.45%) and at 30-days (AA: 3.94% vs. 3.24%; ESD: 4.50% vs. 4.07%). Likewise, the frequency of patients requiring hospital re-admissions or ER visits 30 days after discharge was similar in HaH (AA and ESD) and the corresponding controls. The 27 healthcare providers assessed showed high variability in patients’ age, multimorbidity, severity of episodes, recurrences, and length of stay of AA episodes. Recommendations aiming at enhancing service delivery were produced.

**Conclusions:** Besides confirming safety and value generation of AA, we found that this service is delivered in a case-mix of diferent scenarios, encouraging provider-profiled monitoring of the service, particularly for ESD modalities.

**Impact statement:** We certify that this work is **confirmatory** of Admission Avoidance (AA) as a value-based service by analyzing, with a population-based approach, a five-year period after regional adoption of AA in Catalonia. The research indicates the need for implementing quality assurance programs after service adoption and provides clear insights on how shape quality monitoring. The current study outcomes add novel knowledge to previous reports in the field, such as:

1. Leff B, DeCherrie L v., Montalto M, Levine DM. A research agenda for hospital at home. *J Am Geriatr Soc*. 2022;70(4):1060-1069. doi:10.1111/JGS.17715
2. Levine DM, Ouchi K, Blanchfield B, et al. Hospital-Level Care at Home for Acutely Ill Adults: A Randomized Controlled Trial. *Ann Intern Med*. 2020;172(2):77-85. doi:10.7326/M19-0600
3. Montalto M, McElduff P, Hardy K. Home ward-bound: features of hospital in the home use by major Australian hospitals, 2011-2017. *Med J Aust*. 2020;213(1):22-27. doi:10.5694/mja2.50599
4. Hecimovic A, Matijasevic V, Frost SA. Characteristics and outcomes of patients receiving Hospital at Home Services in the South-West of Sydney. *BMC Health Services Research*. 2020;20(1):1090. doi:10.1186/s12913-020-05941-9
5. LEONG MQ ET AL. Comparison of Hospital-at-Home models: a systematic review of reviews. *BMJ Open*. 2021;11:43285. doi:10.1136/bmjopen-2020-043285

The current manuscript covers relevant knowledge gaps well-identified in the nine dimensions for future research in the field of hospital at home reported by Leff B et al, 2022. Moreover, the population-based approach of the research provides a valuable approach for quality assurance of the different service modalities.

Key Points

- Large scale adoption of Admission Avoidance shows value generation in real-world settings
- Implementation of continuous quality assurance monitoring after service adoption is highly recommended.

Why does this paper matter?
The population-based approach of the study design allows identification of key elements for service improvement after consolidated regional adoption of Hospital at Home
Key strengths of the research are: i) demonstration of healthcare value generation of AA in large scale adoption of the service; and ii) generation of insightful recommendations for enhanced service delivery and continuous quality monitoring.

## INTRODUCTION

Two decades after the first report assessing hospital at home (HaH) services,^1^ this type of care has raised increasing interest as a consolidated alternative to inpatient care for selected groups.^2–4^ HaH, delivered in its two modalities of admission avoidance (AA) and early supported discharge (ESD), has been associated with several advantages, including patient safety, reduction of nosocomial complications, similar or even better health outcomes compared to conventional hospitalization, high satisfaction levels from both patients and caregivers, and cost savings. In addition, by releasing physical beds, HaH contributes to building capacity for highly specialized care inpatient hospitalization. Moreover, in an integrated care scenario, HaH may become a relevant driver of vertical integration between hospital care and community-based health and social services by enhancing the care continuum.

However, due to poor comparability among reported experiences, HaH is often considered a case mix of service profiles.^5^ The findings reported in the literature raise several controversies in different areas, comprising the results of HaH in specific patient groups, the most appropriate implementation strategies for the service, and the quality-of-care delivery after service adoption.^4^ The lack of consensus precludes standardization and continuous quality assurance of the service in real-world settings.^4^ Therefore, understanding the heterogeneities behind the HaH has become crucial to define key performance indicators (KPIs) that can be used to ensure quality and sustainability over time and adjust the country-specific regulations of the service.

In Catalonia, a 7.7 million area in North-East Spain with a single public payer (Catalan Health Service),^6,7^ AA and ESD were successfully deployed during the 2011-2015 regional Health Plan.^6,8–10^ The HaH outcomes from that period were used to establish a specific reimbursement scheme based on all patient-refined diagnosis-related groups (APR-DRG),^11,12^ aiming at consolidating large-scale adoption of AA and ESD by healthcare providers across the region. In 2020, the Catalan Health Service, with the contribution of selected HaH professionals, issued a consensus document to standardize the service^13^ **(Supplementary Material S2)**.

Taking the consensus document as starting point, the current study sought to cover three main objectives: i) assess the adoption of HaH at the regional level during 2015-2019, ii) compare AA and ESD with corresponding matched control groups of inpatient hospitalizations, and iii) characterize the value of HaH within the healthcare system and generate recommendations for long-term continuous monitoring of service quality assurance. The time frame chosen for the analysis corresponds to the achievement of sustainable regional adoption of HaH, avoiding the presumptive impact of the SARS-CoV-2 pandemic on the service.

To this end, we designed a mixed-methods study that includes a quantitative retrospective assessment of patient HaH characteristics and outcomes. The information obtained from the analysis period constituted the basis for a co-creation process conducted with a local group of experts in HaH, aiming to draw conclusions and recommendations for enhanced HaH delivery.

## METHODS

### Overview of study design

The current study combined quantitative and qualitative research methodologies. The quantitative study was a retrospective observational analysis of the characteristics of AA and ESD services recipients and health results between January 1, 2015, and December 31, 2019. For the qualitative assessment, we conducted focus groups and surveys^14^ with a panel of experts in HaH to interpret the results of the quantitative analysis and generate recommendations for continuous quality assurance.

The Ethical Committee for Human Research at Hospital Clinic de Barcelona approved the study on September 8, 2021 (HCB/2021/0768) in the context of the EU project “Joint Action on implementation of digitally enabled integrated person-centered care” (JADECARE).^15^ The team involved in the study accessed only registry data. All the data were handled in compliance with the General Data Protection Regulation 2016/679 on data protection and privacy for all individuals within the European Union. Although no interventions out of routine care were applied, the study was conducted according to relevant legal requirements (Biomedical Research Act 14/2007 of July 3).

The quantitative study was reported according to the STROBE^16^ guidelines for observational studies, and the qualitative analysis was reported according to the SRQR^17^ guidelines.

### Population and data sources

All data used in the quantitative analysis were retrieved from the Catalan Health Surveillance System (CHSS).^18^ Since 2011, the CHSS has collected detailed information on the utilization of healthcare resources by the entire population of Catalonia. The CHSS assembles information on the use of healthcare resources across healthcare tiers, drugs, and other billable healthcare costs, such as non-urgent medical transportation, outpatient rehabilitation, respiratory therapies, and dialysis. We screened the CHSS for all episodes of AA and ESD reported in Catalonia between January 1, 2015, and December 31, 2019.

The same database was used to create two control-matched groups of contemporary conventional hospitalizations (one to be used as a control for AA and the other for ESD). Control-matched groups were created using a 1-to-1 propensity score matching (PSM)^19,20^ and Genetic Matching^21^ techniques based on GENetic Optimization Using Derivatives (GENOUD)^22^ algorithm to check and improve covariate balance iteratively. To ensure the comparability of the matched episodes, we screened contemporary admissions within the same hospital with identical Medicare Diagnosis Related Group^12^ category. In addition, the patient’s baseline characteristics were characterized and matched using data on demographics (i.e., age and gender), utilization of healthcare resources during the previous year (i.e., hospital admissions, emergency room visits, number of pharmacological prescriptions and the total healthcare expenditure), clinical and social risk factors (i.e., the morbidity burden, using the Adjusted Morbidity Groups^23,24^ (AMG) score, and the presence of active diagnoses related with health-related social needs^25^).

The overall comparability of the matched groups was assessed using the Mahalanobis distance,^26^ and Rubin’s B and Rubin’s R metrics. Comparability after PSM was considered acceptable if Rubin’s B was less than 0.25 and Rubin’s R was between 0.5 and 2.^27^

### Variables and Outcomes

The baseline characteristics of study patients (i.e., before admission) included age, sex, morbidity burden measured using the AMG score, hospitalizations, emergency room admissions, and expenditure within the past year. Information regarding the HaH episode included the length of stay (LoS) and the complexity of hospitalization, measured using two case-mix tools: the Case Mix Index-APR-DRG v35 (CMI),^28^ broadly used for payment purposes, and the Queralt index,^29,30^ recently developed by the Catalan Institute of Health and showing higher performance for predicting general hospitalization endpoints.

Besides the baseline and episode characteristics, we gathered information regarding healthcare expenditure, hospitalizations, and visits to the emergency room within the 30- and 90-days period after discharge. Expenditure information was obtained from reimbursements by the Catalan Health Service,^31^ since no operational costs^32,33^ were available for the entire study group. AA delivery is reimbursed as a specific healthcare service, with case costs estimated based on the APR-DRG categories of the main diagnostic, whereas ESD is considered part of conventional hospitalizations in terms of reimbursement. Other relevant outcomes included mortality, during the hospitalization and 30 and 90 days after discharge.

### Statistical analysis

Before the analysis, we removed from the databases all the incomplete records, duplicate entries, and outliers with unrepresentative baseline characteristics or anomalous LoS with a Z-value greater than |3|.

Since *p-values* tend to drop in large population-based samples, yielding significant differences in most comparisons,^34^ we used effect size measures to compare the baseline characteristics of the matched AA and ESD with their respective control groups and establish the impact of the intervention. Cohen’s D test was used to determine the effect size in numerical variables; the magnitude of the difference was assessed according to the following ranges: weak (< 0.20), small (0.2 – 0.5), moderate (0.5 – 0.8), large (0.8 −1.3), and very large (>1.3). Cohen’s W test was used for categorical variables, with the following ranges used to assess the magnitude of difference: weak (< 0.10), small (0.1 – 0.3), moderate (0.3 – 0.5), and large (>50). We computed 1,000 bootstrap replicates in both scenarios to generate the 95% CI.

Categorical variables were summarized as absolute values and percentages, whereas continuous variables were described by the mean and the standard deviation or the median and the interquartile range as appropriate.

To analyze heterogeneity among healthcare providers regarding the patient profile, we described the age and the morbidity burden (measured using the AMG index) of AA patients in each center. We also assessed the inclusion bias of each center by measuring the difference in mean age and AMG index between AA and conventional hospitalizations admitted for the exact cause within the same provider. Heterogeneity was also assessed regarding the LoS, the complexity of the episode and the repetition rate among HaH patients. In addition to the descriptive analysis, we addressed heterogeneity by conducting an ancillary cluster analysis using the K-means^35^ algorithm, incorporating information on the category of the hospital based on the number of hospital beds and their role in their corresponding health district. The average silhouette^36^ method was used to determine the optimal number of clusters.

All the data analyses were performed using R,^37^ version 4.1.1 (R Foundation for Statistical Computing, Vienna, Austria).

### Qualitative assessment

The qualitative study, which followed a grounded theory approach, included two focus group sessions with HaH experts. The first session aimed at interpreting the results obtained in the quantitative analysis described above, whereas the second session sought conclusions on four key items: i) healthcare value generation of HaH;^38^ ii) current challenges on evaluation of case complexities and costs; iii) sources of heterogeneity among providers; and iv) recommendations for quality assurance of HaH delivery after service adoption. The second session was preceded by the administration of a questionnaire (Supplementary material S1) for assessing the consensus strength. In all debates, the 2020 consensus document aiming at regional standardization of HaH^13^ was adopted as a reference.

The panel of 7 experts included 1-to-2 representatives of the most relevant organizations in implementing or assessing HaH services in Catalonia: two members from the Catalan-Balearic Society of Hospital at Home,^39^ two staff members from the Catalan Health Service,^40^ one staff member from the Health Quality and Assessment Agency of Catalonia (AQuAS),^41^ and two HaH experts from the local JADECARE team. A qualitative research and service design specialist was recruited as a facilitator for planning and leading the expert panel discussions. An extended description of the methodological details is provided in the online Supplementary material S1.

## RESULTS

### Adoption and characteristics of hospital at home

The CHSS registry recorded 58,547 episodes of HaH among the 27 healthcare providers offering this service to their catchment populations: 31,901 were AA episodes and 26,646 ESD (**Figure 1**). Overall, the activity of AA steadily increased during the study period from 5,185 to 8,086 episodes per year. In contrast, ESD remained stable from 2015 to 2019, averaging 5,329 episodes yearly. The relative frequency of AA vs ESD episodes was 0.82 in the eighth high-technology hospitals. In contrast, this ratio increased to 1.51 in the twelve general hospitals and 2.00 in the seven community hospitals. **Supplemental Material S1 -Table 1S** depicts yearly AA and ESD activity for each individual provider.

**TABLE 1.** Patient’s clinical characteristics and outcomes of the intervention of all patients admitted in admission avoidance (AA) and early supported discharge (ESD) programs.

|  | All AA<br>n= 31,901 | All ESD<br>n= 26,646 |
| --- | --- | --- |
| **DEMOGRAPHICS & MORBIDITY-COMPLEXITY** |  |  |
| *Age, mean (sd) | 73.11 (16.73) | 68.09 (15.66) |
| *Gender; n (%) |  |  |
| Male | 15,214 (47.69) | 12,938 (48.56) |
| Female | 16,687 (52.31) | 13,708 (51.44) |
| *AMG, mean (sd) | 29.51 (16.48) | 26.89 (16.72) |
| AMG category, n (%) |  |  |
| *Very low risk < P<sub>50</sub>* | 266 (0.83) | 361 (1.35) |
| *Low risk [P<sub>50</sub> - P<sub>80</sub>)* | 1,769 (5.55) | 1,956 (7.34) |
| *Moderate risk [P<sub>80</sub>-P<sub>95</sub>)* | 4,723 (14.81) | 5,223 (19.6) |
| *High risk [P<sub>95</sub>-P<sub>99</sub>)* | 5,476 (17.17) | 5,023 (18.85) |
| *Very high risk ≥ P<sub>99</sub>* | 19,667 (61.64) | 14,083 (52.86) |
| *Patients with HRSN associated to housing and economic conditions, n (%) | 5,063 (15.86) | 2,160 (8.11) |
| *Patients with HRSN associated to family and social environment, n (%) | 9,903 (31.03) | 5,834 (21.89) |
| Patients receiving palliative care, n (%) | 1,437 (4.5) | 606 (2.27) |
| **USE OF RESOURCES 12 MONTHS BEFORE ADMISSION** |  |  |
| *Patients requiring hospital admissions, n (%) | 15,957 (50.16) | 12,907 (48.51) |
| *Patients requiring emergency room visits, n (%) | 25,812 (81.14) | 21,197 (79.67) |
| *Total Expenditure in €, median (P<sub>25</sub>-P<sub>75</sub>) | 4,153.4 (1,695.8 - 8424.6) | 3,876.9 (1,537.0 – 8,170.4) |
| **HOME HOSPITALIZATION EPISODE** |  |  |
| LoS, mean (sd) | 8.47 (6.34) | 10.28 (8.71) |
| Mortality, n (%) | 103 (0.32) | 39 (0.15) |
| Queralt Index, mean (sd) | 28.34 (16.24) | 29.11 (18.52) |
| Case Mix Index | 0.66 | 0.68 |
| **USE OF RESOURCES 30 DAYS AFTER DISCHARGE** |  |  |
| Mortality, n (%) | 1,383 (4.35) | 666 (2.5) |
| Patients requiring hospital admissions, n (%) | 3,609 (11.35) | 3,113 (11.7) |
| Patients requiring emergency room visits, n (%) | 6,136 (19.29) | 5,145 (19.34) |
| Total Expenditure in €, median (P<sub>25</sub>-P<sub>75</sub>) | 279.1 (119.3 - 758.7) | 328.2 (163.3 - 780.1) |
| **USE OF RESOURCES 90 DAYS AFTER DISCHARGE** |  |  |
| Mortality, n(%) | 2,952 (9.28) | 1,431 (5.38) |
| Patients requiring hospital admissions, n (%) | 7,337 (23.06) | 5,887 (22.13) |
| Patients requiring emergency room visits, n (%) | 11,792 (37.07) | 9,237 (34.72) |
| Total Expenditure in €, median (P<sub>25</sub>-P<sub>75</sub>) | 918.93 (364.1 – 2,719.7) | 984.6 (486.5 – 2,688.9) |

AMG stands for Adjusted Morbidity Groups, HRSN for health-related social needs and LoS for length of stay. \* Matching variables.

**FIGURE 1.**
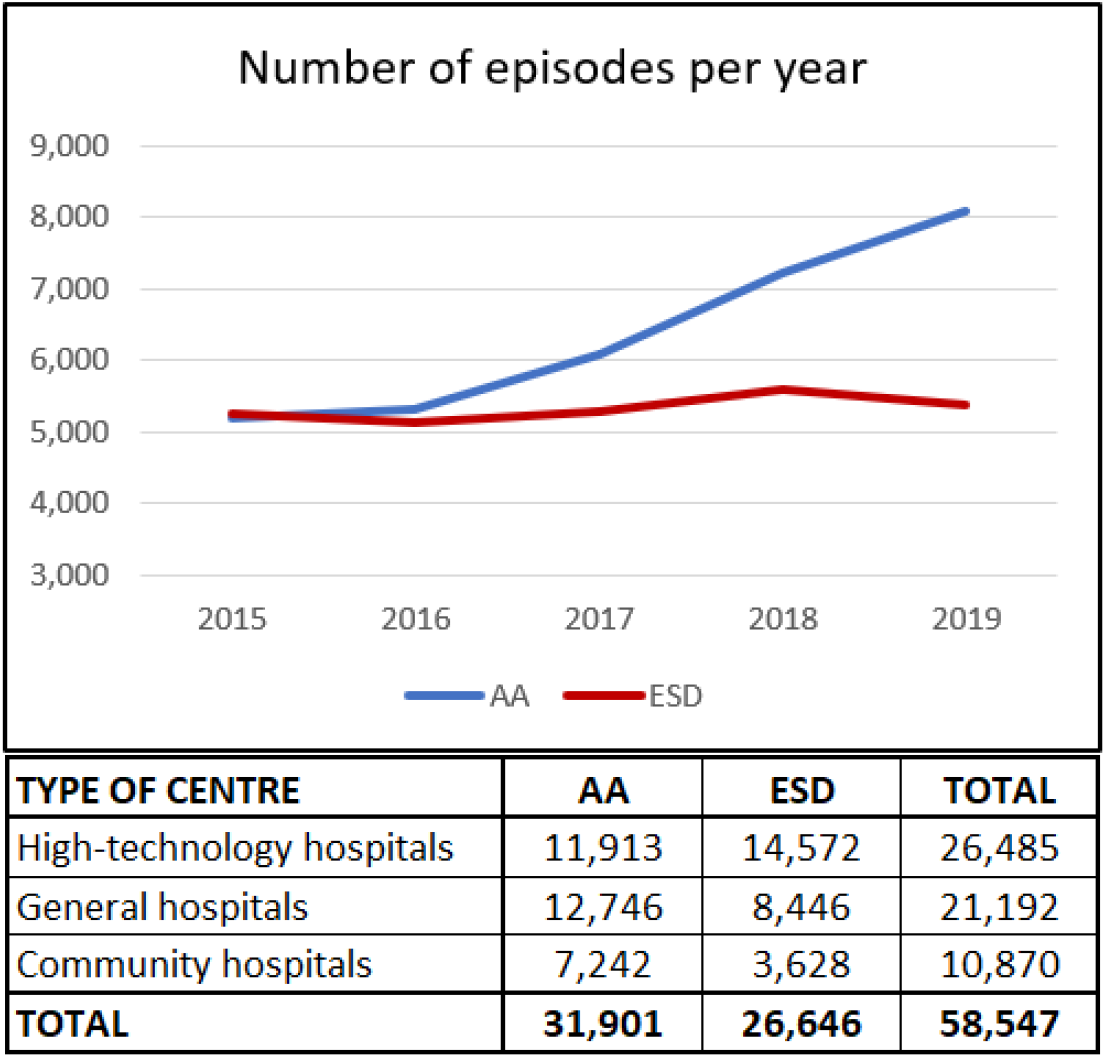
Number of admissions in of admission avoidance (AA) and early supported discharge (ESD) registered in 27 health providers from Catalonia between 2015 and 2019. Top panel – Total number of annual admissions in AA and ESD programs in Catalonia during the study period. Bottom panel – Total number of admissions in AA and ESD programs during the entire study period arranged by type of providers. For specific purposes of the study, healthcare providers were classified into three categories based on the number of hospital beds and their role in the corresponding health district: community hospitals (50–250 beds with internal medicine and general surgery), general or district hospitals (200–800 beds with several medical and surgical specialties), and high-technology hospitals (300–1,500 beds with roles of regional reference centers).

**Table 1** summarizes the main characteristics of the patients included in AA and ESD, distinguishing among four relevant timeframes, covering the patient’s baseline characteristics before the admission, the hospitalization episode, and the health outcomes assessed at 30- and 90-days post-discharge. On average, HaH patients were older, with a slightly higher prevalence of women in the two modalities of HaH. A substantial proportion of AA episodes corresponded to high-risk patients (i.e., with AMG score above the 95^th^ percentile of the AMG distribution for the entire population of Catalonia). In contrast, the number of episodes corresponding to high-risk patients was slightly lower in ESD. The two study groups showed a substantial prevalence of health-related social needs associated with housing and economic conditions. Overall, the two groups had high use of healthcare resources during the year before the acute episode.

The acute episode showed low mortality rates in both AA and ESD, and moderate levels of complexity for the two groups, measured using the Queralt index and CMI. The ten leading main diagnoses at discharge in AA (**Table 2**) were predominantly medical, whereas post-surgical procedures associated with locomotor problems were the leading ESD diagnoses.

**TABLE 2.**
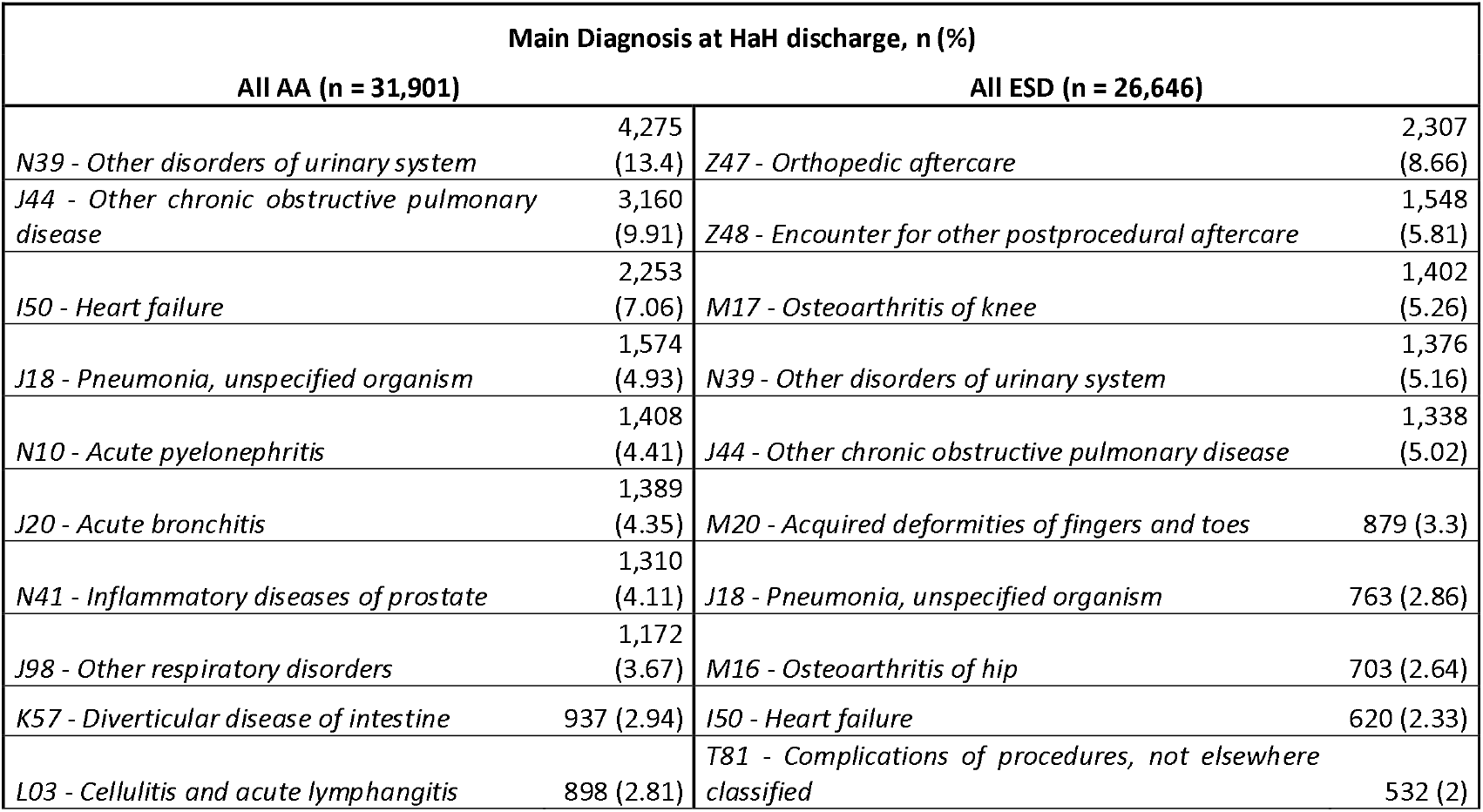
Top 10 of most prevalent main diagnosis at discharge in patients admitted in admission avoidance (AA) and early supported discharge (ESD) programs.

### Comparisons between hospital at home and conventional hospitalizations

**Table 3** compares the characteristics of AA and ESD with the corresponding matched control groups of patients under conventional hospitalization. Mortality during the acute episode was low and similar between intervention and controls. The effect size analyses indicated that the severity of the acute episodes, measured using the Queralt index and the CMI, was significantly higher in conventional hospitalizations than in the corresponding HaH intervention, either AA or ESD. Also, the LoS was significantly longer in AA and ESD than in conventional hospitalizations. Despite the statistical significance, the differences observed in all endpoints between HaH and conventional hospitalization were associated with a small effect size (i.e., the differences between groups were 0.2 to 0.5 times the SD).

**TABLE 3.**
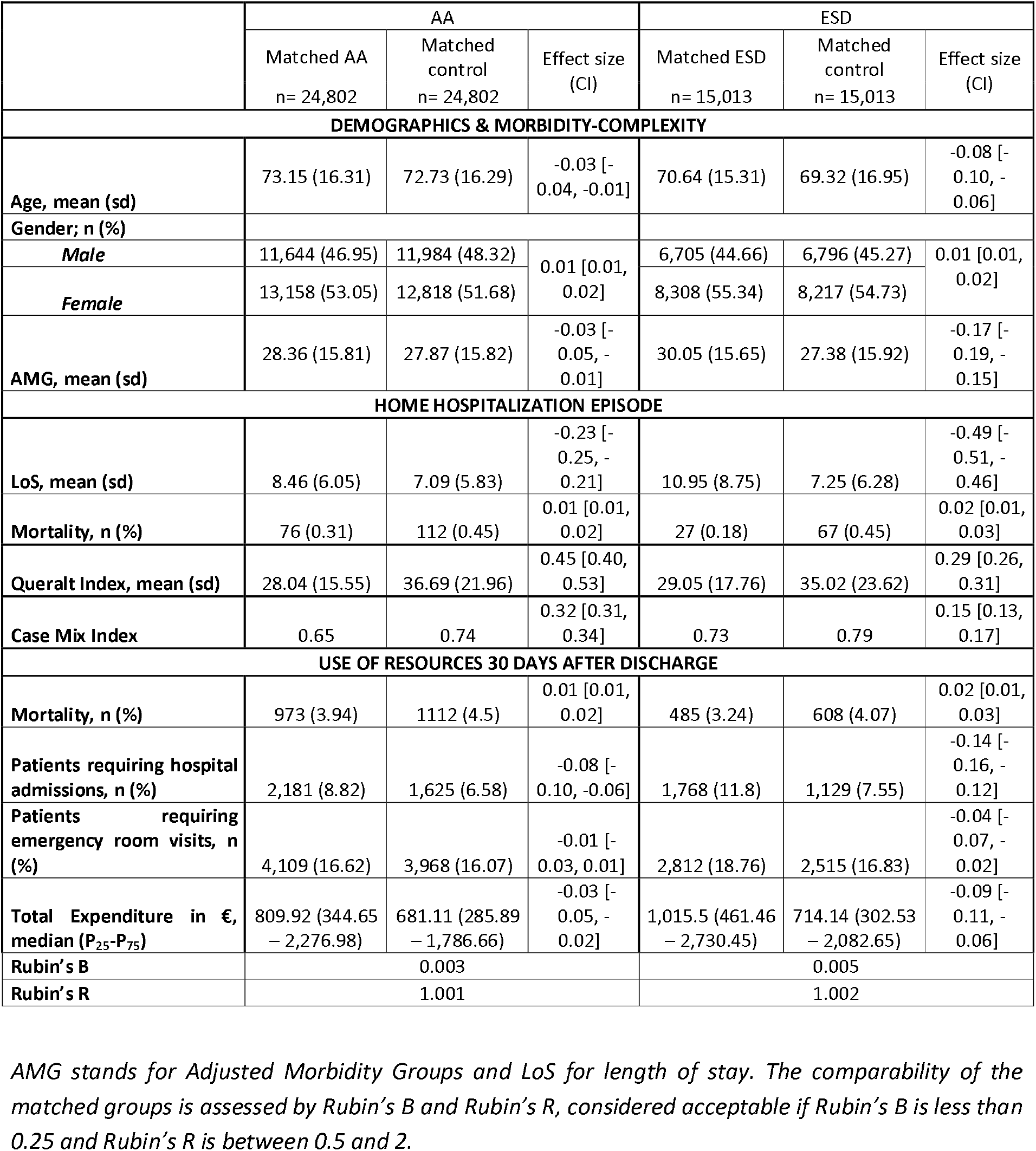
Comparison of patients’ clinical characteristics and the outcomes of the intervention between individuals admitted in Hospital at Home programs, admission avoidance (AA) and early supported discharge (ESD), and their corresponding matched controls of conventional hospitalizations arranged by clusters of providers.

During the 30 days after discharge, mortality rates were low, with no differences between the intervention (AA and ESD) and the corresponding control groups. Likewise, re-admissions, visits to the emergency room, and healthcare expenditure were also similar between HaH and controls.

### Heterogeneities among providers

Comparisons among the 27 healthcare providers showed huge heterogeneities in AA, in several dimensions, including age at admission (provider mean values ranging from 62.16 to 83.39 years), multimorbidity-complexity within the 12-month period before admission expressed by AMG scoring (from 19.47 to 38.79), and severity of the acute episode assessed either using the APR-DRG (from 0.54 to 0.87) or the Queralt index (from 13.34 to 42.31). Likewise, similar inter-provider variability was also observed in all other two variables analyzed: LoS (from 4.8 to 14.7 days) and percent of repeaters, indicating patients with more than one HaH episode during the study period (from 8.8% to 33.6%).

### Qualitative assessments and expert recommendations

The full set of results of the quantitative analyses (**Supplemental Material S1 -Figures 1S-8S** and **Tables 2S-4S**) were presented to the panel of experts for discussion and interpretation. Based on the unaligned delivery and reimbursement policy of ESD services, the panel of experts agreed that recommendations and conclusions regarding AA cannot be applied to ESD and that more exhaustive studies should be conducted to propose specific provider-profiled KPIs for ESD modalities.The consensus achieved by the experts’ group during the second session regarding the four key items is summarized below:

#### Health care value generation of HaH

The experts unanimously concluded that AA: i) is safe, ii) produces similar outcomes compared to conventional hospitalization with equal reimbursement regimens, iii) patients’ and careers’ perception of the service is positive, and iv) health professionals show high levels of engagement. Moreover, AA clearly contributes to building bed capacity and generates a scenario with a high potential for enhancing transitional care, as well as vertical integration.

#### Current challenges on evaluation of case complexities and costs

The experts fully agreed that indices currently used for measuring episode severity, such as APR-DRG, do not appropriately reflect care needs in most patients admitted to HaH.

The experts agreed that AA generates savings compared to conventional hospitalizations.^4,42^ Such cost reductions seem associated with fewer personnel and structure requirements. However, they unanimously encouraged further research on the analytical costs of HaH as a robust basis for future reimbursement policies.

#### Sources of heterogeneities among providers

While experienced hospitals with mature HaH teams are admitting older and complex patients to AA showing good outcomes, other providers display different profiles of AA admissions apparently modulated by one or several concomitant factors, including the following: i) provider-driven strategies focusing on subsets of patients with specific diagnoses; ii) local ecosystem characteristics with the availability of other integrated care services overlapping HaH; or iii) different levels of maturity of HaH teams. The experts agreed on the need for additional field studies to better characterize heterogeneities in AA delivery. These studies should have a two-fold aim: to assess alignment with regional standardization of the service^13^ and to explore the need for provider-specific KPIs.

#### Recommendations for enhanced service delivery

Specific expert recommendations are summarized in **Figure 2**, second and third columns. The experts prioritized the generation of a protocol for continuous quality assurance of AA ensuring its applicability, on routine basis, to all range of providers. A core component should be a dashboard, combining core KPI and provider profiled KPI, to be elaborated as a specific component of the existing Modules for Monitoring Quality Indicators.^43^ A detailed description of the experts’ opinions can be found in the online supplementary material.

**FIGURE 2.**
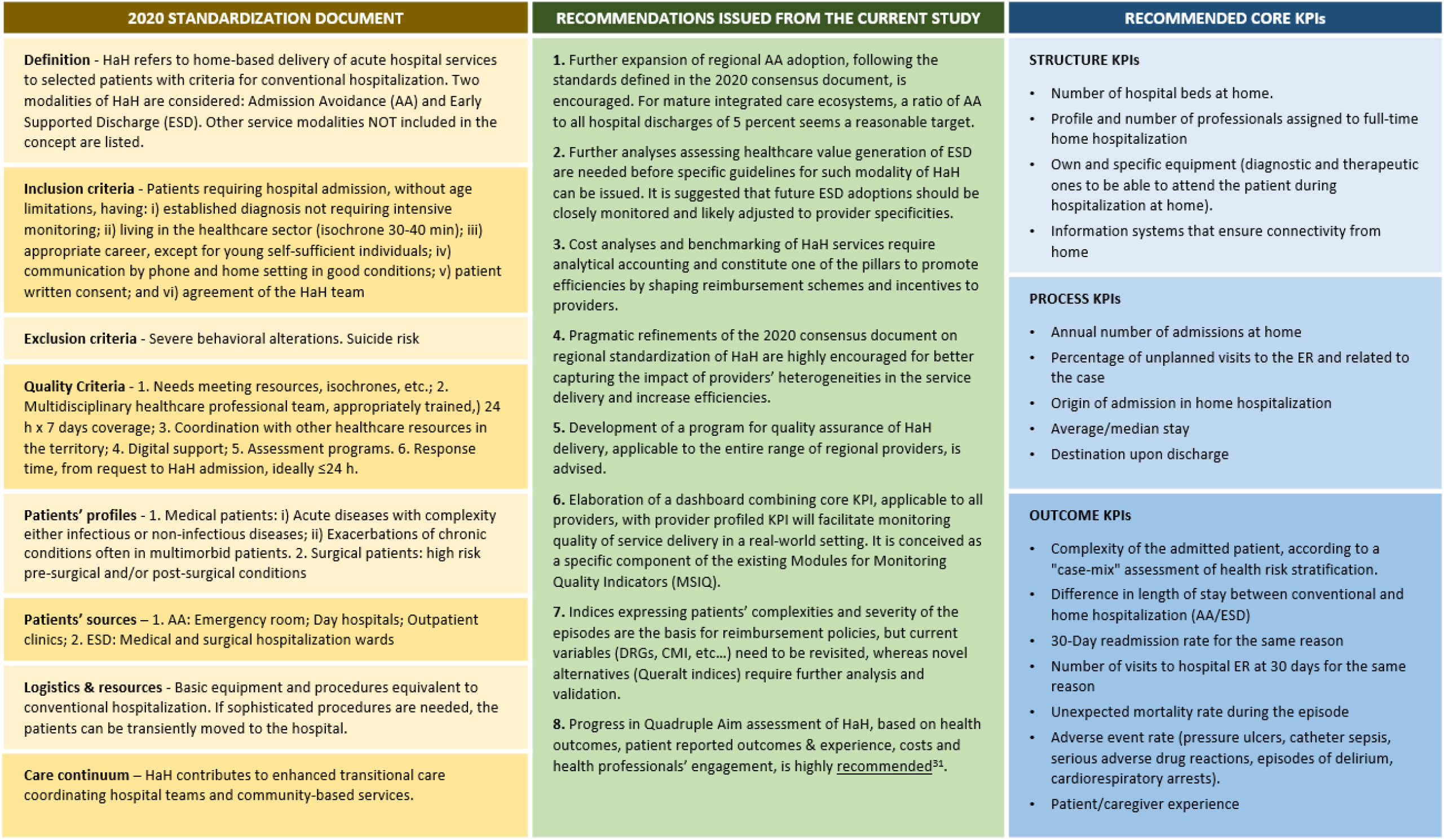
Hospital at Home (HaH): summary of the 2020 document on regional standardization^13^ and current study recommendations.

## DISCUSSION

The mixed-methods approach adopted in the current research contributed to enriching the interpretation of the retrospective quantitative assessment of consolidated HaH delivery in 27 different providers of the same healthcare system. The outcomes observed in AA were aligned with relevant reports^1–4,44–48^ fully supporting the healthcare value generation of this modality of HaH, as well as its potential for capacity building of hospital beds and contributions to the care continuum. Overall, the study outcomes encourage further expansion of the regional adoption of AA, following the recommendations generated by the group of experts. In contrast, the qualitative analysis of ESD results clearly indicated the need for a cautious approach to future developments of this modality of HaH.

Another important conclusion of our study was the need for continuous long-term monitoring of quality assurance after the successful adoption of the service. The report provides recommendations, to elaborate and implement such quality assurance programs adapting existing regional tools. Future refinement of such tools incorporating a Triple or Quadruple Aim approach was already analyzed and highly encouraged.^33^ In the current scenario, we showed that regional deployment of HaH in Catalonia was conducted with a well-structured program within the 2011-2015 Health Plan, and successful adoption of AA was achieved during the study period. However, the report identified a significant potential for improving AA delivery after adoption, as stressed in the recommendations.

A recently reported contemporary assessment of analytical costs in the Hospital Clinic of Barcelona, one of the leading public healthcare providers in the area, demonstrated that AA generates substantial savings, reducing operational costs by half, mainly due to personnel and structure costs.^32,33^ In this regard, it must be emphasized that expenses during the episode of AA, based on APR-DRGs, do not correspond with the operational costs of the intervention. The expert group agreed that the implementation of analytical accounting should be extended to all providers to build up adequate reimbursement strategies. This approach would contribute to enhancing investments in healthcare innovation that, in turn, generate efficiencies both at provider and health system levels. Analytical accounting would also provide a rationale for specific reimbursement plans favoring provider-profiled service delivery.

The observed heterogeneities are consistent with disparities found in the literature.^2–4^ However, the assessment of multiple providers within the same healthcare system allowed us to investigate these differences regardless of the healthcare structure, payment model, cultural constraints and/or type of professionals involved that may vary between countries and systems. The factors explaining these heterogeneities within the same healthcare system were extensively debated by the experts participating in the qualitative analysis. Recommendations for adequate management of the heterogeneities (Figure 2) can be summarized in two complementary lines of action to be supported by specific reimbursement incentives: i) align service delivery with the core components of the 2020 consensus statement,^13^ and ii) foster provider-profiled delivery of AA. All in all, the recommendations generated by the expert group should provide the essential elements for the future expansion of enhanced AA delivery at a regional level. Its application can also be useful in defining specificities of future ESD policies.

We acknowledge some intrinsic limitations of the current study. One of them is the exclusive use of registry data without information on details of complexities and clinical incidences during HaH episodes. Moreover, the lack of analytical costs was also an important constraint assessment of the potential of value generation of AA, as well as to explore the impact of reimbursement policies on providers’ heterogeneities. As described above, all economic calculations in the current study were based on expenditure data.^31^ However, we believe that the characteristics of the study design and the availability of clinical and analytical data from the area^32,33^ positively influenced the analyses carried out in the current research and facilitated recommendations for enhancing the quality of service delivery that can be generalized to other integrated care services.

## Conclusions

Besides confirming safety and value generation of AA, we found that this service is delivered in an heterogeneous case-mix of healthcare scenarios that may also result in heterogeneous outcomes. Therefore, aside from general key performance indicators, provided-profiled indicators should be established to monitor for continuous quality assurance of the service after adoption. The close monitoring of HaH is particularly relevant for ESD modalities, which seem to require a specific treatment. The recommendations from a panel of experts provided in this study can be used as basis for planning HaH monitoring in other countries.

## Data Availability

All data produced in the present study are available upon reasonable request to the authors

## ACRONYMS

AA: Admission Avoidance
AMG: Adjusted Morbidity Groups
APR-DRG: All-Patient Refined Diagnosis Related Groups
CHSS: Catalan Health Surveillance System
CMI: Case Mix Index
ESD: Early Supported Discharge
LoS: Length of Stay
HaH: Hospital at Home
KPI: Key Performance Indicator
PSM: Propensity Score Matching

## AUTHORS CONTRIBUTION

JR, IC and JPJ designed and directed the project. RGC led the execution of the quantitative analysis, processed the experimental data, performed the statistical analysis and created the figures. EV generated the study database and provided statistical support. GCS led the execution of the qualitative research, developed the study surveys, and summarized the conclusions of the focus groups. FO designed and moderated the focus groups. ME, CH, FXJ, DN, MS, ET, EVB composed the panel of experts consulted in the study. All authors discussed the results and contributed to the final manuscript.

## REFERENCES

1. Leff B et al. Hospital at home: Feasibility and outcomes of a program to provide hospital-level care at home for acutely ill older patients. Ann Intern Med. 2005;143(11). doi:10.7326/0003-4819-143-11-200512060-00008

2. Conley J et al. Alternative Strategies to Inpatient Hospitalization for Acute Medical Conditions: A Systematic Review. JAMA Intern Med. 2016;176(11):1693–1702. doi:10.1001/JAMAINTERNMED.2016.5974

3. Wong JB et al. Hospital care at home: Better, cheaper, faster? Ann Intern Med. 2020;172(2):145–146. doi:10.7326/M19-3714

4. Leong MQ et al. Comparison of Hospital-at-Home models: a systematic review of reviews. BMJ Open. 2021;11:43285. doi:10.1136/bmjopen-2020-043285

5. Pigott T et al. Identifying, documenting, and examining heterogeneity in systematic reviews of complex interventions. J Clin Epidemiol. 2013;66(11):1244–1250. doi:10.1016/J.JCLINEPI.2013.06.013

6. DEPARMENT OF HEALTH. Government of Catalonia Health Plan for 2011–2015.; 2012.

7. DEPARTMENT OF HEALTH. Goverment of Catalonia Health Plan for 2016–2020.; 2016.

8. Hernández C et al. Implementation of Home Hospitalization and Early Discharge as an Integrated Care Service: A Ten Years Pragmatic Assessment. Int J Integr Care. 2018;18(2):12. doi:10.5334/ijic.3431

9. Torre JA De La et al. Differences in Results and Related Factors Between Hospital-at-Home Modalities in Catalonia: A Cross-Sectional Study. J Clin Med. 2020;9(5). doi:10.3390/JCM9051461

10. AGENCIA DE QUALITAT I AVALUACIÓ SANITARIES DE CATALUNYA. Informes AQuAS: Hospitalización a Domicilio.; 2018.

11. Mihailovic N et al. Review of Diagnosis-Related Group-Based Financing of Hospital Care. Heal Serv Res Manag Epidemiol. 2016;3. doi:10.1177/2333392816647892

12. Goldfield N. The evolution of diagnosis-related groups (DRGs): From its beginnings in casemix and resource use theory, to its implementation for payment and now for its current utilization for quality within and outside the hospital. Qual Manag Health Care. 2010;19(1):3–16. doi:10.1097/QMH.0B013E3181CCBCC3

13. SERVEI CATALÀ DE LA SALUT (CATSALUT). Model Organitzatiu d’hospitalització a Domicili de Catalunya: Alternativa a l’hospitalització Convencional.; 2020.

14. Abookire S et al. Health Design Thinking: An Innovative Approach in Public Health to Defining Problems and Finding Solutions. Front Public Heal. 2020;8:459. doi:10.3389/FPUBH.2020.00459/FULL

15. JADECARE (2020-2023). Joint Action on implementation of digitally enabled integrated person-centred care. Published 2020. https://www.jadecare.eu/

16. Von Elm E et al. The Strengthening the Reporting of Observational Studies in Epidemiology (STROBE) statement: Guidelines for reporting observational studies. Ann Intern Med. 2007;147(8):573–577. doi:10.7326/0003-4819-147-8-200710160-00010

17. O’Brien BC et al. Standards for reporting qualitative research: A synthesis of recommendations. Acad Med. 2014;89(9):1245–1251. doi:10.1097/ACM.0000000000000388

18. Farré N et al. Medical resource use and expenditure in patients with chronic heart failure: a population-based analysis of 88 195 patients. Eur J Heart Fail. 2016;18(9):1132–1140. doi:10.1002/ejhf.549

19. Austin PC. An Introduction to Propensity Score Methods for Reducing the Effects of Confounding in Observational Studies. Multivariate Behav Res. 2011;46(3):399. doi:10.1080/00273171.2011.568786

20. A T et al. Should I stay or should I go? A retrospective propensity score-matched analysis using administrative data of hospital-at-home for older people in Scotland. BMJ Open. 2019;9(5). doi:10.1136/BMJOPEN-2018-023350

21. Diamond A et al. Genetic Matching for Estimating Causal Effects: A General Multivariate Matching Method for Achieving Balance in Observational Studies. Rev Econ Stat. 2013;95(3):932–945. doi:10.1162/REST_a_00318

22. Mebane WR et al. Genetic Optimization Using Derivatives: The rgenoud Package for R. JSS J Stat Softw. 2011;42.

23. Monterde D et al. Validity of adjusted morbidity groups with respect to clinical risk groups in the field of primary care. Aten Primaria. 2018;51(3):153–161. doi:10.1016/J.APRIM.2017.09.012

24. Vela E et al. Performance of quantitative measures of multimorbidity: a population-based retrospective analysis. BMC Public Heal 2021 211. 2021;21(1):1–9. doi:10.1186/S12889-021-11922-2

25. Bensken WP et al. ICD-10 Z-Code Health-Related Social Needs and Increased Healthcare Utilization. Am J Prev Med. 2022;62(4):e232–e241. doi:10.1016/J.AMEPRE.2021.10.004

26. De Maesschalck R et al. The Mahalanobis distance. Chemom Intell Lab Syst. 2000;50(1):1–18. doi:10.1016/S0169-7439(99)00047-7

27. Rubin DB. Using Propensity Scores to Help Design Observational Studies: Application to the Tobacco Litigation. Heal Serv Outcomes Res Methodol 2001 23. 2001;2(3):169–188. doi:10.1023/A:1020363010465

28. Fetter RB et al. Case mix definition by diagnosis-related groups. Med Care. 1980;18(Suppl 2):1–53.

29. Monterde D et al. Performance of Comprehensive Risk Adjustment for the Prediction of In-Hospital Events Using Administrative Healthcare Data: The Queralt Indices. Risk Manag Healthc Policy. 2020;13:271. doi:10.2147/RMHP.S228415

30. Monterde D et al. Performance of Three Measures of Comorbidity in Predicting Critical COVID-19: A Retrospective Analysis of 4607 Hospitalized Patients. Published online 2021. doi:10.2147/RMHP.S326132

31. Vela E et al. Análisis poblacional del gasto en servicios sanitarios en Cataluña (España): ¿qué y quién consume más recursos? Gac Sanit. 2019;33(1):24–31. doi:10.1016/J.GACETA.2017.05.017

32. Hernandez C et al. Value of Admission Avoidance: Cost-Consequence Analysis of One-Year Activity in a Consolidated Service. Appl Health Econ Health Policy.

33. Herranz C et al. Prospective cohort study for assessment of integrated care with a triple aim approach: hospital at home as use case. BMC Health Serv Res. 2022;22(1):1–12. doi:10.1186/S12913-022-08496-Z/FIGURES/3

34. Lin M et al. Too Big to Fail: Large Samples and the p-Value Problem. Inf Syst Res. 2013;24(4):906–917. doi:10.1287/ISRE.2013.0480

35. Macqueen J. Some Methods for Classification and Analysis of Multivariate Observations. Proc 5th Berkeley Symp Math Stat Probab. 1967;1(Statistics, University of California Press, Berkeley):281–297.

36. Kaufman L et al. Finding Groups in Data: An Introduction to Cluster Analysis. Biometrics. 1991;47(2):788. doi:10.2307/2532178

37. R CORE TEAM. R: A language and environment for statistical computing. Published online 2021.

38. Porter ME et al. Redefining Health Care: Creating Value-Based Competition on Results.; 2006.

39. SOCIETAT CATALANO-BALEAR D’HOSPITALITZACIÓ DOMICILIÀRIA. Accessed January 21, 2023. http://webs.academia.cat/societats/hospdomicil/index.php

40. CATALAN HEALTH SERVICE. Accessed January 21, 2023. https://catsalut.gencat.cat/ca/inici/

41. HEALTH QUALITY AND ASSESSMENT AGENCY OF CATALONIA (AQUAS). Accessed January 21, 2023. https://aquas.gencat.cat/ca/inici/index.html#googtrans(ca%7Cen)

42. Arsenault-Lapierre G et al. Hospital-at-Home Interventions vs In-Hospital Stay for Patients With Chronic Disease Who Present to the Emergency Department: A Systematic Review and Meta-analysis. JAMA Netw open. 2021;4(6). doi:10.1001/JAMANETWORKOPEN.2021.11568

43. MODULES FOR MONITORING QUALITY INDICATORS (MSIQ). Accessed January 21, 2023. https://transparencia.csi.cat/document/10140/detail/

44. Sullivan JL et al. Hospital In Home: Evaluating Need and Readiness for Implementation (HENRI) in the Department of Veterans Affairs: protocol for a mixed-methods evaluation and participatory implementation planning study. Implement Sci Commun 2022 31. 2022;3(1):1–10. doi:10.1186/S43058-022-00338-7

45. Montalto M et al. Home ward bound: features of hospital in the home use by major Australian hospitals, 2011-2017. Med J Aust. 2020;213(1):22–27. doi:10.5694/MJA2.50599

46. Hecimovic A et al. Characteristics and outcomes of patients receiving Hospital at Home Services in the South West of Sydney. BMC Health Serv Res. 2020;20(1). doi:10.1186/S12913-020-05941-9

47. Tibaldi V et al. Hospital at home for elderly patients with acute decompensation of chronic heart failure: a prospective randomized controlled trial. Arch Intern Med. 2009;169(17):1569–1575. doi:10.1001/ARCHINTERNMED.2009.267

48. Federman AD et al. Association of a Bundled Hospital-at-Home and 30-Day Postacute Transitional Care Program With Clinical Outcomes and Patient Experiences. JAMA Intern Med. 2018;178(8):1033–1041. doi:10.1001/JAMAINTERNMED.2018.2562

