## Supplementary Material S1 for "Five years of Hospital at Home adoption in Catalonia: impact and challenges"

Supplementary appendix

### CONTENT

1 Supplementary Methods ... 3

1.1 Supplementary Methods of the Quantitative study ... 3

1.1.1 Population and data sources ... 3

1.2 Supplementary Methods of the Qualitative study ... 4

1.2.1 Composition of the Panel of experts ... 4

1.2.2 Phases of the qualitative study ... 4

1.2.3 Characteristics of the First Focus Group ... 4

1.2.4 Pre-session survey: structured questionnaire ... 5

1.2.5 Characteristics of the second focus group ... 7

2 Supplementary Results ... 8

2.1 Supplementary Results of the Quantitative study ... 8

TABLE 1S – Number of admissions in of Admission Avoidance (AA) and Early Supported Discharge (ESD) registered in 27 health providers from Catalonia between 2015 and 2019. ... 9

TABLE 2S – Comparison of the patient's clinical characteristics at admission and the complexity of the intervention in Admission Avoidance (AA) programs among clusters of providers. ... 11

TABLE 3S – Patient's clinical characteristics and outcomes of the intervention of all the patients admitted in hospital admission avoidance programs arranged by clusters of providers. ... 12

TABLE 4S – Comparison of patients' clinical characteristics and the outcomes of the intervention between individuals admitted in hospital admission avoidance programs and their corresponding matched controls of conventional hospitalizations arranged by clusters of providers. ... 14

FIGURE 1S - Comparison of the patient's age among providers in hospital admission avoidance programs. ... 15

FIGURE 2S - Comparison of the patient's clinical complexity assessed with the Adjusted Morbidity Groups (AMG) score among providers in hospital admission avoidance programs. ... 16

FIGURE 3S - Comparison of the hospitalization complexity among providers assessed with the APR-DRG score in hospital admission avoidance programs. ... 17

### 1 SUPPLEMENTARY METHODS

#### 1.1 SUPPLEMENTARY METHODS OF THE QUANTITATIVE STUDY

##### 1.1.1 Population and data sources

All data used in the quantitative analysis of the hospital at home (HaH) programs, including admission avoidance (AA) and early supported discharge (ESD), were retrieved from the Catalan Health Surveillance System (CHSS). [1] We screened the CHSS for all episodes of AA and ESD reported in Catalonia between January 1, 2015, and December 31, 2019. To differentiate between the two modalities of HaH, we considered that a patient was admitted in an ESD program if (1) a conventional hospitalization with discharge date ±1 day apart from the HaH admission was registered, and (2) the duration of the previous episode of conventional hospitalization lasted at least two days, except for hospitalizations due to surgical intervention, which could be shorter. The remaining HaH episodes were considered AA.

The same database was used to create two control-matched groups of contemporary conventional hospitalizations (one to be used as a control for AA and the other for ESD). Control-matched groups were created using a 1-to-1 Propensity Score Matching (PSM)[2,3] and Genetic Matching[4] techniques based on GENetic Optimization Using Derivatives (GENOUD)[5] to iteratively check and improve covariate balance. Matching was conducted in two steps to ensure the comparability of AA and ESD episodes with their respective control groups.

In the first step, we selected up to ten conventional hospitalization candidates for each HaH episode, using a logistic regression model based on sex and age and meeting the following criteria: (1) contemporary episodes from the same hospital, (2) same Medicare Diagnosis-Related Group (DRG)[6] category, and (3) length of stay of at least one day. The use of DRG category as the sole complexity indicator prevented us from the overmatching that may occur with other indicators that depend on procedures or nosocomial complications.

In the second step, we conducted the 1-to-1 PSM (calliper: 0.05, function: logit, replace: FALSE, ratio: 1:1, matching method: Genetic Matching) for each healthcare provider, allowing only episodes with the same DRG code. The following variables were used to adjust the baseline characteristics of patients: age, gender, number of admissions within the previous year, number of visits to the emergency room within the previous year, number of drugs prescribed within the previous year, individual healthcare expenditure across the health system in the previous year, presence of an active diagnosis of health-related social needs,[7] and the morbidity burden according to the Adjusted Morbidity Groups (AMG)[8,9] index. The AMG is a population-based risk tool that summarizes comorbidity burden using a weighted sum of all chronic and relevant acute diagnostic codes and has shown to correlate with mortality and use of healthcare resources, among others[10].

#### 1.2 Supplementary Methods of the Qualitative study

##### 1.2.1 Composition of the Panel of experts

| | |
|---|---|
| **Elvira Torné** | Catalan Health Service[11] |
| **Montserrat Suárez** | Catalan Health Service[11] |
| **Eulalia Villegas** | Catalan-Balearic Society of Hospital at Home[12] |
| **Francesc Xavier Jiménez** | Catalan-Balearic Society of Hospital at Home[12] |
| **Mireia Espallargues** | Health Quality and Assessment Agency of Catalonia (AQuAS)[13] |
| **Carme Hernández** | Local JADECARE team |
| **David Nicolás** | Local JADECARE team |

##### 1.2.2 Phases of the qualitative study

| | Objectives | Activities |
|---|---|---|
| ***Session 1*** | — Interpretation of the results of the quantitative analysis. | a. First focus group. |
| ***Session 2*** | — Consensus conclusions regarding healthcare value generation of HaH.<br>— Identify challenges on evaluation of case complexities and costs.<br>— Identify sources of heterogeneity among providers.<br>— Provide recommendations for quality assurance of HaH delivery after service adoption. | b. Survey (structured questionnaire).<br>c. Second focus group. |

##### 1.2.3 Characteristics of the First Focus Group

**Date:** November 15, 2022

**Duration:** 1.5 hours

**Modality:** Virtual session (recorded after agreement of all participants)

**Instruments and technologies:**

- Teams platform for the virtual meeting
- Power Point presentation shared in real-time through both the screen and a link to a SharePoint for editing.

**Agenda:**

- Overall goals, roles, and rules
- Brief introduction to the study
- Interpretation of the quantitative analysis
- Wrap up

##### 1.2.4 Pre-session survey: structured questionnaire

**Instruments and technologies**: Google Forms (anonymous response).

**Content** (as it was displayed to panelists):

**Introduction**

This survey continues the journey we started together on November 15. The contributions during the first working session helped to interpret the results of the quantitative analysis and suggested that it will be necessary to carry out a deeper and more localized study in order to understand all the nuances that lie behind the concept of home hospitalization in Catalonia.

The next session will serve, among other things, to draw conclusions (ideally in the form of recommendations) that will be useful to an international audience and, therefore, as independent as possible from local contexts.

In order to make the session as flexible as possible, we ask you to fill in this form before Monday 12 December at noon. It is designed to be completed in less than 10 minutes.

The survey is anonymous.

**Economic evaluation and efficiency**

Although quantitative measures of efficiency do exist, the scientific literature is heterogeneous in this regard and there is room for bias in their evaluation.

*Rate the following statements related to economic or resource efficiency on a scale of 1 to 5, where 1 indicates "totally disagree" and 5 "totally agree".*

| *Statements* | 1 | 2 | 3 | 4 | 5 |
|---|---|---|---|---|---|
| *In general, home hospitalization generates savings for the health system.* | | | | | |
| *Given the lack of detailed information on costs (cost accounting), the case-mix tools currently used (e.g., DRGs) provide adequate information to assess the economic impact of hospitalization at home.* | | | | | |

**Value contribution**

Over the last few years, as home hospitalization practices have increased, several advantages of home hospitalization have been identified. However, the contribution of home hospitalization may be uneven in each of them.

For each of the following values identified in the literature in relation to home hospitalization in general, indicate what relative level of impact can be expected from the implementation of a home hospitalization service.

*A score of 1 indicates an irrelevant impact on the achievement of this value or goal and 5 indicates a major impact on the achievement of this value.*

| *Statements* | *1* | *2* | *3* | *4* | *5* |
|---|---|---|---|---|---|
| *Adding overall value to the health care system* | | | | | |
| *Increasing bed capacity* | | | | | |
| *Promoting continuity of care* | | | | | |
| *Increasing user satisfaction* | | | | | |
| *Reducing hospitalization costs* | | | | | |
| *Improving health outcomes* | | | | | |

**General key performance indicators**

The document *Organisational model for home hospitalization in Catalonia* identifies a list of indicators (structure, process and outcome) to be used to evaluate the home hospitalization service. Some of these indicators consider territorial aspects or have a level of exhaustiveness that is possible to achieve in Catalonia due to the organizational characteristics of the health system.

However, given that the article is intended to have an international scope, applicable to almost any organizational and territorial model, you are asked to identify the indicators (KPIs) that are considered essential for the evaluation and monitoring of home hospitalization in any health care provider.

***Structure indicators***

*Rate on a scale from 1 to 5 the relevance of the following indicators as generalizable KPIs for the evaluation of the quality of home hospitalization. 1 indicates "irrelevant" and 5 "very relevant".*

| *Statements* | *1* | *2* | *3* | *4* | *5* |
|---|---|---|---|---|---|
| *Number of hospital beds at home.* | | | | | |
| *Profile and number of professionals assigned to full-time home hospitalization* | | | | | |
| *Own and specific equipment (diagnostic and therapeutic ones to be able to attend the patient during hospitalization at home).* | | | | | |
| *Information systems that ensure connectivity from home* | | | | | |

*If you find any missing structure indicator, write here other ones that you consider very relevant and generalizable:* ___________________________________________

***Process indicators***

*Rate on a scale from 1 to 5 the relevance of the following indicators as generalizable KPIs for the evaluation of the quality of home hospitalization. 1 indicates "irrelevant" and 5 "very relevant".*

| *Statements* | *1* | *2* | *3* | *4* | *5* |
|---|---|---|---|---|---|
| *Average number of visits* | | | | | |
| *Annual number of admissions at home* | | | | | |
| *Percentage of visits to the Emergency Room (without admission) during home stay* | | | | | |

| |
|---|
| *Origin of admission in home hospitalization* |
| *Average/median stay* |
| *Destination upon discharge* |

*If you find any missing process indicator, write here other ones that you consider very relevant and generalizable:* ____________________________________________

***Outcome indicators***

*Rate on a scale from 1 to 5 the relevance of the following indicators as generalizable KPIs for the evaluation of the quality of home hospitalization. 1 indicates "irrelevant" and 5 "very relevant".*

| *Statements* | 1 | 2 | 3 | 4 | 5 |
|---|---|---|---|---|---|
| *Complexity of the admitted patient, according to a "case-mix" assessment of health risk stratification.* | | | | | |
| *Difference in length of stay between conventional and home hospitalization* | | | | | |
| *30-Day readmission rate for the same reason* | | | | | |
| *Number of visits to hospital ER at 30 days for the same reason* | | | | | |
| *Mortality rate during the episode* | | | | | |
| *Adverse event rate (pressure ulcers, catheter sepsis, serious adverse drug reactions, episodes of delirium, cardiorespiratory arrests).* | | | | | |
| *Patient/caregiver experience* | | | | | |

*If you find any missing outcome indicator, write here other ones that you consider very relevant and generalizable:* ____________________________________________

##### 1.2.5 Characteristics of the second focus group

**Date:** December 14, 2022

**Duration:** 1.5 hours

**Modality:** Virtual session (recorded after agreement of all participants)

**Instruments and technologies:**

- Teams platform for the virtual meeting
- Power Point presentation shared in real-time through both the screen and a link to a SharePoint for editing. The power point presented the results of the structured questionnaire anonymously.

**Agenda:**

- Overall goals, roles, and rules
- Addressing disagreements
- Conclusions and recommendations
- Final steps

### 2 Supplementary Results

#### 2.1 Supplementary Results of the Quantitative study

*Adoption of hospital at home*

**Table 1S** depicts yearly AA and ESD activity for each health provider.

*Heterogeneities among providers*

**Figures 1S-8S** depict the descriptive analysis of heterogeneities among health providers.

*Cluster analysis*

**Table 2S** depicts the summary results of the four clusters of health providers identified.

Cluster 1 included all seven community hospitals (P21-P27) and P16. Patients, 76.2 [4.0] years, showed both age and AMG scoring slightly above their respective providers. Complexity of the episodes was on the group average and length of stay was the lowest.

Cluster 2 included five out of eight high-tech hospitals and P9. Patients had similar age, 75.7 [5.0] years, but they had more complexity than Cluster 1. This second cluster showed the highest complexity (AMG) and severity of episodes (CMI) of all four groups. The patients were also older and more complex relative to their respective providers.

Cluster 3 including the majority, nine out of twelve, of the general hospitals, this cluster showed values close to average for most indices, compared to the other clusters and to their respective hospitals. Interestingly, the group displayed the lowest percent of repeaters.

Cluster 4 is composed of four hospitals, three from high-tech hospitals (P1, P2 and P5) and one from general hospitals (P11). Patients included in the group are the youngest, with lower complexity before admission and less severe episodes (AMG and CMI, respectively) compared to other groups, but also to their respective providers. However, they showed the longest LoS.

**Table 3S-4S** depict the characteristics of the four clusters of health providers identified.

TABLE 1S – Number of admissions in of Admission Avoidance (AA) and Early Supported Discharge (ESD) registered in 27 health providers from Catalonia between 2015 and 2019.

Number of annual admissions in AA and ESD programs arranged by health providers. The health centers are classified into three categories based on the number of hospital beds and their role in their corresponding health district: High-technology hospitals, General Hospitals and Community Hospitals. The hospitals are ranked within their categories according to the total number of admissions. H stands for hospital, HU for University Hospital and PS for Health Complex.

| | | AA | | | | | | ESD | | | | | | GRAND TOTAL |
|---|---|---|---|---|---|---|---|---|---|---|---|---|---|---|
| | | **2015** | **2016** | **2017** | **2018** | **2019** | **Total** | **2015** | **2016** | **2017** | **2018** | **2019** | **Total** | |
| | **HIGH-TECHNOLOGY HOSPITALS** | | | | | | | | | | | | | |
| P1 | H. U. Arnau de Vilanova de Lleida | 435 | 480 | 607 | 628 | 660 | 2,810 | 334 | 319 | 358 | 366 | 385 | 1,762 | 4,572 |
| P2 | H. de Sabadell | 158 | 150 | 165 | 172 | 216 | 861 | 569 | 597 | 744 | 664 | 646 | 3,220 | 4,081 |
| P3 | H. U. Germans Trias i Pujol de Badalona | 278 | 260 | 210 | 297 | 250 | 1,295 | 543 | 570 | 498 | 568 | 562 | 2,741 | 4,036 |
| P4 | H. Clinic de Barcelona | 377 | 405 | 564 | 750 | 706 | 2,802 | 136 | 116 | 172 | 258 | 201 | 883 | 3,685 |
| P5 | H. U. Joan XXIII de Tarragona | 243 | 281 | 280 | 291 | 295 | 1,390 | 504 | 378 | 386 | 296 | 352 | 1,916 | 3,306 |
| P6 | H. U. de Bellvitge | 152 | 148 | 119 | 95 | 87 | 601 | 411 | 416 | 459 | 501 | 527 | 2,314 | 2,915 |
| P7 | H. U. Vall Hebron | 256 | 237 | 220 | 211 | 237 | 1,161 | 366 | 313 | 198 | 295 | 233 | 1,405 | 2,566 |
| P8 | H. del Mar | 62 | 70 | 130 | 321 | 410 | 993 | 70 | 49 | 71 | 71 | 70 | 331 | 1,324 |
| | **High-technology hospitals TOTAL** | **1,961** | **2,031** | **2,295** | **2,765** | **2,861** | **11,913** | **2,933** | **2,758** | **2,886** | **3,019** | **2,976** | **14,572** | **26,485** |
| | **GENERAL HOSPITALS** | | | | | | | | | | | | | |
| P9 | H.Dos de Maig | 312 | 346 | 511 | 667 | 623 | 2,459 | 117 | 127 | 142 | 198 | 175 | 759 | 3,218 |
| P10 | H. de Hospitalet Moises Broggi | 271 | 293 | 336 | 302 | 333 | 1,535 | 227 | 232 | 258 | 230 | 224 | 1,171 | 2,706 |
| P11 | H. U. de Vic | 105 | 122 | 122 | 149 | 212 | 710 | 374 | 393 | 412 | 383 | 366 | 1,928 | 2,638 |
| P12 | H.de Viladecans | 280 | 227 | 226 | 184 | 183 | 1,100 | 294 | 291 | 306 | 327 | 285 | 1,503 | 2,603 |
| P13 | P. S. Sant Joan de Deu H. General | 156 | 209 | 278 | 292 | 275 | 1,210 | 245 | 243 | 288 | 275 | 242 | 1,293 | 2,503 |
| P14 | H.de Mataro | 272 | 226 | 279 | 368 | 372 | 1,517 | 112 | 122 | 90 | 101 | 119 | 544 | 2,061 |
| P15 | H. U. Mutua de Terrassa | 193 | 194 | 138 | 178 | 441 | 1,144 | 108 | 104 | 105 | 130 | 111 | 558 | 1,702 |
| P16 | H. de Tortosa Verge de la Cinta | 344 | 276 | 272 | 275 | 256 | 1,423 | 39 | 21 | 23 | 28 | 21 | 132 | 1,555 |
| P17 | H. de Mollet | 82 | 125 | 178 | 166 | 150 | 701 | 122 | 88 | 97 | 88 | 85 | 480 | 1,181 |
| P18 | H. de Terrassa | 0 | 0 | 0 | 0 | 439 | 439 | 0 | 0 | 0 | 0 | 50 | 50 | 489 |
| P19 | H. General de Granollers | 0 | 0 | 9 | 176 | 187 | 372 | 0 | 0 | 0 | 0 | 6 | 6 | 378 |
| P20 | H. Sant Joan de Deu (Martorell) | 0 | 0 | 0 | 65 | 71 | 136 | 0 | 0 | 0 | 13 | 9 | 22 | 158 |
| | **General hospitals TOTAL** | **2,015** | **2,018** | **2,349** | **2,822** | **3,542** | **12,746** | **1,638** | **1,621** | **1,721** | **1,773** | **1,693** | **8,446** | **21,192** |

| | | | | | | | | | | | | | | |
|---|---|---|---|---|---|---|---|---|---|---|---|---|---|---|
| **COMMUNITY HOSPITALS** | | | | | | | | | | | | | | |
| P21 | H. Sant Jaume de Calella i H. de Blanes | 262 | 514 | 576 | 755 | 805 | 2,912 | 67 | 167 | 146 | 236 | 188 | 804 | 3,716 |
| P22 | Althaia H. de Sant Joan de Deu | 206 | 197 | 230 | 253 | 255 | 1,141 | 218 | 186 | 149 | 192 | 255 | 1,000 | 2,141 |
| P23 | Fundacio H. de Esperit Sant | 160 | 219 | 299 | 283 | 274 | 1,235 | 99 | 157 | 159 | 165 | 130 | 710 | 1,945 |
| P24 | Pius H. de Valls | 211 | 209 | 197 | 185 | 204 | 1,006 | 88 | 110 | 134 | 107 | 76 | 515 | 1,521 |
| P25 | H. Municipal de Badalona | 167 | 77 | 63 | 111 | 100 | 518 | 132 | 75 | 75 | 62 | 33 | 377 | 895 |
| P26 | H. de Sant Celoni | 51 | 64 | 60 | 58 | 45 | 278 | 61 | 51 | 25 | 28 | 28 | 193 | 471 |
| P27 | H. Comarcal de Blanes | 152 | 0 | 0 | 0 | 0 | 152 | 29 | 0 | 0 | 0 | 0 | 29 | 181 |
| | **Community hospitals TOTAL** | **1,209** | **1,280** | **1,425** | **1,645** | **1,683** | **7,242** | **694** | **746** | **688** | **790** | **710** | **3,628** | **10,870** |
| **GRAND TOTAL** | | **5,185** | **5,329** | **6,069** | **7,232** | **8,086** | **31,901** | **5,265** | **5,125** | **5,295** | **5,582** | **5,379** | **26,646** | **58,547** |

TABLE 2S – Comparison of the patient's clinical characteristics at admission and the complexity of the intervention in Admission Avoidance (AA) programs among clusters of providers. The clusters are identified through K-means clustering methods considering the age, the patient's morbidity burden assessed with the Adjusted Morbidity Groups (AMG) score, the selection bias by measuring the distance in mean age and AMG score between AA and conventional hospitalizations, the length of stay (LoS), the complexity of the episode assessed with the Case Mix Index (CMI), the patient reiteration rate and the category of the hospital based on the number of hospital beds and their role in their corresponding health district.

| # cluster | n | Age, mean (sd) | AMG, mean (sd) | CMI | LoS, mean (sd) | Distance Age, mean (sd) | Distance AMG, mean (sd) | Repeaters, n (%) | Providers |
|---|---|---|---|---|---|---|---|---|---|
| 1 | 8 | 76.2 (4.08) | 29.83 (3.08) | 0.65 (0.04) | 7.96 (3.2) | 2.22 (4.32) | 3.83 (3.19) | 21.65 (6.43) | P16; P21; P22; P23; P24; P25; P26; P27 |
| 2 | 6 | 75.67 (5.01) | 34.38 (4.07) | 0.7 (0.03) | 9.8 (2) | 3.27 (3.04) | 6.52 (4.23) | 23.76 (5.62) | P3; P4; P6; P7; P8; P9 |
| 3 | 9 | 71.51 (3.42) | 29.14 (3.05) | 0.69 (0.08) | 8.28 (1.5) | -2.02 (2.98) | 0.51 (2.6) | 12.03 (7.54) | P10; P12; P13; P14; P15; P17; P18; P19; P20 |
| 4 | 4 | 65.21 (3.3) | 22.05 (1.91) | 0.6 (0.05) | 9.96 (2.71) | -3.84 (3.34) | -1.12 (2.51) | 20.02 (9.4) | P1; P2; P5; P11 |

TABLE 3S – Patient's clinical characteristics and outcomes of the intervention of all the patients admitted in hospital admission avoidance programs arranged by clusters of providers.

Patients' information is itemized into five categories, covering the patient's clinical characteristics at admission, the utilization of healthcare resources 12 months before the admission, the hospitalization episode, and the health outcomes of the intervention assessed at 30 and 90 days post-discharge. The morbidity burden is assessed with the AMG score, and the complexity of the hospitalization is assessed with the Queralt Index. The health providers are distributed among clusters as follows: Cluster 1: P16, P21, P22, P23, P24, P25, P26, P27; Cluster 2: P3, P4, P6, P7, P8, P9; Cluster 3: P10, P12, P13, P14, P15, P17, P18, P19, P20; Cluster 4: P1, P2, P5, P11. AMG stands for Adjusted Morbidity Groups and HRSN for health-related social needs.

| | Cluster 1<br>n = 8,665 | Cluster 2<br>n = 9,311 | Cluster 3<br>n = 8,154 | Cluster 4<br>n = 5,771 |
|---|---|---|---|---|
| **DEMOGRAPHICS & MORBIDITY-COMPLEXITY** | | | | |
| **Age, mean(sd)** | 76.81 (14.8) | 75.87 (14.95) | 70.46 (17.11) | 66.8 (19.02) |
| **Gender, n(%)** | | | | |
| ***Male*** | 4,432 (51.15) | 4,402 (47.28) | 3,587 (43.99) | 2,793 (48.4) |
| ***Female*** | 4,233 (48.85) | 4,909 (52.72) | 4,567 (56.01) | 2,978 (51.6) |
| **AMG, mean(sd)** | 30.64 (15.2) | 33.68 (16.85) | 28.25 (17.06) | 22.84 (14.39) |
| **AMG category, n (%)** | | | | |
| ***Very low risk < $P_{50}$*** | 41 (0.47) | 44 (0.47) | 85 (1.04) | 96 (1.66) |
| ***Low risk [$P_{50}$ - $P_{80}$)*** | 309 (3.57) | 283 (3.04) | 563 (6.9) | 612 (10.6) |
| ***Moderate risk [$P_{80}$-$P_{95}$)*** | 1,030 (11.89) | 920 (9.88) | 1,443 (17.7) | 1,329 (23.03) |
| ***High risk [$P_{95}$-$P_{99}$)*** | 1,489 (17.18) | 1,432 (15.38) | 1,435 (17.6) | 1,120 (19.41) |
| ***Very high risk ≥ $P_{99}$*** | 5,796 (66.89) | 6,632 (71.23) | 4,628 (56.76) | 2,614 (45.3) |
| **Patients with HRSN associated to housing and economic conditions, n (%)** | 1,881 (21.71) | 1,683 (18.08) | 919 (11.27) | 576 (9.98) |
| **Patients with HRSN associated to family and social environment, n (%)** | 2,751 (31.75) | 3,901 (41.9) | 2,038 (24.99) | 1,210 (20.97) |
| **Patients receiving palliative care, n (%)** | 466 (5.38) | 643 (6.91) | 168 (2.06) | 160 (2.77) |
| **USE OF RESOURCES 12 MONTHS BEFORE ADMISSION** | | | | |
| **Patients requiring hospital admissions, n (%)** | 4,442 (51.5) | 5,210 (56.12) | 3,675 (45.13) | 2,622 (45.65) |
| **Patients requiring emergency room visits, n (%)** | 7,413 (85.94) | 7,704 (82.98) | 6,334 (77.78) | 4,349 (75.71) |
| **Total Expenditure in €, median ($P_{25}$-$P_{75}$)** | 4,137.85 (1,868.09 - 7,854.48) | 5,493.6 (2,278.53 - 10,664.12) | 3,518.25 (1,322.13 - 7,481.88) | 3,320.77 (1,173.94 - 7,064.05) |

| | | | | |
|---|---|---|---|---|
| **HOME HOSPITALIZATION EPISODE** | | | | |
| **LoS, mean(sd)** | 6.92 (5.44) | 9.11 (6.39) | 8.27 (5.57) | 10.02 (7.81) |
| **Mortality, n(%)** | 39 (0.45) | 27 (0.29) | 10 (0.12) | 27 (0.47) |
| **Queralt Index, mean (sd)** | 29.06 (14.67) | 32.46 (15.04) | 29.43 (16.75) | 18.93 (13.43) |
| **Case Mix Index** | 0.66 | 0.7 | 0.66 | 0.62 |
| **USE OF RESOURCES 30 DAYS AFTER DISCHARGE** | | | | |
| **Mortality, n (%)** | 551 (6.39) | 457 (4.92) | 200 (2.46) | 174 (3.03) |
| **Patients requiring hospital admissions, n (%)** | 1,016 (11.78) | 1,148 (12.37) | 806 (9.9) | 636 (11.07) |
| **Patients requiring emergency room visits, n (%)** | 1,735 (20.11) | 1,952 (21.03) | 1,501 (18.43) | 946 (16.47) |
| **Total Expenditure in €, median ($P_{25}$-$P_{75}$)** | 267.01 (116.31 - 664.13) | 340.93 (144.58 - 1,058.96) | 243.12 (106.27 - 591.5) | 260.44 (111.65 - 800.37) |
| **USE OF RESOURCES 90 DAYS AFTER DISCHARGE** | | | | |
| **Mortality, n (%)** | 1,098 (12.73) | 1,007 (10.85) | 487 (5.98) | 359 (6.25) |
| **Patients requiring hospital admissions, n (%)** | 2,007 (23.27) | 2,431 (26.18) | 1,690 (20.75) | 1,205 (20.98) |
| **Patients requiring emergency room visits, n (%)** | 3,235 (37.5) | 3,783 (40.75) | 2,967 (36.43) | 1,802 (31.37) |
| **Total Expenditure in €, median ($P_{25}$-$P_{75}$)** | 842.55 (350.1 - 2541.8) | 1,233.1 (487.06 - 3297.74) | 791.95 (323.85 - 2328.94) | 818.43 (307.02 - 2707) |

TABLE 4S – Comparison of patients' clinical characteristics and the outcomes of the intervention between individuals admitted in hospital admission avoidance programs and their corresponding matched controls of conventional hospitalizations arranged by clusters of providers.

Patients' characteristics are itemized into three categories, covering the patient's clinical characteristics at admission, the hospitalization episode, and the health outcomes of the intervention assessed at 30 post-discharge. The morbidity burden is assessed with the AMG score and the complexity of the hospitalization is assessed with the Queralt Index. The health providers are distributed among clusters as follows: Cluster 1: P16, P21, P22, P23, P24, P25, P26, P27; Cluster 2: P3, P4, P6, P7, P8, P9; Cluster 3: P10, P12, P13, P14, P15, P17, P18, P19, P20; Cluster 4: P1, P2, P5, P11. AMG stands for Adjusted Morbidity Groups.

| | **Cluster 1** | | **Cluster 2** | | **Cluster 3** | | **Cluster 4** | |
|---|---|---|---|---|---|---|---|---|
| | **Matched AA**<br>**n = 6,264** | **Matched control**<br>**n = 6,264** | **Matched AA**<br>**n = 7,374** | **Matched control**<br>**n = 7,374** | **Matched AA**<br>**n = 6,673** | **Matched control**<br>**n = 6,673** | **Matched AA**<br>**n = 4,491** | **Matched control**<br>**n = 4,491** |
| **DEMOGRAPHICS & MORBIDITY-COMPLEXITY** | | | | | | | | |
| **Age, mean(sd)** | 76.09 (14.79) | 75.7 (14.45) | 74.92 (15.27) | 75 (15.41) | 71.54 (16.69) | 71.19 (16.65) | 68.54 (18.05) | 67.13 (17.81) |
| **Gender, n(%)** | | | | | | | | |
| ***Male*** | 3,131 (49.98) | 3,151 (50.3) | 3,380 (45.84) | 3,661 (49.65) | 2,968 (44.48) | 3,153 (47.25) | 2,326 (51.79) | 2,019 (44.96) |
| ***Female*** | 3,133 (50.02) | 3,113 (49.7) | 3,994 (54.16) | 3,713 (50.35) | 3,705 (55.52) | 3,520 (52.75) | 2,165 (48.21) | 2,472 (55.04) |
| **AMG, mean(sd)** | 28.83 (14.7) | 28.67 (14.88) | 31.55 (16.11) | 31.43 (16.15) | 27.91 (16.58) | 26.99 (16) | 23.11 (14.15) | 22.22 (14.49) |
| **HOME HOSPITALIZATION EPISODE** | | | | | | | | |
| **LoS, mean(sd)** | 6.75 (4.88) | 7.27 (5.98) | 9.19 (6.13) | 7.76 (6.32) | 8.32 (5.51) | 6.74 (5.35) | 9.85 (7.42) | 6.26 (5.29) |
| **Mortality, n(%)** | 28 (0.45) | 30 (0.48) | 18 (0.24) | 38 (0.52) | 8 (0.12) | 24 (0.36) | 22 (0.49) | 20 (0.45) |
| **Queralt Index, mean(sd)** | 28.07 (14.44) | 35.37 (19.56) | 31.91 (15.12) | 41.28 (21.99) | 29.6 (16.09) | 38.96 (22.9) | 19.31 (13.41) | 27.61 (20.73) |
| **Case Mix Index** | 0.65 | 0.73 | 0.68 | 0.79 | 0.66 | 0.75 | 0.62 | 0.7 |
| **USE OF RESOURCES 30 DAYS AFTER DISCHARGE** | | | | | | | | |
| **Mortality, n (%)** | 353 (5.66) | 338 (5.42) | 311 (4.23) | 344 (4.69) | 168 (2.52) | 255 (3.84) | 141 (3.16) | 175 (3.91) |
| **Patients requiring hospital admissions, n (%)** | 527 (8.45) | 391 (6.27) | 737 (10.02) | 628 (8.56) | 477 (7.16) | 322 (4.84) | 440 (9.85) | 284 (6.35) |
| **Patients requiring emergency room visits, n (%)** | 1050 (16.84) | 953 (15.29) | 1350 (18.35) | 1408 (19.19) | 1061 (15.92) | 990 (14.89) | 648 (14.5) | 617 (13.8) |
| **Total Expenditure in €, median ($P_{25}$-$P_{75}$)** | 244.36 (113.1 - 532.12) | 213.18 (86.63 - 433.41) | 304.83 (133.67 - 782.24) | 286.28 (121.02 - 641.13) | 228.99 (102.21 - 492.29) | 212.3 (96.5 - 414.74) | 253.07 (112.07 - 713.05) | 221.26 (100.6 - 481.77) |

FIGURE 1S - Comparison of the patient's age among providers in hospital admission avoidance programs.

The red line corresponds to all providers' mean patient's age, and the Whisker boundaries are set to 1.5 x IQR.

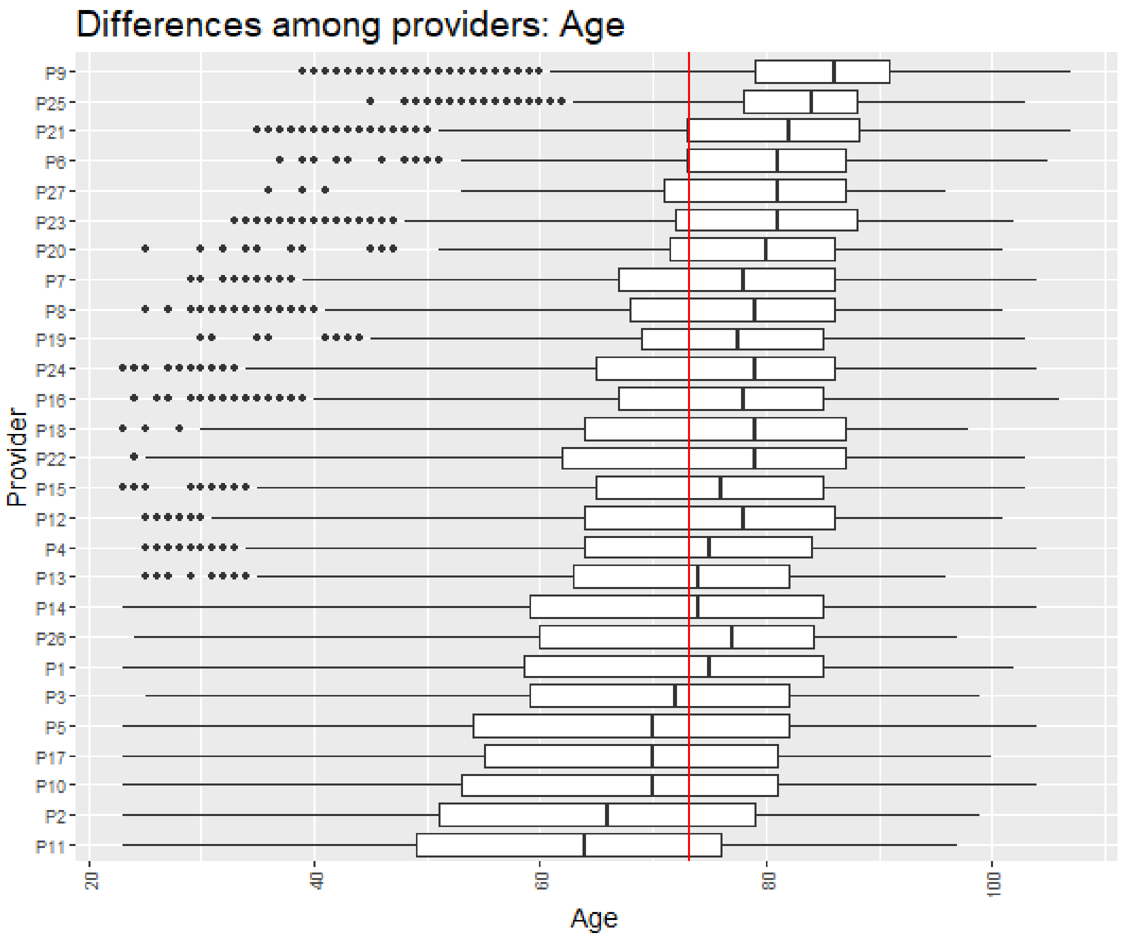

FIGURE 2S - Comparison of the patient's clinical complexity assessed with the Adjusted Morbidity Groups (AMG) score among providers in hospital admission avoidance programs.

The red line corresponds to all providers' mean AMG, and the Whisker boundaries are set to 1.5 x IQR.

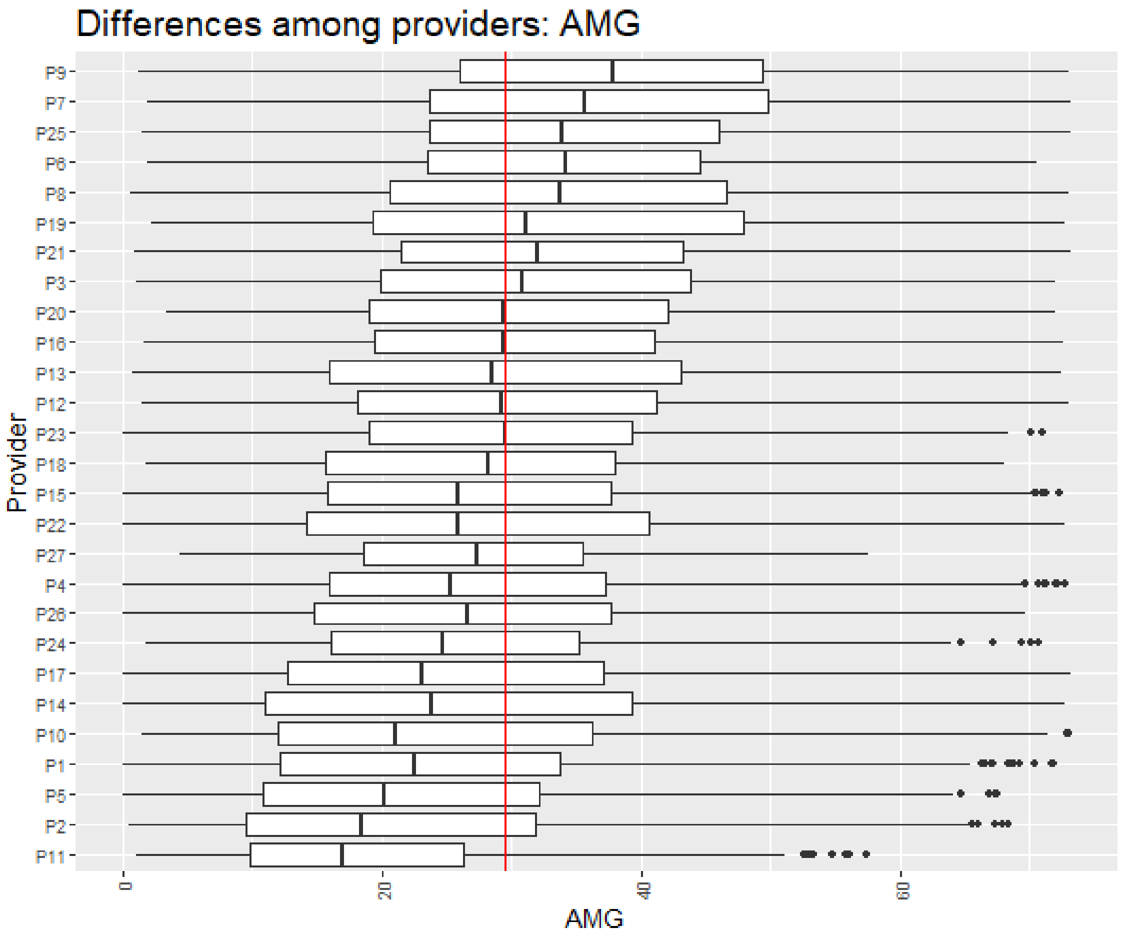

FIGURE 3S - Comparison of the hospitalization complexity among providers assessed with the APR-DRG score in hospital admission avoidance programs.

The red line corresponds to all providers' mean Case Mix Index, and the Whisker boundaries are set to 1.5 x IQR.

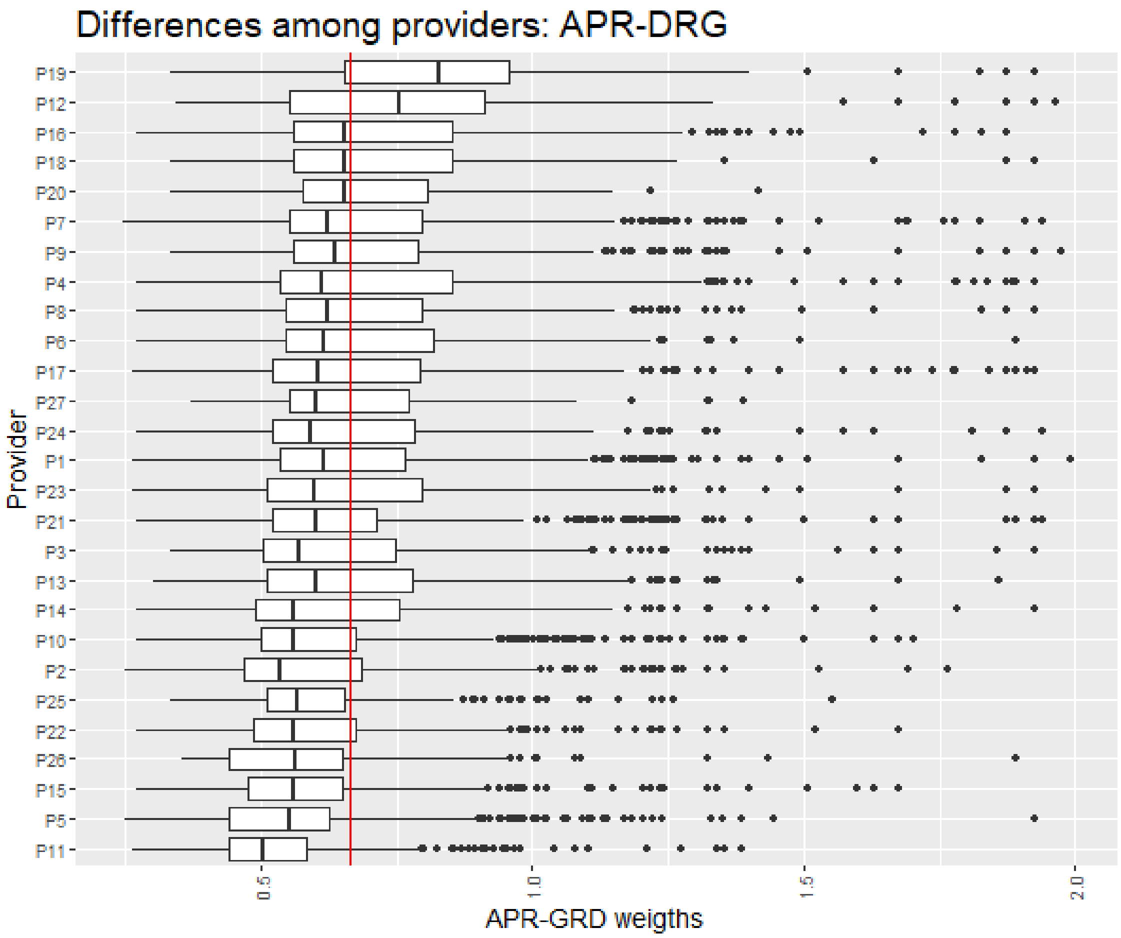

FIGURE 4S - Comparison of the length of stay among providers in hospital admission avoidance programs.

The red line corresponds to all providers' mean length of stay, and the Whisker boundaries are set to 1.5 x IQR. LoS stands for length of stay.

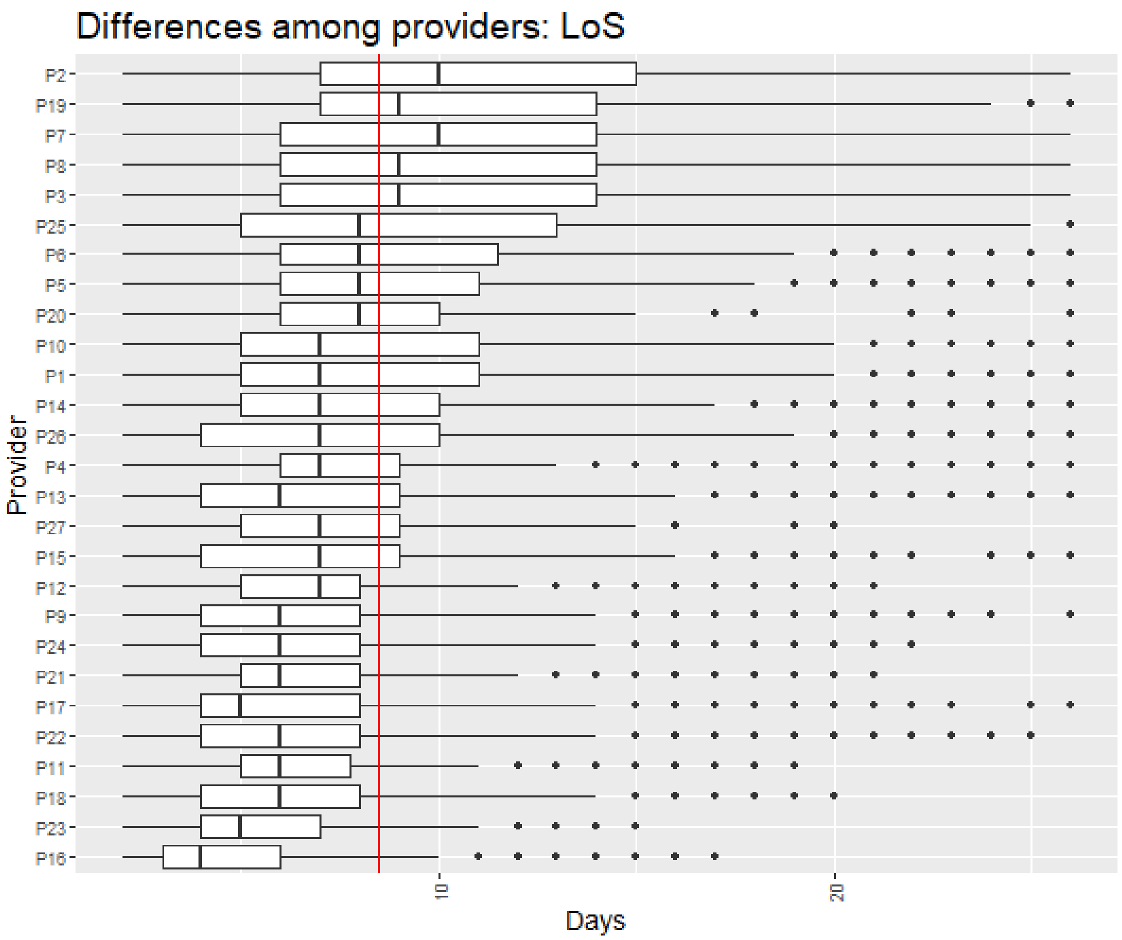

FIGURE 5S – Comparison of the hospitalization complexity among providers assessed with the Queralt index in hospital admission avoidance programs.

The red line corresponds to all providers' mean Queralt index score, and the Whisker boundaries are set to 1.5 x IQR.

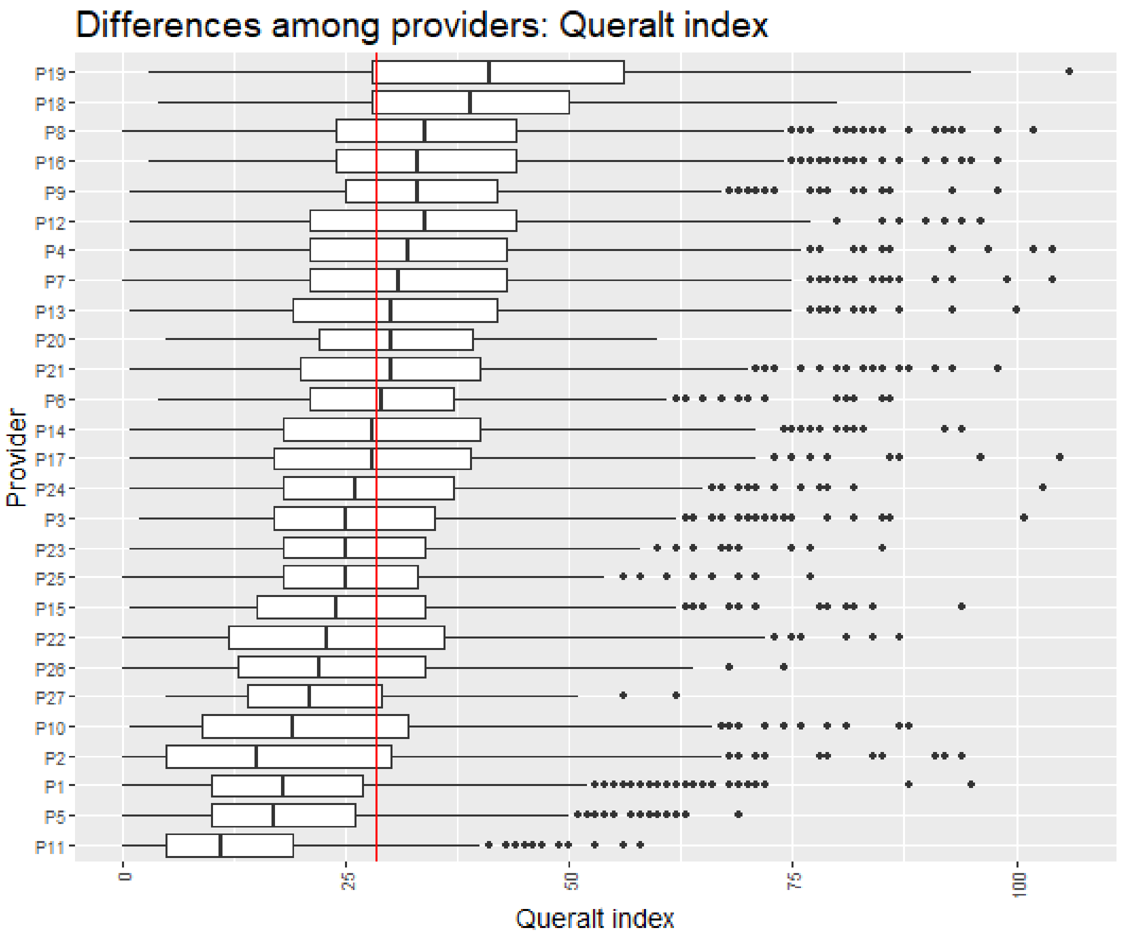

FIGURE 6S – Comparison among providers of the percentages of episodes corresponding to previously admitted patients in the hospital admission avoidance programs.

The red line corresponds to all providers' mean patient reiteration percentage.

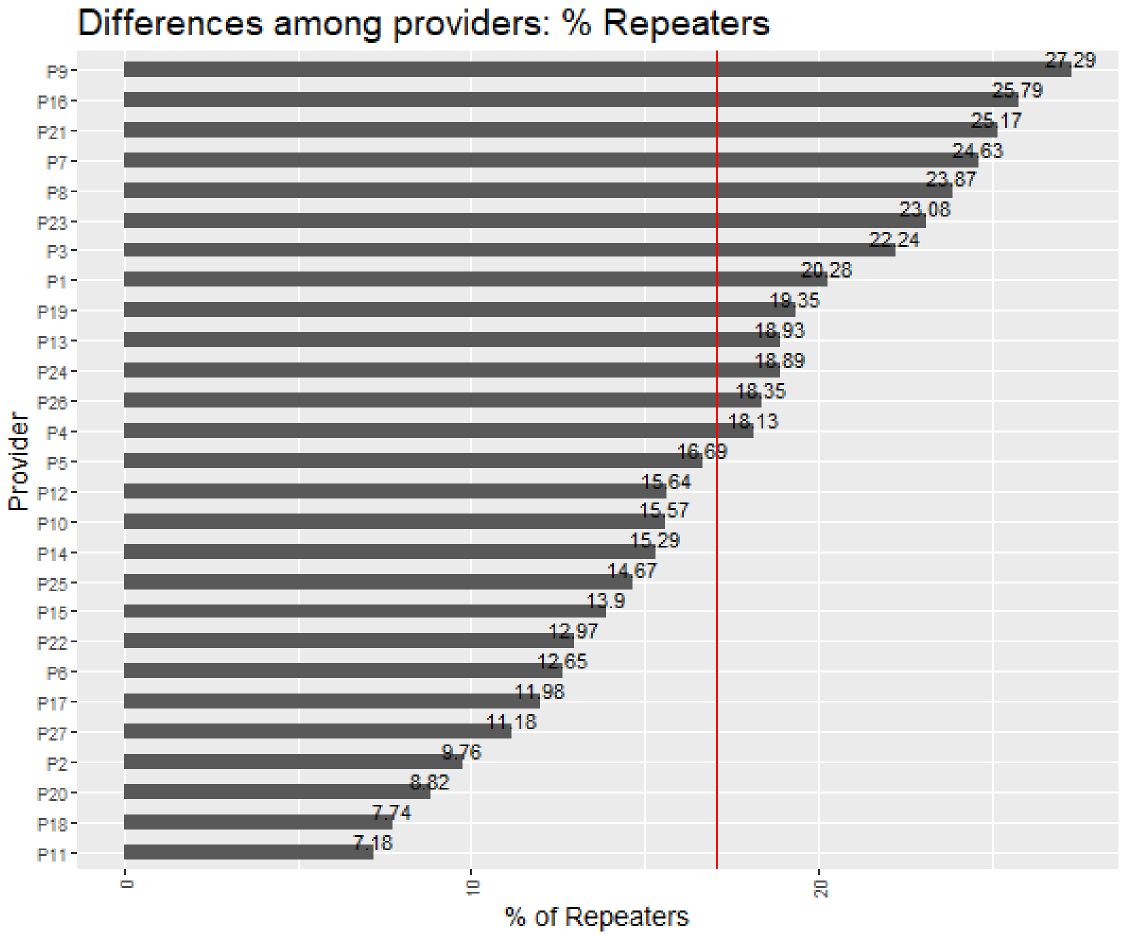

FIGURE 7S – Comparison among providers of the distance between the average age of the patients admitted under hospital admission avoidance and conventional hospitalization regimes before the matching.

The providers in which the patients admitted in hospital admission avoidance programs have a lower age on average respect the patients admitted under conventional hospitalizations for the same cause within the same provider are depicted in light blue. In contrast, the opposite cases are depicted in light red. The red line corresponds to all providers' mean.

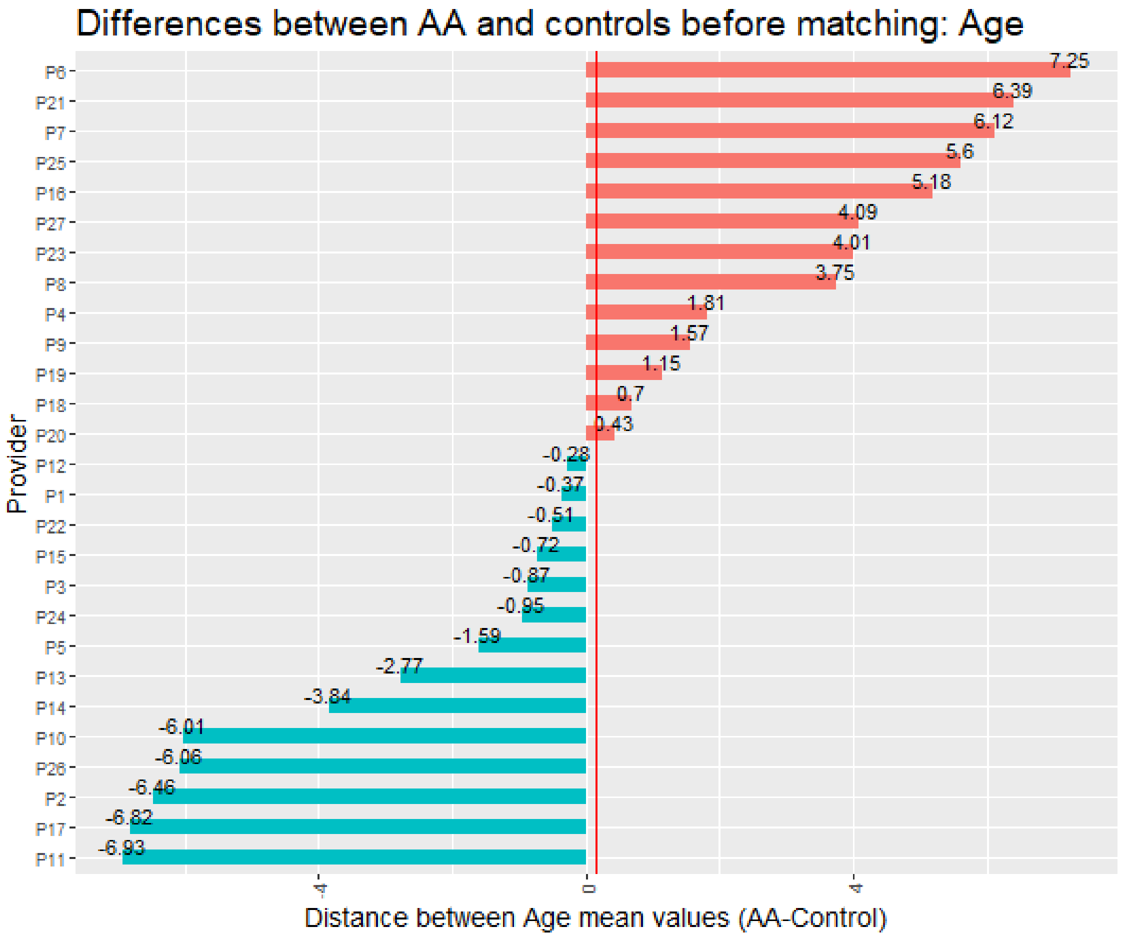

FIGURE 8S – Comparison among providers of the distance between the average clinical complexity, assessed with the AMG score, of the patients admitted under hospital admission avoidance and conventional hospitalization regimes before the matching.

The providers in which the patients admitted in hospital admission avoidance programs have a lower clinical complexity in average respect the patients admitted under conventional hospitalizations for the same cause within the same provider are depicted in light blue, in contrast, the opposite cases are depicted in light red. The red line corresponds to all providers' mean.

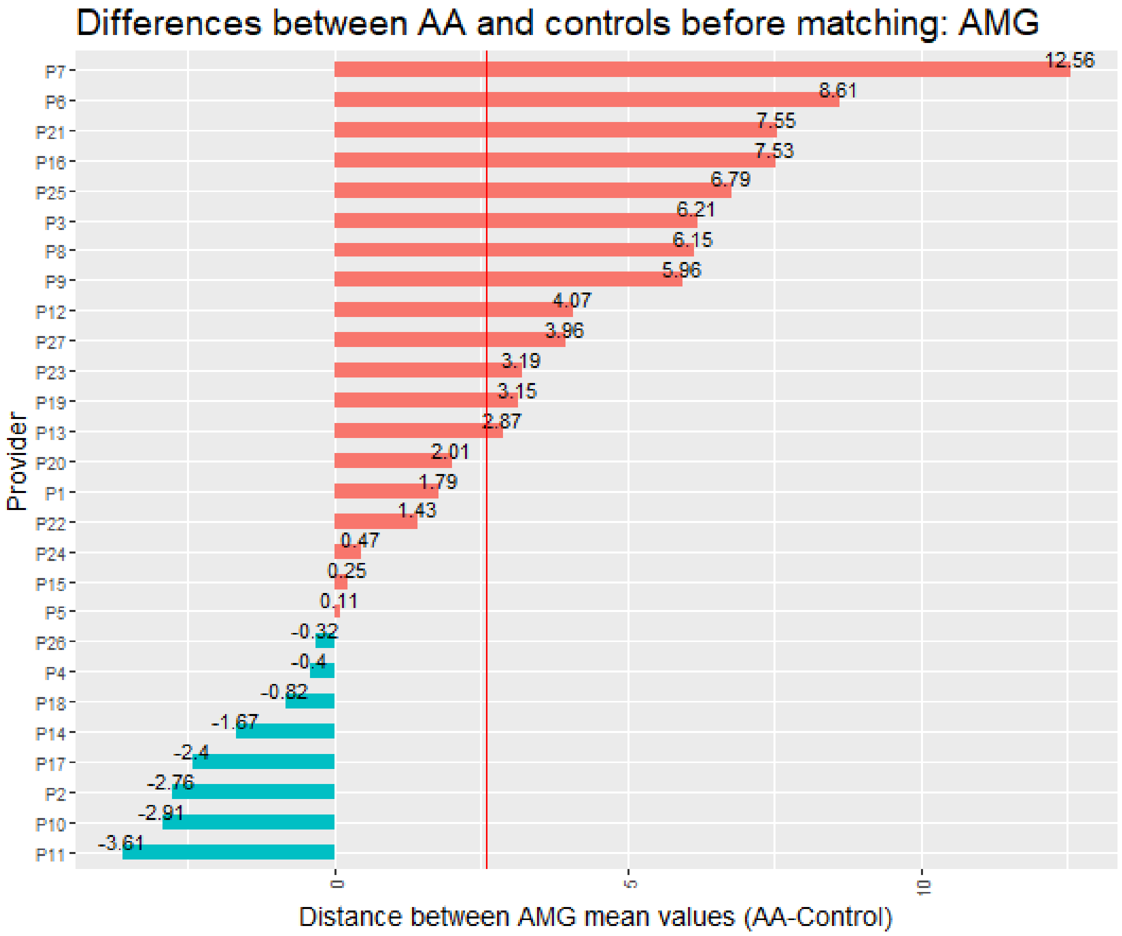

#### 2.2 Supplementary Results of the Qualitative Study

##### 2.2.1 Results of the First Focus Group

###### *2.2.1.1 General conduct and participation*

No technical incidents were reported. The session exceeded by 10 minutes the expected schedule. All experts intervened in each of the result section presented.

###### *2.2.1.2 Highlights*

###### 2.2.1.2.1 Overarching insights

**Definition of the comparator population.** Various experts highlight that, in our local context, some hospitals are reference centers in the area and, therefore, receive referrals from other hospitals to the HaH service. This scenario, which occurs only in some centers, challenges the definition of the catchment population used for the propensity score matching.

**Generalizability and temporal validity of the results.** In 2020, the Catalan Health Service, in collaboration with other stakeholders, conducted the first harmonization process of the service, which ended with the publication of an Organization Model of HaH in the Catalan Health Service on June 2020. Although the publication of the model is very recent, HaH practices observed during the analyzed period may change in the near future. Therefore, a pre-post retrospective analysis or a Quality Improvement Study for assessing the impact of this harmonization should be considered among future works.

**Overall heterogeneity**. It is expected a certain degree of heterogeneity regarding HaH delivery. Different healthcare providers may prioritize different types of interventions or services of HaH (e.g., based on strategic planning, each provider may decide to channeling HaH admissions to a particular process or main diagnosis). This diversity in types of services prioritized may influence not only health outcomes but also the profile of patients admitted to HaH. An in-depth analysis of heterogeneities is, therefore, difficult to be generalizable to an international context. Instead, interviews with providers that take into account the local perspective would be necessary.

###### 2.2.1.2.2 Users' profile: age

- Age is likely to be affected by the overarching highlights described previously, including the referral from other hospitals.
- Based on these heterogeneities, and considering the potential biases associated with overarching highlights , matching for currently accepted indices of clinical complexity, like DRG, might not be sufficient.
- Although data were collected before the issue of the 2020 consensus of the Catalan Health Service on HaH, it is unlikely that age profile has changed. Still, future pre-post studies shall be done to confirm this trend.
- Biases towards younger patients may be associate with limited capacity of some hospitals to attend highly complex (multimorbidity) patients.

###### 2.2.1.2.3 Users' profile: clinical complexity

- Most measures of clinical complexity have been designed for assessing multimorbidity. Although the Adjusted Morbidity Groups also consider acute conditions, the complexity of hospitalizations, which should also consider the procedure or the severity of the primary diagnosis, may not be adequately represented.
- Like age, clinical complexity must be measured by comparing with conventional hospitalization of the same catchment population.
- The complexity profile is consistent with age across providers.

###### 2.2.1.2.4 Recurrence

- Overall, older patients have higher hospitalization frequency. Patients are generally satisfied with HaH; therefore, they very often ask for HaH. This could be the reason why centers with higher mean ages in HaH also have higher recurrence.

###### 2.2.1.2.5 Length of stay

- The extremely short time of the lower limit of the range suggests that some centers are including (as part of HaH service) activities that would not fit the classical definition of HaH (e.g., sample extraction at home).
- Despite the extreme values of the range, the overall median (8 days) seems consistent with the expected time for a HaH service.
- The length of stay analysis does not make sense without adjusting or stratifying for clinical complexity (e.g., AMG) and procedure.

##### 2.2.2 Results of the Second Focus Group

###### *2.2.2.1 General conduct and participation*

No technical incidents were reported. All experts intervened in each of the result section presented.

###### *2.2.2.2 Highlights*

###### 2.2.2.2.1 Economic evaluation and efficiency

- HaH not necessarily reduce the overall cost of the healthcare system (e.g., it may be used to increase the capacity of hospital services) but increases efficiency if implemented appropriately.

- In case of maintaining the total capacity of the hospital, HaH reduces costs, mostly because of the lower need of infrastructure.
- Case-mix tools currently used for payment (e.g., DRGs) are not suitable for benchmarking or cost analysis of HaH because they do not reflect the complexity of the intervention adequately. Other emerging tools, such as the Queralt system, may be more appropriate.
- Without analytical accounting, it is not possible to assess the economic impact of HaH.
- The DRGs may nevertheless provide relevant information.

###### 2.2.2.2.2 Value contribution

- HaH adds overall value to the healthcare system.
- HaH is appropriate to increase hospitalization capacity.
- HaH facilitates the continuity of care.
- In our environment, post-discharge activity is rarely assessed; therefore, it is unknown the efficiency of all activities performed at discharge to ensure the continuity of care.
- HaH increases the satisfaction of patients and their relatives.
- HaH increases the efficiency of the hospital.
- In most cases, HaH improves health results.

###### 2.2.2.2.3 General key performance indicators

- The number of discharges is a better indicator than the number of beds. Rather than the absolute number, the number of discharges relative to the conventional hospitalization in the area should be considered.
- The number and profile of full-time healthcare professionals dedicated to HaH is a good indicator of the quality of the service.
- HaH is a flexible and dynamic service; therefore, the number and profile of healthcare professionals is more relevant than the equipment.
- Professional capacity is among the challenges of HaH. This type of hospitalization requires broad and specialized skills that are not covered by any specialty. Therefore, continuous training is necessary.
- Interoperability though a good integration of information systems is important for HaH service provision.
- The number of visits is not a good indicator of quality because it strongly depends on the patient profile and the type of procedure. Furthermore, this indicator cannot be measured in conventional hospitalization and, therefore, comparisons are not possible. Also, telemedicine can reduce the number of visits without compromising the quality of care.
- The yearly number of admissions should be considered when assessing HaH services.
- Admissions to an intensive care unit during the HaH episode are rare. Conversely, visits to the emergency room and non-scheduled hospital admissions during the episode are a good indicators or the quality of HaH.
- [origin: previous setting]
- The length of stay is a good indicator for hospital avoidance if compared with the same procedure. In the case of early discharge, it is very difficult to appraise. Furthermore, long times of stay in EAD may be due to complications during conventional hospitalizations that increase the overall hospital stay time.
- [Destination on discharge]
- The complexity of admission, according to a case-mix tool, should be considered.

- Mortality is not generalizable as an indicator because some patients may be admitted at home during end-of-life pathways. Non-anticipated mortality could be considered as an indicator.
- Adverse events rate (e.g., ulcer, sepsis) is a good indicator of the quality of care.
- The experience of patients and caregivers or relatives is a good indicator of the quality of care.

### 3 Supplementary References

1. Farré N, Vela E, Clèries M, Bustins M, Cainzos-Achirica M, Enjuanes C, et al. Medical resource use and expenditure in patients with chronic heart failure: a population-based analysis of 88 195 patients. Eur J Heart Fail. 2016 Sep 1;18(9):1132–40.
2. Austin PC. An Introduction to Propensity Score Methods for Reducing the Effects of Confounding in Observational Studies. Multivariate Behav Res. 2011 May;46(3):399.
3. A T, G E, S B, P L, DJ S, S S. Should I stay or should I go? A retrospective propensity score-matched analysis using administrative data of hospital-at-home for older people in Scotland. BMJ Open. 2019 May 1;9(5).
4. Diamond A, Sekhon JS. Genetic Matching for Estimating Causal Effects: A General Multivariate Matching Method for Achieving Balance in Observational Studies. Rev Econ Stat. 2013 Jul 30;95(3):932–45.
5. Mebane WR, Sekhon JS. Genetic Optimization Using Derivatives: The rgenoud Package for R. JSS J Stat Softw. 2011;42.
6. Goldfield N. The evolution of diagnosis-related groups (DRGs): From its beginnings in case-mix and resource use theory, to its implementation for payment and now for its current utilization for quality within and outside the hospital. Qual Manag Health Care. 2010 Jan;19(1):3–16.
7. Bensken WP, Alberti PM, Stange KC, Sajatovic M, Koroukian SM. ICD-10 Z-Code Health-Related Social Needs and Increased Healthcare Utilization. Am J Prev Med. 2022 Apr 1;62(4):e232–41.
8. Dueñas-Espin I, Vela E, Pauws S, Bescos C, Cano I, Cleries M, et al. Proposals for enhanced health risk assessment and stratification in an integrated care scenario. BMJ Open. 2016 Apr 1;6(4):e010301.
9. Monterde D, Vela E, Clèries M. Adjusted morbidity groups: A new multiple morbidity measurement of use in Primary Care. Aten primaria. 2016 Dec 1;48(10):674–82.
10. Vela E, Piera-Jiménez J. Performance of Quantitative Measures of Multimorbidity: A Population-Based Retrospective Analysis. 2021 Mar 1;
11. Catalan Health Service [Internet]. Available from: https://catsalut.gencat.cat/ca/inici/
12. Societat Catalano-Balear d'Hospitalització Domiciliària [Internet]. Available from: http://webs.academia.cat/societats/hospdomicil/index.php
13. Health Quality and Assessment Agency of Catalonia (AQuAS).
