## Supplementary Material S2 for "Five years of Hospital at Home adoption in Catalonia: impact and challenges"

### Organisational model for Hospital at Home in Catalonia

Alternative to conventional hospitalisation

**Catalan Health Service**

June 2020

/Salut Generalitat de Catalunya

**Coordination of the document:**

Casanovas-Guitart, Cristina
Hernández Carcereny, Carme[1]
Villegas Bruguera, Eulàlia B.[2]
Aloy Punzano, John
Ricart Conesa, Assumpta
Guarga Rojas, Àlex

**Working group:**

Benavent Areu, Jaume
Biescas Prat, Herminia
Blanch Andreu, Jordi
Caballero Nieto, Maria
Contel Segura, Juan Carlos
Corominas, Antoni
Costas Muñoz, Emma
Güell Viaplana, Francesc
Inzitari, Marco[3]
Jiménez Moreno, Francesc Xavier
Leal Negre, Mireia
Mas Bergas, Miquel Àngel[3]
Mompart Penina, Anna
Pontes Garcia, Caridad
Reynolds, Jillian
Roca Casas, Jordi Ruiz
Riera, Rafael
Sabaté, Rosa Anna[3]
Santaeugènia Gonzalez, Sebastià
Seijas, Núria
Torras Boatella, Maria Glòria
Torné Vilagrasa, Elvira
Viñas Folch, Paulina

[1] Working group coordinator
[2] On behalf of the Catalan-Balearic Society for Home Hospitalisation (SCBHD).
[3] On behalf of the Catalan Society of Geriatrics and Gerontology (SCGiG).

Index

#### 1. Prologue

Hospital at Home (HaH) is a care model that belongs to the hospital setting and is designed as an alternative to conventional hospitalisation. Its main function is admission avoidance (AA) or to reduce the length of patients' hospital stay (early supported discharge [ESD]).

This care model was an emerging initiative some years ago and has shown, through scientific evidence, to have positive results in terms of mortality, readmissions, costs, and patient and family satisfaction.

In Catalonia, the experience of HaH has been consolidated over the years, and many hospitals have already deployed this care model. However, there is evidence of notable heterogeneity between their implementations. For this reason, the Catalan Health Service (CatSalut) has proposed an in-depth analysis and reflection of this care model to revise, update and specify the contents of the care and portfolio of services, as well as the organisational characteristics of HaH, and reach a consensus on the best organisational model for HaH in Catalonia.

Likewise, there is a need to set up integrated territorial care between the different care levels, with a well-defined portfolio of services in each case, where complementarity and continuity of care ensure safe transitions and facilitate maximum resolution at each level, especially in the case of primary and community care and intermediate care.

To do so, experts, scientific societies, as well as other professionals from primary and community care to intermediate care, and health planning managers and referents from the Catalan Department of Health were invited to participate.

As a result, a care delivery model has been defined and proposed with common criteria and oriented to the territory, which should allow for the provision of quality and assessable services and, on the other hand, facilitate orderly planning of the allocation of resources with criteria of efficiency and sustainability. It is also of great importance to place the provision of services in the clinical and care process linked to primary and community care, emergencies, and intermediate care, where the aim is to consolidate a portfolio of services of the highest resolution and to strengthen the role and leadership of the nurse in the entire care process.

#### 2. Acronyms

| | |
|---|---|
| Admission avoidance | AA |
| Advanced chronic disease | MACA (for Catalan “malaltia crònica avançada”) |
| Advanced life support | ALS |
| Automated external defibrillators | AED |
| Basic health area | ABS (for Catalan “àrea bàsica de salut) |
| Primary homecare program | ATDOM |
| Basic life support | BLS |
| Bi-level positive airway pressure | BiPAP-BPAP |
| Cardiopulmonary resuscitation | CPR |
| Care management area | AGA (for Catalan “àrea de gestió assistencial”) |
| Agency for Health Quality and Assessment of Catalonia | AQuAS (Agència de Qualitat i Avaluació Sanitàries de Catalunya) |
| Catalan-Balearic Society of Home Hospitalisation | SCBHD (Societat Catalano-Balear d’Hospitalització Domiciliària) |
| Catalan Society of Geriatrics and Gerontology | SCGiG (Societat Catalana de Geriatria i Gerontologia) |
| Chronic obstructive pulmonary disease | COPD |
| Complex chronic patient | PCC (for Catalan “pacient crònic complex) |
| Comprehensive geriatric assessment | CGA |
| Continuous and urgent territorial care | ACUT (for Catalan “Atenció continuada i urgent de base territorial”) |
| Continuous positive pressure on the airway | CPAP |
| Diagnosis-related group | DRG |
| Early supported discharge | ESD |
| Emergency Medical Service | EMS |
| Family and Community Nursing Association of Catalonia | AIFICC (Associació d’Infermeria Familiar i Comunitària de Catalunya) |
| Palliative care programme and support teams | PADES (for Catalan “ Programa d’atenció domiciliària i equips de suport”) |
| Home care programme at primary care | ATDOM (for Catalan “Programa d’atenció domiciliària de l’atenció primària”) |
| Hospital at Home | HaH |
| Outpatient hospital medication prescription | MHDA (for Catalan “Medicació hospitalària de dispensació ambulatòria”) |

| | |
|---|---|
| Information and communication technologies | ICT |
| International normalized ratio | INR |
| Major outpatient surgery | MOS |
| Minimum basic data set | CMBD (for Catalan “Conjunt mínim bàsic de dades”) |
| Modules for the monitoring of quality indicators | MSIQ (for Catalan “Mòduls per al seguiment d’indicadors de qualitat”) |
| Nasogastric probing | NGP |
| National emergency plan of Catalonia | PLANUC (for Catalan “Pla nacional d'urgències de Catalunya”) |
| Plan for integrated social and healthcare | PAISS (for Catalan “Pla d’atenció integrada social i sanitària”) |
| Primary and community care | APIC (for Catalan “Atenció primària i comunitària”) |
| Primary care medical record | ECAP (for Catalan “Estació clínica d'atenció primària”) |
| Primary care emergencies | CUAP (for Catalan “Centre d'Urgències Atenció Primària”) |
| Primary care team | EAP (for Catalan “Equip d’atenció primària”) |
| Rapid diagnostic unit | RDU |
| Shared healthcare record in Catalonia | HC3 |
| Spanish Society of Home Hospitalisation | SEHAD (Sociedad Española de Hospitalización a Domicilio) |

#### 3. Introduction

We live in a time of change, of increased complexity and needs of the healthcare system. The complexity of patients (with or without chronic pathology) throughout the entire process requires the creation or reorganization of the different services in open and dynamic models based on the best available evidence, offering the patients the service that best meets their needs with the least risks. Home hospitalisation (HaH) falls into this category of services.

The internationally accepted definition of HaH is a hospital resource that offers the patient, at home, medical and nursing treatment in the same quantity and quality as in the hospital. The services should be similar to those provided in a hospital setting. The aim is to attend to acute processes that require admission. If the service is unavailable, the patient must remain admitted to the hospital. This aspect clearly differentiates it from other community services. It is meant to treat patients with a limited time expectation of treatment and care. The service treats medical and surgical patients, with or without chronic pathology. The hospital maintains clinical, logistical, and legal responsibilities. The HaH episode has an admission and discharge report/log.

HaH is a real alternative to conventional hospitalisation because it is safe, sustainable, and satisfying for the patient as long as it fulfils its function and is not confused with other home care services; otherwise, it can be inefficient and/or redundant. When we talk about HaH, we cannot fail to mention the group led by Professor Bruce Left of the Johns Hopkins University School of Medicine for its great contribution both at scientific and organizational levels. Since the initial report by Left *et al*., which showed the feasibility and cost-effectiveness of HaH, the evaluation of the programs has historically shown significant complexities due to the heterogeneity of the study groups. Recent reviews by Cochrane appear to overcome these limitations.

In this context, HaH is an option for specialised, territorial-based care that offers complex, hospital-intensive care at home when the patient requires hospital admission. It supports all hospitals in the territory, the basic health areas (ABS in Catalan) and the residential centres. It takes on the complexity of the patient at home until discharge. HaH places great importance on continuity of care and offers patients the best facility to meet their needs. For these reasons, HaH is expected to be a care modality that improves the adequacy of complex, specialised care, reduces the size and promotes the future hospital's evolution and contributes to the system's sustainability.

The expansion of HaH should be done while ensuring the adequacy of a common care model according to the specific characteristics of each territory (population density, geographical characteristics, coverage of the different territorial resources, etc.). It should be kept in mind that the care results of a HaH unit are fundamentally related to two factors: the proper selection of patients to be treated in the HaH regime and the resolution capacity of the care team. Consequently, a HaH care network cannot be improvised without planning the foreseen activity and the necessary resources. A critical point lies in having motivated, properly trained and experienced teams with enough management support.

HaH can take care of a wide range of patient profiles with conditions that would require hospital admission. They include medical (serious infectious diseases, cancer, etc.) and post-surgical complex care processes, regardless of age. In any case, one of the profiles in which the benefit of this modality has been demonstrated the most is that of patients with comorbidity, mostly related to chronic conditions. Thus, these multidisciplinary HaH units can be established as the first-choice facility for all patients who meet the criteria for hospital admission, as is already the case in some territories.

### 4. Objectives

#### 4.1. Main objective

To define the model of HaH care in Catalonia as an alternative to conventional hospitalisation, with territorial suitability criteria.

#### 4.2. Secondary objectives

To define the portfolio of services considering the intensity and complexity.

To define the criteria for providing HaH care of the highest quality and safety, with the same guarantees as if the patient were in an acute care hospital.

To define an integrated territorial organization of HaH that provides a safe and efficient response, understood as an opportunity wherever it can be guaranteed.

To ensure safe and collaborative transitions from HaH to the other appropriate territorial care services at the time of discharge.

To ensure efficient coordination of pre-admission and post-admission to HaH between the different services and levels of care in the same territory or others.

To have common evaluation indicators that allow for quality assessment and public benchmarking between the HaH services of the different providers.

### 5. Characteristics of the care model

#### 5.1. Definition of the model

HaH is a care model in which hospital healthcare professionals provide active treatment to the patient at home for a condition that would otherwise require the patient’s admission to a healthcare centre.

Home hospitalisation is NOT:

- Low-complexity patient care.
- Chronic disease management.
- Palliative care.
- Follow-up by professionals from a single health discipline.
- Follow-up of patients who do not need hospital admission.

- Urgent home care.
- Own care or treatments in the primary and community care (APIC), according to the Catalan primary homecare program (ATDOM).
- Check-ups for major outpatient surgery (MOS) or post-surgical follow-up without any other complications.

##### 5.2. Patient inclusion/exclusion criteria

**Inclusion**

- Clinical criteria for the hospital admission.
- All ages.[4]
- Established diagnosis and absence of criteria for intensive monitoring.
- Patient home within the service area, always within a 30-40-minute radius.
- Existence of a capable primary caregiver, adequate for the patient's care needs. This criterion must be assessed individually in young patients or those with a high level of autonomy.
- Home with a phone or mobile phone and adequate hygienic conditions.
- Acceptance of the resource by the patient, caregiver, and/or family.[5] The acceptance of the service must be clearly explained in the admission or discharge report in the computer system.
- Acceptance by the HaH team.

**Exclusion**

- Non-compliance with a single inclusion criterion
- Severe behaviour disorders (agitation, violent behaviour).
- Risk of suicide.

##### 5.3. Efficiency and quality criteria

To help the healthcare planning of HaH and its efficiency and quality, key elements such as the reference population, the profile of the eligible patients (section 5.2. Patient inclusion/exclusion criteria), the portfolio of services, as well as the minimum requirements and resources for their implementation must be analysed. The aspects to be developed are:

**Population elements: population data must be analysed from different perspectives**

- According to the territorial units (minimum ABS and the groupings to be

[4] The paediatric HaH service line is a proposal for future work.
[5] Given the reality of women's naturalised feeling of obligation to care, it is necessary for professionals to consider that acceptance does not respond to social pressure, but that there is a clear willingness to assume this role. For this reason, an independent interview with the caregiver and/or family must be included to confirm their acceptance and avoid the imposed altruism.

considered).

- According to population density for the analysis of territorial dispersion.
- Estimate medium-term population projections (10 years) by sex and age groups (> 18 years).
- According to eligible candidates (clinical profile of the patient, sociodemographic elements, and others).
- According to isochrony and territorial conditions (a standard 30-minute radius is proposed, adaptable to a maximum of 40 minutes, according to local conditions, intensity and complexity of home visits, road communications and/or other specific elements).

##### Elements linked to the team of professionals

The team of professionals must be interdisciplinary, with sufficient capacity according to the therapeutic intensity required by the patient, that can incorporate the portfolio of services of a healthcare territory for its population. The team must have the ability to coordinate and clinically interact with the rest of the resources and levels of care (primary and community care, intermediate care, reference hospital care, and others) while maximizing an integrated system that collaborates in the same area of reference. The following proposals should be considered:

- Team made up of a minimum of 2 physicians and 4 full-time nurses (facilitating a flexible organization). The number of professionals depends on the number of patients and the level of intervention at home.
- A minimum of 2 HaH teams must be available in the area to provide coverage 7 days a week.
- Hourly coverage (for more details, see section 5.4. Coverage schedule and guarantee of continuous care).
- Consider the support of other professional profiles (occupational therapist, social worker, physiotherapist/rehabilitation therapist, etc.).
- Clear circuits and fast coordination with the acute care hospital of reference and the Emergency Medical Service (EMS).

##### Other elements to consider

- Ensure connectivity (availability of ICT tools) for on-site patient registration and monitoring and connectivity with the entire system (shared information) and other technological elements.
- The incorporation of professionals from other fields and services to the HaH team, according to the characteristics of each territory, always supervised by the latter and working functionally to obtain the same health results.
- Shared field management, especially with primary and community care, which facilitates the relationship and effective coordination between the different levels of care and the shared management of processes, adapting, if necessary, clinical practice guidelines or care pathways.
- Consider aspects of training and minimum experience of the HaH team members and promote research lines.
- Ensure territorial equity.

- Formulate assessment elements.

##### 5.4. Coverage schedule and guarantee of continuous care

HaH coverage must be 24 hours a day (24/7), 365 days a year. The teams of HaH professionals must ensure care for patients 24 hours a day and must guarantee home visits, if necessary, at any time.

Therefore, conceptually, HaH must guarantee the following:

- Continuous telephone response by a HaH member, 24 hours a day.
- Daytime presence of physician and nurse, 7 days a week (8 a.m. to 8 p.m.).
- HaH health professionals are reachable at night (8 p.m. to 8 a.m.) and, when necessary, available for home visits.

Each territory, considering the characteristics of its population and the relationship of HaH with the different health resources and services, can create strategic alliances, for example, with primary and community care services, to ensure 24/7 coverage, within a defined 30-to-40-minute radius, according to territorial conditions and road communications.

##### 5.5. Levels of care complexity

To define the different levels of care that HaH can provide, the following constitutive requirements must be considered, distinguishing it from other home care forms.

**Complexity.** Patients treated in this care modality must have acute health problems that require specialised knowledge for their care, as well as the participation of other areas of expertise and different professional categories.

**Intensity.** Patients cared for in HaH must require a frequency of care and interventions by HaH professionals that is typical of hospital care.

**Temporality**. Patients are treated in HaH for defined periods in relation to acute or re-exacerbated problems, for which they must be treated (usually days/weeks, considering the admission diagnosis and the evolution of the episode).

**Territoriality.** The scope of action of HaH must be that of the territory defined by the sum of the ABS (for example, the territorial health division AGA – care management area - of the Catalan Health Service, CatSalut).[6] In those territories in which there is no reference hospital with HaH, it can be foreseen that this service will be provided by the nearest hospital with HaH, on condition that this does not imply non-compliance with quality and safety criteria (such as isochrony and efficiency), and with the support of the existing resources in the territory.

[6] Territorial boundaries based on the grouping of basic health areas. This delimitation corresponds to criteria of operational planning, coordination, and analysis of the main flows between primary care and basic hospital aimed at promoting integrated management.

**Complementarity.** HaH does not invade other non-hospital intensity home-based care types, as HaH exclusively replaces a hospital admission (AA) and/or reduces the number of hospital days (ESD). Each of the different health resources has a defined role in the patient's home, and they collaborate to give the patient at all times the resources and health support they need (ATDOM, Primary Homecare Program and Support Teams [PADES], HaH). The coordinated and collaborative action of all of them allows for a safe, effective, and sustainable response, adapted to the needs of each case at the time of discharge, ensuring the best clinical management and safety for the patient in transitions.

Some of the benefits described in HaH over conventional hospitalisation are high patient satisfaction and the avoidance of adverse events associated with hospital stays. Additionally, this function represents for the territory the advantage of freeing up beds from healthcare centres, which contributes to dynamic patient flow. However, currently HaH admissions in Catalonia from any origin (hospital or territorial) are less than 3% of hospital discharges globally in Catalonia.[7]

To carry out its function, HaH can primarily act under two main care schemes or strategies depending on the patient's origin, depending on whether the patient is admitted to a hospital at the time of referral to HaH (ESD strategy) or needs admission directly to HaH (AA), in both cases to receive a hospital-intensity treatment.

**Early supported discharge (ESD) strategy**. It consists of continuing the hospital care for patients at their homes by shortening the planned stay in conventional hospitalisation for a specific diagnostic-related group (DRG). This modality effectively reduces the number of stays in conventional hospitalisation for patients already admitted, medical and/or surgical. The Catalan-Balearic Society of Home Hospitalisation (SCBHD) estimates that a proportion of 40% of HaH discharges should belong to this alternative, although from the Catalan Minimum Basic Data Set (CMBD)'s review of the HaH discharges in 2014 (Agency for Health Quality and Assessment of Catalonia - AQuAS report), it can be inferred that in the case of larger hospitals or with tertiary medical services, this proportion could be higher (50%).

**Admission avoidance (AA) strategy.** These are all those admissions to HaH that are made from a facility different from conventional hospitalisation (emergency department, APIC, PADES, day hospitals, etc.) to avoid admission in patients who require hospital care for an acute process. This care scheme provides directly saving in admissions. The SCBHD considers that the ideal percentage of HaH activity for AA would be about 60% of HaH discharges.

#### 5.6. Patient profile

##### Medical (AA/ESD)

- Acute and high complexity pathology (infectious and non-infectious),

[7] Alepuz L, Antón F, Arias J, Espallargues M, Estrada MD, Estrada O, Hermida L, Fernández M, Massa B, Mirón M, Murja A, Muñoz L, Ponce MA, Rio M, Torres A Hospitalització domiciliària. Barcelona: Agència de Qualitat i Avaluació Sanitàries de Catalunya. Departament de Salut. Generalitat de Catalunya; 2018.

medical or surgical when surgery is ruled out, and conservative treatment (for example, diverticulitis) is chosen.
- Acute exacerbation of chronic pathology.

Most prevalent diagnoses:

- Acute exacerbation of chronic pathology (COPD, heart failure, asthma, bronchiectasis, etc.).
- Acute pathology:
  - Infectious pathology (flu, pneumonia, pyelonephritis, cellulitis, COVID-19, etc.).
  - Non-infectious pathology (deep vein thrombosis, pulmonary thromboembolism, etc.).
- Complex chronic patients or frail elderly patients admitted to acute hospitals, affected by acute disabling processes (e.g., stroke, etc.), as an alternative to prolonging the admission to acute hospitals or admission to an intermediate care hospital (only in case of AA).

**Surgical (AA/ESD)**

- Simple post-surgery as early discharge to reduce the days of hospitalisation and complete this admission at home.
- Post-surgical, for complications related to the surgical process, either due to the underlying comorbidity and/or postoperative complications: infections, seromas, dehiscence, fistulas, etc.
- Pre-surgical patients with a need for complex hospital treatment prior to surgery to decrease pre-surgical stays.
- Complex chronic patients or frail elderly patients admitted to the acute hospital, affected by acute disabling surgical processes (orthogeriatric processes, etc.), as an alternative to continuing in the acute hospital or being transferred to the intermediate care hospital.

In either of the two patient profiles (surgical and medical), patients with infectious diseases (methicillin-resistant staphylococcus aureus [MRSA], COVID, extended-spectrum beta-lactamases [ESBLs], etc.) are expected to be admitted to HaH. The infection control measures at home that must be extended to the family or caregivers before admission to HaH are relevant to adopting the appropriate protection and management measures.

#### 5.7. Patient origin

##### Admission avoidance

This type of admission to HaH can be from a different care facility than conventional hospitalisation for patients who require hospital-level care for an acute process. This care model directly provides for total admission avoidance to the hospital.

Types of admission:

- Hospital-based facilities (except hospitalisation areas): hospital emergencies, outpatient clinics, day hospitals and rapid diagnostic units (RDUs).
- Community-based facilities: preferably primary care team (EAP), primary care emergencies (CUAP) or continuous and urgent territorial care (ACUT), primary and community care (APIC), as well as PADES, nursing home support teams, etc.

**Early supported discharge from a hospital ward**

This type of HaH admission is from a conventional hospitalisation facility for patients who require hospital care intensity for an acute process. This care scheme provides for shortening the days of hospital admission.

Admission routes:

- Hospital medical and surgical services (except for hospital emergencies, outpatient clinics, day hospitals and RDU).

#### 5.8. Time response to HaH admission request

The measurement of the response time (less than 24/48 hours) is the difference between the time of request for admission to HaH and the actual admission to HaH. Since the general response time for admission from the emergency room to the hospital ward uses the time of arrival to the emergency room and not the time of formal request for admission, the same measurement should be used as an indicator to compare results with conventional hospitalisation.

In the case of visits from emergency and territorial services, the HaH response must be ≤ 24 hours, provided that, when the consult to the HaH is formalized, the consulting department has checked the criteria for inclusion/exclusion to/from the HaH.

In the case of visits from the hospital ward, a response time of fewer than 48 hours is considered ideal, provided that, when the consult with the HaH is formalized and the consulting department has checked the criteria for inclusion/exclusion to/from the HaH.

#### 5.9. Equipment and resources

In the field of diagnosis and treatment, the HaH unit has the same resources as the hospital. If a technique cannot be carried out at home (complementary examinations, diagnostic techniques or treatments), the patient is transferred to the hospital promptly for its realization.

**Diagnostic resources**

- Electrocardiography
- Forced spirometry
- Paracentesis
- Thoracentesis
- Point-of-care and continuous overnight pulse oximetry
- Cooximetry
- AED (mask, resuscitation bag)
- Ultrasound
- Collection of samples, tests, arterial blood gas test

**Therapeutic resources**

- Oxygen therapy
- Paracentesis
- Thoracentesis
- Aerosol therapy (bronchodilator and antibiotic)
- Complex wound care
- Intravenous treatments. Perfusion pumps, elastomeric pumps
- Blood components transfusions (Annex 2)
- Intravenous iron therapy (Annex 3)
- Ostomy and tracheostomy care and management
- Home mechanical ventilation, BIPAP and CPAP
- Fingertip puncture for INR control
- Enteral or invasive nutrition (surgical gastrostomy, percutaneous endoscopy [PEG], jejunostomy)
- Parenteral nutrition management
- Surgical drains
- Pleural drains
- Bladder catheterization
- Tube and catheter replacement
- Treatments with negative pressure therapy for complex wounds
- Paracentesis
- Basic and advanced life support

##### 5.10. Transitional models

All citizens have the right to receive the highest quality and safest care in the most appropriate place according to their needs at a given time and provided by the best professionals. The coordination and integration of healthcare are compromised by complex challenges related to transitions between levels of care, a high prevalence of chronic diseases and high complexity in patients with or without chronic pathology. A single patient may need different intensities of care and attention, so different healthcare resources and/or intermediate care can be used to give them the best response regarding quality and safety. To provide all the safety to this resource integration, it is essential that the step from one resource to another (the transition) be made with the best coordination and collaboration between them. Transitions, understood in a bidirectional way between hospital and community teams, affect all patients treated. The typology of the patient with or without chronic pathology makes it necessary, at different times of their process, to move through the health system (transitions) and/or need important changes in their treatment (for example, starting home dialysis or oxygen therapy, etc.).

Transitions are inevitable, but it should be borne in mind that they always represent an added risk to the patient, so for their safety, it is necessary to avoid unnecessary transitions. It is known that transitions are not always optimal and that the factors involved are multiple. Ineffective transitions negatively impact the patient, lead to a delay in the process, increase the risk of adverse events and an increase in stays, unplanned immediate readmissions, visits to emergency services and overall costs. Unplanned re-entry is an indicator that the transfer has not been optimal and that the coordination and the type of intervention must improve. Different initiatives have been established to improve transitions, especially on the patient, the process, and the results. These are the so-called transitional models.

Improving transitions is a complicated and multifactorial process. There is no single problem and no single solution. Aspects such as quality, efficiency, safety, comfort and the opinion of patients and family should be the central points for promoting change. To improve transitions, it is necessary to identify everything that does not add value, everything we do that poses a risk of human error, and everything we do that has high variability.

The different actions that are proposed must be carried out with the maximum possible consensus, with the best available evidence, that benefits the patient and facilitates the work of the professionals, but have an impact on the system, can be evaluated continuously and in line with the policies of the hospital and the region. The measures are more effective when the structure or care process is based on strong scientific evidence linked to good outcomes.

HaH plays an important role in transitions by providing the patient with the best service to meet their needs at their discharge from HaH. The approach to transition is made as early as possible in the care process, as it requires careful planning. Additionally, a bidirectional circuit is established: on the one hand, HaH professionals support community healthcare teams with APIC leadership to ensure transitions, and on the other hand, community healthcare teams, following HaH criteria, identify patients early on as candidates for HaH and coordinate with HaH teams at the time of admission to HaH.

<u>Carrying out safe transitions implies for HaH professionals</u>:

- Having up-to-date patient information at the time of admission to HaH (hospital staff, community teams) to confirm the suitability of the resource.
- Making an overall assessment of the patient during admission to HaH and contacting the professional teams that best meet the patients’

needs at discharge time.

- In groups of highly complex patients at home, offering the possibility of a joint home visit with the teams that will monitor them in the post-discharge period.
- Reflecting the type of follow-up and contacts in the discharge report of the shared healthcare record in Catalonia (HC3).

<u>For consulting professionals, from the hospital and the community, making safe transitions implies:</u>

- Facilitating that all patients with admission criteria can benefit from the alternative to conventional hospitalisation (HaH).
- Contacting the HaH teams (consultation).
- Providing updated patient information necessary for HaH to establish an individual work plan with the utmost safety for the patient.
- Working jointly with the HaH teams for safe transitions. HaH should collaborate in the transition planning, but under no circumstances should it delay discharge due to possible delays in managing the schedules of other resources.

#### 5.11. Professionals

##### Functions of professionals

As described in the previous sections, patients admitted to HaH may have a wide range of pathologies that, combined with the individual profile of the patient, condition very different HaH intervention needs, from intensive medical-surgical treatment with intravenous therapies and/or high complexity care (these interventions should be the majority in HaH) to care and follow-up of intensity corresponding to admission avoidance to an intermediate complexity hospital.

The current situation with an increasing number of patients with complex treatments and/or of advanced age, with fragility, multiple chronic conditions, complex chronic patients (PCC) and patients with advanced chronic disease (MACA), susceptible to suffering an increase in complications during hospital admissions, leads us to the fact that the health system must rethink strategies to address this situation by improving care both in terms of healthcare delivery model (person-centred care), efficiency and complexity management.

In recent decades, care provided by multidisciplinary teams using comprehensive geriatric assessment (CGA) in the elderly[8] at different levels of care, from APIC to specialised care, has shown benefits in mortality, functional decline, and cognitive impairment in the elderly population at risk.[9]

[8] See Annex 7. Comprehensive geriatric assessment (CGAI).
[9] Pilotto A, Cella A, Pilotto A, Daragjati J, Veronese N, Musacchio C et al. Three Decades of

Crisis management in elderly patients at home by HaH units that use CGA has also shown favourable results in recent years. The current scientific literature indicates that multidisciplinary interventions incorporating professionals with profiles specialised in geriatrics reduce complications during admissions such as delirium and functional decline, improve the degree of satisfaction of caregivers and have an impact on costs for the system[10]

HaH staff must have specific regulated training in basic life support (BLS) and the use of automatic external defibrillators (AED). Centres with HaH teams must ensure the accreditation of all HaH staff in BLS + AED and/or ALS and equip HaH vehicles with first-response material for cardiac arrest, including AED equipment. This training, equipment provision, and maintenance must be integrated into the strategy of the intrahospital cardiopulmonary resuscitation (CPR) commission.

It should be considered that the organizational model of HaH progressively incorporates sufficient training to attend: 1) patients with complex treatments at home; 2) neonatology and paediatric; 3) fragile and complex patients with acute and subacute health crises. Within this context, multidisciplinary work and CGA play a key role in promoting individualized care adapted to complex needs, as well as support and health education to patients, families, and caregivers, offering a versatile vision with the possibility of working on aspects of rehabilitation, both in terms of prevention and treatment and facilitating the continuity of post-crisis care.[11]

On the other hand, complex care for patients with mental health and addiction at home (which would be equivalent to mental health HaH) is another future area of work as a care model to be developed and integrated within the territorial health resources.

##### Professional support profiles

- Advanced practice nurse.
- Nurse with geriatric expertise or specialist.
- Occupational therapy.
- Social work.
- Speech therapy.
- Physiotherapy.
- Rehabilitation.
- Clinical psychology (linked to the mental health network).

---

Comprehensive Geriatric Assessment: Evidence Coming From Different Healthcare Settings and Specific Clinical Conditions. J Am Med Dir Assoc 2017;18(2):192.e1-192.e11.

[10] Mas MÀ, Santaeugènia S. Hospitalización domiciliaria en el paciente anciano: revisión de la evidencia y oportunidades de la geriatría. Rev Esp Geriatr Gerontol 2015;50(1):26-34

[11] Mas MÀ, Santaeugènia SJ, Tarazona-Santabalbina FJ, Gámez S, Inzitari M. Effectiveness of a Hospital-at-Home Integrated Care Program as Alternative Resource for Medical Crises Care in Older Adults With Complex Chronic Conditions. J Am Med Dir Assoc 2018;19(10):860-863.

- Geriatrician.
- Nutritionist.
- Hospital specialists (pulmonologists, oncologists, psychiatrists, surgeons, etc.).

**Professional training**

HaH requires specific knowledge and skills. That is why it should be integrated into the official healthcare training programs (MIR/EIR in Spain), not only because it represents a possible future line of work but also because knowledge of this area allows health professionals to promote HaH at the highest level and with the maximum guarantees of quality and safety.

The key element for developing this care model is the knowledge of all the healthcare professionals of the system and managers of the resolving capacity of HaH for acute health problems that would otherwise require admission to a hospital.

At the same time, it is necessary to develop teaching objectives that lead to a specific curricular itinerary, such as:

- Incorporation into teaching programs and MIR/EIR rotation.
- Incorporation into the curricular itinerary of training and teaching such as undergraduate, postgraduate or master's degree.

#### 6. Evaluation/indicators

When we need to evaluate, qualitatively and quantitatively, the changes in the health status of the population, to know the current health situation and to be able to apply appropriate planning in health policies, it is necessary to use statistical parameters, which in this case are indicators, obtained from rigorous, stable, and reliable sources of information.

In HaH, all those indicators that are of interest must be incorporated, divided into the following information areas: 1) structure; 2) process; 3) outcomes, the latter also in subareas of information (general, mortality, safety, satisfaction, re-entry at 30 days, specific to the HaH provider and territorial coordination), and 4) evaluation in the transfusion or administration of blood components in HaH. These are described in the following sections.

For its sensitivity, level of results and evolution, the Catalan Health Service has selected the highest priority and key indicators for its annual monitoring and the possibility of assessing its continuation. These indicators are shared and published by CatSalut, and the list of these indicators is detailed in Annex 8, HaH indicator cards.

##### 6.1. Structure indicators

- Defined territorial area.

- % of the defined reference population that can be admitted to HaH.
- Number of HaH beds.
- Recognition/accreditation of the unit.
- Profile and number of professionals assigned to HaH equivalent to full-time equivalents.
- Infrastructure. HaH installations linked to a hospital facility.
- Teams of hospital professionals from HaH, to ensure patient's care during admission to HaH, 24 h x 365 days/year. HaH call reception circuit 24 h x 365 days/year.
- Personal and specific equipment (diagnostic and therapeutic) for attending patients during HaH admission, adapted to the type of patients treated in each territory, in accordance with the minimum requirements of the diagnostic and therapeutic resources section of point 5.9. Equipment and resources.
- Information systems that guarantee connectivity from home and with the hospital, to access and record throughout electronic information systems (medical records, HC3, etc.).

#### 6.2. Process indicators

- Collaborative work: the existence of one or more protocols agreed upon the community and hospital APIC teams (YES/NO).
- Average number of visits/patients in one year by the HaH team.
- Number of patients with HaH.
- Number of contacts with HaH.
- Number of patients and the number of HaH contacts per hospital.
- Number and type of procedures during HaH (according to the discharge report record).
- Requirements for safe transitions after HaH discharge (that meet the criteria in the transition section) (YES/NO).
- % of conventional hospital admissions as HaH discharge during HaH hospitalization
- HaH modalities per hospital, avoidance of admission and early discharge.
- Yearly number of HaH admissions per patient.
- % of visits to emergencies (without admission) during HaH.
- % of admissions with response time < 24 h/48 h (waiting time from the moment the patient enters the emergency room and is admitted to HaH).
- % of origin of admission to HaH.
- % of destination of admission to HAH.
- Occupancy rate.
- Total stays.
- Average and median stay.
- % of hospital HaH discharges compared to conventional discharges from the same hospital.
- % of HaH discharges from the hospital compared to the sum of conventional discharges from the territory.

- % of HaH discharges from the territory compared to conventional registrations according to population.
- Destination upon discharge (home, exitus, intermediate care centre, etc.).
- Patient profile: age groups, sex.
- Main diagnosis.
- Secondary diagnoses.
- Procedures during HaH.
- Participation in research groups.

##### 6.3. Outcome indicators

- **General**
    - Complexity according to the diagnosis-related group (DRG).
    - Stay adjusted to DRG, in case of AA.
    - Comparison of HaH stays versus conventional stays for the same pathology.
    - Number of readmissions after 30 days for the same diagnosis.
    - Visits to hospital emergencies for the same diagnosis.
    - Costs of drugs consumed by HaH patients.

- **Mortality**
    - Mortality rate during the HaH episode.
    - DRG-adjusted mortality.

- **Safety**
    - Catheter-related sepsis rate.
    - Pressure ulcer rate.
    - Fall rate.
    - Cardiorespiratory arrest rate (unexpected death that requires manoeuvres) or requirement of basic life support manoeuvres.
    - Rate of severe adverse drug reactions.
    - Rate of delirium episodes.

- **Satisfaction**
    - Patient satisfaction (information source, biennal: PLAENSA [Catalan plan of satisfaction questionnaires])
    - Patient/caregiver experience (patient's reported experience measurements, PREM).

- **Readmissions of HaH patients within 30 days after discharge**

  These indicators provide information on the quality of HaH care for patients when analysed for specific, never general, diagnoses. In addition, the analysis of readmissions within 30 days for a diagnosis (e.g., heart failure) must always include the patient's context (age, functional situation, the severity of the underlying pathology, recent prior admissions, etc.) and that of the response of the territorial resources after HaH discharge.

Therefore, when evaluating the outcome indicators within 30 days after discharge, the following information is needed:

- Type of intervention and the number of contacts per patient from the destination resource upon HaH discharge (visits, calls).
- Time elapsed between HaH discharge and the first in the ECAP recorded in-person visit at the patient’s home by the responsible territorial team.

- **Specific to the HaH provider**
  - Type of intervention and a number of contacts per patient by community teams (visits, calls).
  - Identification of the team responsible for follow-up.
  - % of joint HaH visits and the most relevant or appropriate resource for HaH patient discharge.
  - % of unplanned conventional hospital admissions (define the HAH patients who must re-enter HaH urgently) and collected by each HaH.
  - Negatives per HaH admitted patient (exclusion reason) specific to each HaH.
  - Negatives per HaH admitted caregiver (exclusion reason) specific to each HaH.
  - Lack of caregivers (exclusion reason) specific to each HaH.
  - Number of visits to HaH.
  - Number of accepted visits.

- **Territorial coordination (transitions)**
  - Number of HaH coordinations at discharge with APIC (preparation of discharge programme [PREALT], contact with case managers, etc.).
  - Number of admissions to HaH at the request of community facilities: APIC, PADES, etc., always under the supervision of the APIC.

##### 6.4. Blood transfusion or administration evaluation

- Number of blood transfusions carried out at home in patients admitted to HaH.
- Number of medical records containing transfusion data (the type of component, identification number, start and end time of the transfusion, and nursing professional responsible for the administration).
- Number of errors or incidents related to the request and non-correspondence between the request data and the sample.
- Number of correctly filled-out transfusion control sheets (calculation carried out by sampling).
- Number of blood components returned to the Blood Bank that has to be rejected for exceeding the time allowed outside thermal control.
- Case analysis, if applicable.

#### 7. Medication dispensation

The prescription of hospital medication is done through medical orders according to the corresponding hospital model and using the necessary information system to guarantee the recording and traceability of the prescription and the correct dispensation by the responsible hospital pharmacy service.

The hospital must guarantee the dispensation of the necessary hospital medication for the active episode treatment, which should be dispensed by the corresponding hospital pharmacy in unit dose format, as well as provide all medication on prescription (non-hospital) that the patient had prescribed electronically prior to the episode and needs to be continued during home hospitalisation.

Also, when applicable, the hospital pharmacy responsible for the care of the episode should provide the outpatient dispensing hospital medication (MHDA) that may be necessary during the home hospitalisation period.

#### 8. Future plans

To continue the implementation of HaH as a care modality that aims to transform healthcare, new approaches will be added and included in the future.

- Promoting networking among different providers.
- Healthcare coverage with safety and efficiency guarantees.
- Reported Outcome Measures.
- HaH in neonatology and paediatrics.
- HaH in mental health and addictions.
- Coordination between HaH and medical emergencies.
- Establishing coordination and cooperation mechanisms for territorial social work for integrated care focused on the needs of the patient and their family.
- Assessing and developing PREMs (Patient Reported Experience Measures) and PROMs (Patient Reported Outcome Measures).
- Adapting the information of CatSalut's information systems (MSIQ, CMBD).
- Automation of exploitation of reports and feedback to providers.
- Information systems that provide return information to the HaH team, bi-directional information and ECAP as a starting point.
- Possibility of negotiating night-time coverage among several HaH at a territorial level.
- Achieving a 5% of hospital discharges that must be via direct home hospitalisation.
- Ensuring that the nurses develop their nursing and leadership skills to the fullest in HaH.
- Developing a cross-sectional and joint approach between home care (ATDOM) and HaH.

#### 9. Annexes

##### Annex 1. Information for the HaH information leaflet

*This is a proposal for basic information collected in this document that is useful for information leaflets prepared by SISCAT providers.*[12]

**What is it?**

Home hospitalisation (HaH) is a type of care in which hospital professionals provide active treatment to the patient at their home for a condition that would otherwise require acute or intermediate hospital care.

**Coverage and schedule**

The teams of professionals must ensure care for the patient 24 hours a day, 365 days a year. The team must also provide all necessary materials and medication for the patient during their admission at home, except for the chronic medication (on electronic prescription).

During the day, 7 days a week, **(8 a.m. to 8 p.m.)** **there will be a presence of a doctor and a nurse**. **During the night** (**8 p.m. to 8 a.m.)** HaH health professionals are reachable and, when necessary, available for home visits. We will always call you before going to your home.

**HaH activation time**

For those visits made through emergency units, the HaH response time must be ≤ 24 hours.

For visits made from the hospital (excluding those made from emergencies), the ideal response time is less than 48 hours.

If you have any doubts or emergencies, you can call the phone number of your HaH, which has been provided to you, or the emergency number 061. When you call these phone numbers, please explain that you are admitted to the HaH you have been assigned.

**Equipment and resources**

In terms of diagnosis and treatment, the unit has the same resources as a hospital. If a technique cannot be carried out at home (complementary exams, diagnostic techniques or treatments), the patient must be transferred to the hospital on a case-to-case basis to perform them.

**Hospital at Home offers patients the best service to respond to their needs, as well as all the necessary hospital care at home.**

[12] *It is the initiative and responsibility of each provider to make and print these brochures.*

#### Annex 2. Blood component transfusion at home for HaH

Action procedure for cases of HaH. Description of blood component transfusion at home. The sequence of actions to administer a blood component (red blood cell concentrate, platelet concentrate or plasma) to a patient.

##### Introduction

The increase in life expectancy leads to a proportionally larger number of people who require a transfusion at some point. Although blood component transfusions traditionally take place in the hospital setting, to improve care in terms of accessibility and convenience for patients with chronic pathology, blood components can occasionally be administered outside the hospital setting. In fact, transfusion of blood components at home is a growing practice that will likely increase in the coming years, with similar indications as those given in the hospital setting. This transfusion option would be appropriate for patients with chronic pathology associated with anaemia or thrombocytopenia.

The risks of home transfusion are essentially the same as those of traditional transfusion, but its outpatient nature requires maximizing safety measures and the implementation of complementary requirements to guarantee the safety of the patient. The most important factor that differentiates the administration of blood components at home or on an outpatient basis from doing so in a hospital setting is the limited availability of emergency medical care in case of serious complications. This makes it crucial to strictly and adequately select patients who are candidates, train and prepare the home transfusion team, and have procedures that cover all aspects of the transfusion process outside of the hospital.

It highlights the proper and rigorous selection of candidates for this transfusion modality, a trained and qualified home transfusion team, and solid and validated work procedures.

##### Objective

The objective of this document is to consolidate an action procedure for the transfusion of blood components in HaH patients via the SISCAT system. Specifically, it aims to:

- Ensure the correct handling and administration of blood components.
- Ensure patient safety throughout the treatment.
- Prevent and/or detect at an early stage the appearance of potential problems related to treatment.
- Know the appropriate action in case of an adverse reaction or incident.
- Provide guidelines for the safety of the healthcare professional performing the task.

**Inclusion and exclusion criteria for a patient to receive a blood component transfusion at home within the framework of HaH**

Inclusion criteria:

- Meeting the general criteria for admission to home hospitalisation (HaH) (see section 5.2. Inclusion/exclusion criteria, of the HaH model).
- Clinical stability despite symptomatic anaemia with the transfusion indication according to hospital protocol.
- Patient with no history of transfusion reactions (transfusion-associated acute lung injury [TRALI], febrile non-haemolytic reaction or allergy).
- Conscious and oriented patients who can give their informed consent for the transfusion, as well as report any symptoms derived from a possible reaction. If the patient cannot give consent, it must be given by the family or legal guardian.
- The location of the home must allow for access so that the patient can be attended by emergency personnel in case of severe complications within a maximum of 30-40 minutes.

Exclusion criteria:

A person cannot be accepted as a candidate for blood transfusion at HaH if one or more of the following circumstances occur:

- The patient or legal guardian (family or guardian) refuses to give informed consent for blood transfusion.
- The patient is unable to provide the necessary information for proper identification.
- Absence of a HaH physician connected with the home transfusion team.
- Absence of an adult at home during the procedure and the hours after the procedure, to monitor and assist the patient in the hours after the transfusion. The companion must be instructed on how to act in case of a reaction, especially on contacting the reference hospital's corresponding department.
- Alteration in cardiac or lung function, or any other process that puts the patient's clinical stability at risk. In these cases, the referral to a hospital to receive the transfusion should be considered.
- If the patient is a carrier of an autoantibody or alloantibody, and strictly compatible red blood cell concentrates are unavailable. In this case, the transfusion should be performed in a hospital under strict monitoring and with the possibility of quick treatment, in case of a possible reaction.
- History of transfusion complications or repeat reactions.

**Blood components and other necessary materials for home transfusion**

The most commonly transfused blood component at home is red blood cell concentrate, but platelet or plasma transfusions are also planned.

It is recommended that no more than two erythrocytes or plasma concentrates be transfused in each transfusion episode, and in the case of platelets, no more than a mixture of 4-5 units or thrombocytapheresis.

For each home transfusion, the following materials must be used:

- Blood component in perfect condition (intact bag and no visible anomalies, such as air bubbles or clots, among others, which would indicate possible bacterial growth) and labelled with the patient's name.
- Refrigerated bag or fridge validated for the transport of hemoderivatives.
- Blood grouping reagents.
- Standard transfusion equipment that incorporates a drip chamber with a 170–260-micron filter provided by the Blood Bank.
- Infusion pump suitable for hemoderivatives and equipment for the specific pump that allows us to control the infusion speed.
- 50 cc of physiological serum.
- Disposable gloves.
- 20 mg Furosemide vials for pre-transfusion/post-transfusion administration.
- Transfusion equipment in case of transfusion reaction, which must contain:
  - 2 vials of adrenaline.
  - 1 vial of Urbason (methylprednisolone) 40 mg.
  - 2 vials of Polaramine (dexclorfeniramina) 2 mg.
  - 1 vial of Paracetamol 1g.
  - 2 vials of Hidrocortisona 100 mg.
- Documentation: control sheet.

**<u>Healthcare team responsible for the blood transfusion procedure at home</u>**

The staff responsible for the process, from the extraction of the blood sample, and transfusion compatibility to the administration of the component, must be trained and qualified to perform the transfusion with the highest proficiency. They must have the following knowledge:

- Thorough knowledge of procedures related to the safe administration of blood components.
- Thorough knowledge of the risks of blood transfusion (immediate and delayed reactions), as well as the protocol for dealing with each type of transfusion reaction, including pharmacological treatment of shock.
- Knowledge of the procedure to be used and collected within the framework of the blood transfusion procedure, as well as the necessary steps to record it and incorporate it into the appropriate information systems.

Each of the home transfusion team members must have a defined and clear role. The competences of this staff must be reviewed periodically. If the procedure is carried out exclusively by nursing staff, there is a medical professional located to resolve any doubts, give instructions for action in the event of a transfusion reaction, and, if necessary, quickly go to the home.

**Procedure for home blood transfusion**

The team that performs a blood transfusion at home within the HaH context must have, be familiar with and follow the validated procedures:

- Procedure for blood sample extraction.
- Procedure for obtaining and recording informed consent.
- Procedure for safely administering blood and/or blood components (decline and perfusion pump).
- Procedure for the collection of the blood components from the responsible transfusion service.
- Procedure for the safe transport of components.
- Procedure for returning the bag and any other elements used in the transfusion that is susceptible to rejection.
- Procedures for the registration of each transfused component, its recipient and any observations that are deemed appropriate in the event of a possible transfusion reaction or incident to be highlighted.
- Procedure to deal with each of the acute transfusion reactions.

**Records:**

- Transfusion request.
- Evolution sheet or unit-specific ones (including, for example, number of transfused units, bag identifier number, date, time of the start of transfusion, duration, observations, and incidents).
- Transfusion control sheet.
- Transfusion reaction notification (if applicable).
- Clinical variables sheet.
- Work accident record.

#### Annex 3. Recommendations for the administration of intravenous iron therapy at home

HaH healthcare personnel are exposed to the probability of witnessing critical clinical situations, including cardiac arrest, due to the increasing complexity and chronicity of patients treated at home. At the same time, interventions that are performed, such as administering antibiotics, intravenous iron, blood product transfusions or other medications can trigger serious anaphylactic reactions.

It is, therefore, necessary that HaH staff have specific regulated training in basic life support (BLS) and the use of automatic external defibrillators (AEDs), and that the mobile units are equipped with the first aid material for cardiac arrest (mask, bag-valve mask and AED device) to be able to start the manoeuvres without delay while alerting the emergency medical services to activate the advanced life support (ALS). This requirement of HaH units constitutes an additional safety element for patients, and, in addition, it supports society as it increases the availability of initial care for cardiac arrest in public areas through the same trained volunteer citizens, schools, and security forces.

Centres with HaH teams must ensure the accreditation of all their HaH professionals in BLS + DEA and/or ALS and provide HaH vehicles with first intervention material in cardiac arrest, including DEA equipment. This training, provision and maintenance of the equipment must be integrated with the intrahospital cardiopulmonary resuscitation commission (CPR) strategy.

Related links:

*Reference of the Spanish Agency of Medicines and Health Products.*
*https://www.aemps.gob.es/informa/notasInformativas/medicamentosUsoHumano/seguridad/2013/docs/NI-MUH_FV_20-2013-hierro_intravenoso.pdf*
*Technical data sheets of administration recommendations.*
***Venofer:*** *https://cima.aemps.es/cima/dochtml/ft/64000/FichaTecnica_64000.html*
***Ferinject:*** *https://cima.aemps.es/cima/dochtml/ft/69771/FT_69771.html*

#### Annex 4. ICD-10-CM/PCS coding for HaH

Recommendations for the use of ICD-10-CM/PCS.

Since 2018, the new **ICD-10-CM/PCS** classification has already been generally used. This classification incorporates a large number of new codes, both for diagnoses and procedures (therapeutic and diagnostic).

The ICD-10-CM/PCS is the new standard for coding clinical-healthcare data (morbidity and procedures) of the activity carried out in the health network of Catalonia and is part of the Catalogue of diagnoses and procedures of the CatSalut Systems Plan.

Regarding **diseases** (going from 16,019 to 94,444 codes), information relevant to outpatient care is added, injury codes are expanded, combinations of symptom and diagnostic codes are created to reduce the number of codes required to fully describe an impairment, a sixth and seventh character are added, and laterality is incorporated into the relevant codes. On the other hand, the new structure allows for future expansion of the classification.

As for the **procedures** (going from 4,646 to 79,758), the code structure is completely changed, and all codes have seven characters; the codes are built from values using algorithms made up of several tables. It is an exhaustive classification (each procedure has its code), expandable (the structure allows for the inclusion of new codes), multiaxial (the position of each digit has its own meaning according to the section), and it incorporates standardized terminology (precise definitions, each term has a specific meaning).

Related links:

*Official website of the Catalan Health Service.*
https://catsalut.gencat.cat/ca/proveidors-professionals/registres-catalegs/catalegs/diagnostics-procediments/suport-cim-10-mc-scp/

*The website of the Catalan Health Service (Servei Català de la Salut) provides information on the use of eCIEmaps, which is a tool provided by the Spanish Agency of Medicines and Medical Devices to search for specific codes. It shows the original classification, the desired destination classification, and the standardized description of the code.*
https://eciemaps.mscbs.gob.es/ecieMaps/browser/indexMapping.html#code=&source=cie9mc&target=cie10mc

*The World Health Organization (WHO) website provides information on the International Classification of Diseases (ICD-10), which was approved in May 1990 by the 43rd World Health Assembly. It is cited in over 20,000 scientific articles and is used by more than 100 countries worldwide.*
International Classification of Diseases (ICD)

**Annex 5. The HaH care process**

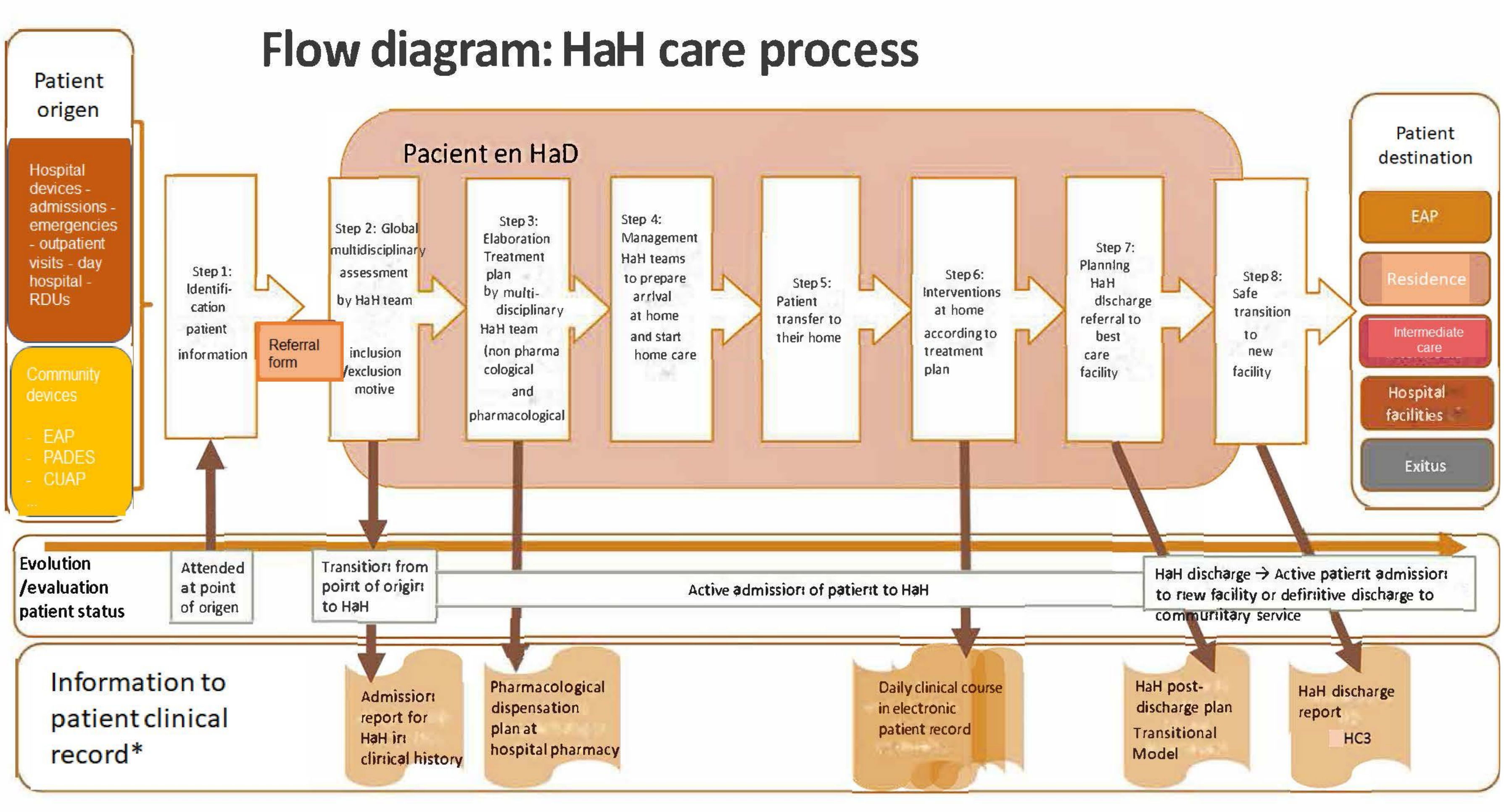

*Only included in electronic patient record when indicated

##### Annex 6. Electronic patient information flow

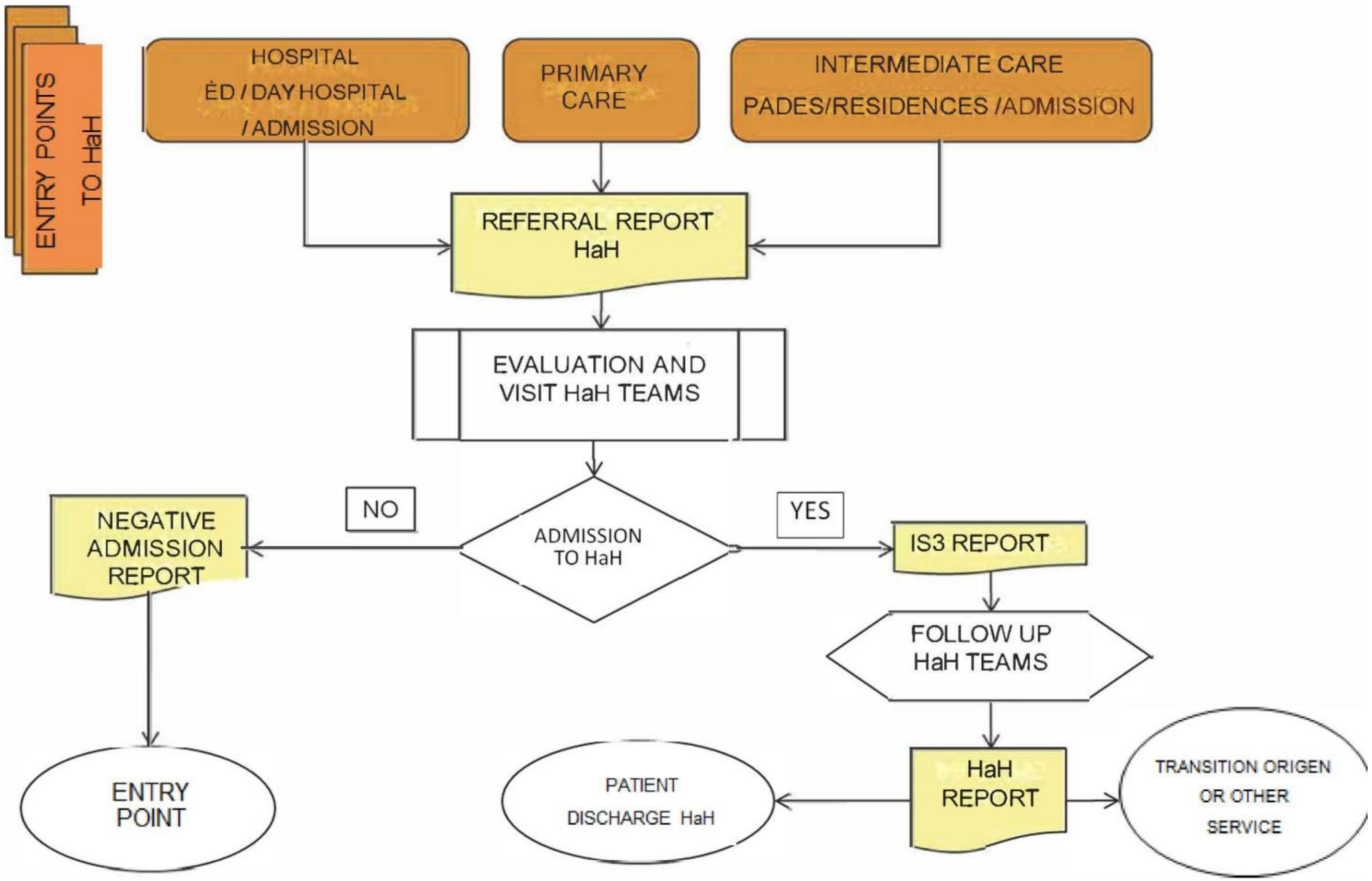

#### Annex 7. Comprehensive geriatric assessment (CGA)

In elderly patients who are admitted to HaH, CGA has shown multiple benefits: 1) a holistic view of the patient and their needs during admission while promoting individualized and adapted care for complex needs; 2) reduction of complications during admissions, such as delirium, functional decline or other geriatric syndromes; 3) improvement of health outcomes at discharge; 4) decrease in direct costs compared to conventional hospitalisation; 5) reduction of length of stay and readmissions; 6) therapeutic education for patients, families and caregivers and facilitating the continuity of post-crisis care.

##### Objectives of the assessment/comprehensive geriatric assessment

The first objective of the multidimensional assessment/CGA is to help professionals approach the situational diagnosis of people. In other words: 1) to assess the degree of reserve or frailty in people; 2) to identify which deficits or dimensions are affected (clinical, functional, emotional, cognitive, social, etc.), and 3) to identify people's needs (Figure 1). In this diagnostic process, it is important to consider both the situation at a moment in time (the "photograph" of the situation; the degree of severity) and its dynamic evolution (the "film" or progression criteria).

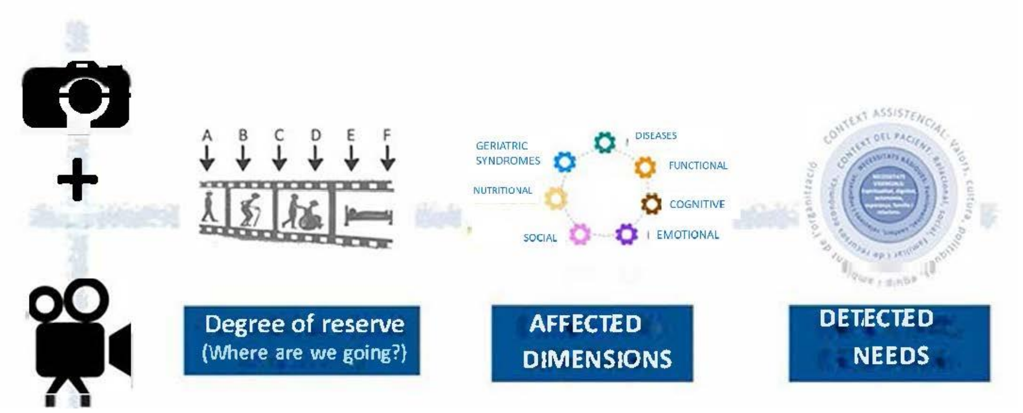

Figure 1. Key elements of the situational diagnosis. *Source: own elaboration.*

Accordingly, the CGA is the keystone for the individualization of interventions in elderly, frail or complex care needs (PCC) and advanced situation (MACA), as it: 1) facilitates the adequacy of the therapeutic intensity/harmonization of the care goals with the person's situation and their values and preferences; and 2) identifies the dimensions in which intervention is required, as well as the needs to be addressed (Figure 2).

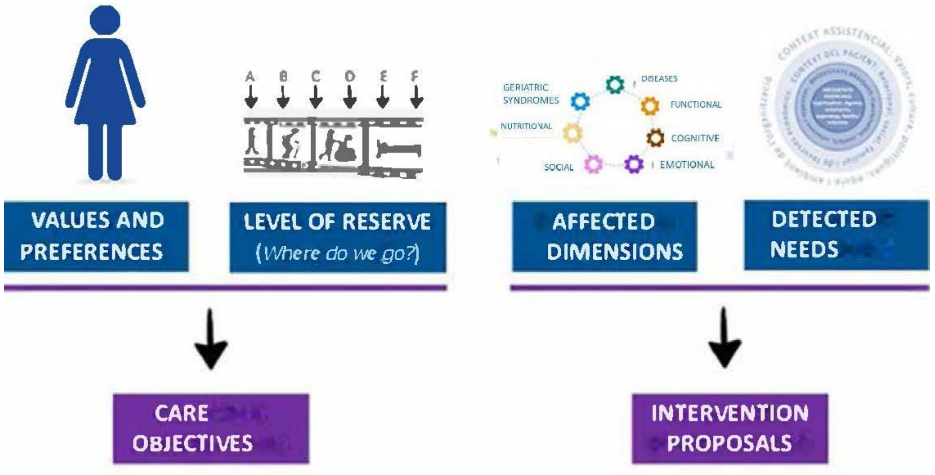

Figure 2. Care objectives and intervention proposals as key elements for developing a person-centred care plan. *Source: own elaboration*.

##### Scales and assessment instruments for different dimensions

Although CGA should not be understood as a simple accumulation of scales, the evaluation of each of the dimensions is essential. Some of the most commonly used scales are mentioned below:

| DIMENSION | | PROPOSED ASSESSMENT INSTRUMENT |
|---|---|---|
| **FUNCTIONAL** | **Instrumental** activities of Daily Living Scale (IADL) | Brody and Lawton scale |
| | **Basic** activities of daily living (ADL) | Barthel index |
| **MENTAL** | Cognitive | Mini-Mental State Examination (MMSE) Pfeiffer Test<br>Mini-Cog |
| | Emotional | Yesavage Geriatric Depression Scale |
| **SOCIAL** | Social risk | Gijón Scale<br>Self-Sufficiency Matrix |
| | Caregivers' burden | Zarit Test |
| **NUTRITIONAL** | | Mini-Nutritional Assessment (MNA) |
| **GERIATRIC SYNDROMES and SYMPTOMS** | Delirium | Confusional Assessment Method (CAM) |
| | Pressure ulcers | Braden scale<br>Norton Scale |
| | Dysphagia | Volume-viscosity test |
| | Balance and gait | Tinetti scale |
| | Symptoms | Edmonton Symptom Assessment System (ESAS) |
| **QUALITY OF LIFE** | | Euroqol 5D |

Assessment of frailty:

- To distinguish frail from non-frail individuals, tools such as the Gérontopôle Frailty Screening Tool (GFST) or the FRAIL scale, or functional tests such as the Short Physical Performance Battery (SPPB) or walking speed.
- For assessing the degree of reserve/fragility of the person, both clinical frailty scales and fragility indices (for example, the index, of which there is an Excel calculator) may be useful.

In order to be able to apply the CGA properly, it is recommended that health professionals in HaH units receive training and/or that geriatric specialists be incorporated into existing HaH units.

In Catalonia, professionals from different areas of the healthcare and social system have been working on the consensus for a rapid multidimensional/geriatric assessment tool (CGA-express; VIG-express in Catalan), which will allow other health professionals to have a fast and shared multidimensional approach.

This consensus has been promoted by the Department of Health of the Generalitat de Catalunya and endorsed by the Integrated Social and Health Care Plan (PAISS) and will be published soon.

In summary, elderly patients, those with frailty, multimorbidity and in advanced disease situations (PCC and MACA) must be treated in HaH following the geriatric model, using the same methodology developed and validated in the acute and intermediate geriatric hospitalisation units based on interdisciplinary work, CGA as a working tool and the consensus of complex interventions.

#### Appendix 8. HaH indicator sheets

**CODE: 01**

##### IDENTIFICATION

**INDICATOR NAME: Patients with hospital at home (HaH).**

##### DESCRIPTION

**DEFINITION:** Number of patients identified by NIA (personal identification number) with HaH per centre.

**OBJECTIVE:** To know the number of people who have been treated in the HaH unit in each centre.

**CALCULATION FORMULA:** Number of HaH patients counted from the NIA.

**UNIT OF MEASURE:** Number of patients.

**INTERPRETATION AND LIMITATIONS:** To know the people who have been treated in the HaH unit of each centre in order to monitor and evaluate their evolution.

**PERIODICITY:** Annual.

##### SOURCES OF INFORMATION

**DATA SOURCE:** CMBD - Acute hospitals.

**RESPONSIBLE STRUCTURE:** CatSalut.

##### ANALYSIS DIMENSIONS

**GENERIC ANALYSIS:** Public hospitals in Catalonia.

**CODE: 02**

#### IDENTIFICATION

**INDICATOR NAME: Number of episodes (contacts) with hospital at home (HaH).**

#### DESCRIPTION

**DEFINITION:** Number of hospital contacts, that is, of records with HaH.

**OBJECTIVE:** To know the number of contacts that have originated patients with HaH in each hospital.

**CALCULATION FORMULA:** Number of records with HaH per centre.

**UNIT OF MEASURE:** Number of contacts.

**INTERPRETATION AND LIMITATIONS:** To know the number of records that have been completed with HaH. To differentiate it from the number of patients, it expands the information of how many times the same patient called upon HaH. It also allows us to monitor and evaluate their evolution.

**PERIODICITY:** Annual.

#### INFORMATION SOURCES

**DATA SOURCE:** CMBD - Acute hospitals.

**RESPONSIBLE STRUCTURE:** CatSalut.

#### ANALYSIS DIMENSIONS

**GENERIC ANALYSIS:** Public hospitals in Catalonia.

**CODE: 03**

#### IDENTIFICATION

**INDICATOR NAME: HaH admission ratio per patient.**

#### DESCRIPTION

**DEFINITION:** HaH admission ratio per patient, regardless of diagnosis, in each of the centres and globally for Catalonia.

**OBJECTIVE:** To have information on the HaH admission ratio per patient of each hospital centre and to be able to compare it with that of Catalonia.

**CALCULATION FORMULA:**

**Numerator:** Number of records with HaH per centre.

**Denominator:** Number of patients with HaH per centre.

**UNIT OF MEASURE:** Ratio, numerical decimal (one decimal).

**INTERPRETATION AND LIMITATIONS:** Knowing the HaH admission ratio per patient per centre gives us global information on readmissions to HaH. It is easier to monitor and evaluate its evolution. It is used to detect the frequency of patients in this type of admission.

**Limitations**: It does not identify readmissions for the same pathology, it aims for a global analysis of frequency by HaH and patient.

**PERIODICITY:** Annual.

#### INFORMATION SOURCES

**DATA SOURCE:** CMBD - Acute hospitals.

**RESPONSIBLE STRUCTURE:** CatSalut.

#### ANALYSIS DIMENSIONS

**GENERIC ANALYSIS:** Public hospitals in Catalonia.

**CODE: 04**

#### IDENTIFICATION

**INDICATOR NAME: HaH modalities: admission avoidance (AA) and early supported discharge (ESD).**

#### DESCRIPTION

**DEFINITION:** Number of contacts, i.e., records with HaH for each modality and centre.

**Admission avoidance**: When the person is admitted directly to HaH without prior admission to the same hospital for the same reason. The "origin" variable is emergencies, day hospital, outpatient visits, PADES or APIC are considered 'discharge' with admission avoidance.

**Early supported discharge**: There is a previous conventional hospitalisation contact.

**OBJECTIVE:** To know how the use of HaH is distributed for each modality and centre.

**CALCULATION FORMULA:** Number and percentage of records with HaH of each modality and for each centre.

**UNIT OF MEASURE:** Number and percentage of contacts for each of the HaH modalities.

**INTERPRETATION AND LIMITATIONS:** Knowing the number of records of HaH by modality, describes the actions of each hospital. It gives us information on the percentage variability of the modalities according to the behaviour of the centre and allows us to study the existing heterogeneity.

**PERIODICITY:** Annual.

#### INFORMATION SOURCES

**DATA SOURCE:** CMBD - Acute hospitals.

**RESPONSIBLE STRUCTURE:** CatSalut.

#### ANALYSIS DIMENSIONS

**GENERIC ANALYSIS:** Public hospitals in Catalonia.

**CODE: 05**

#### IDENTIFICATION

**INDICATOR NAME: Patient by age and sex in both HaH modalities.**

#### DESCRIPTION

**DEFINITION:** Number of patients with HaH according to sex and age group, in each hospital.

**Sex:** Male and female.

**Age group**: < 15 years, 15-44, 45-64, 65-74, 75-84 and > 84 years.

**OBJECTIVE:** To know the profile of patients admitted to HaH according to age and sex for each of the modalities and hospitals.

**CALCULATION FORMULA:** Number and percentage of patients by sex and age group with HaH for each modality and hospital.

**Numerator:** Number of patients in HaH according to modality.

**Denominator:** Number of patients with HaH. It should be specified in each age group and by sex.

**UNIT OF MEASURE:** Number and percentage of patients.

**INTERPRETATION AND LIMITATIONS:** Knowing the characteristics of patients who use HaH by age groups guides us to reinforce resources in a group, if necessary.

**PERIODICITY:** Annual.

#### INFORMATION SOURCES

**DATA SOURCE:** CMBD - Acute hospitals.

**RESPONSIBLE STRUCTURE:** CatSalut.

#### ANALYSIS DIMENSIONS

**GENERIC ANALYSIS:** Public hospitals in Catalonia.

**CODE: 06**

#### IDENTIFICATION

**INDICATOR NAME: Destination at discharge.**

#### DESCRIPTION

**DEFINITION:** Define the destination at discharge of contacts admitted to HaH according to modality (admission avoidance *vs.*early supported discharge).

**Type of destination at discharge:**

Home or residence
Discharge with continuity of care in another centre
Discharge with continuity of care in the same centre
Voluntary discharge
Evasion or administrative discharge
Exitus

**OBJECTIVE:** To have information on how the HaH episode is resolved in each modality.

**CALCULATION FORMULA:** Number and percentage of the destination at discharge of contacts with HaH for each modality and hospital.

**UNIT OF MEASURE:** Number and percentage of contacts.

**INTERPRETATION AND LIMITATIONS:** Knowing the destination at discharge of contacts helps to know the behaviour of patients admitted to HaH.

**Limitations**: The information about the evolution of discharged patients is lost, unless a specific analysis is carried out later, for example, we know hospital mortality, but not if the patient has died two days later after leaving the centre.

**PERIODICITY:** Annual.

#### INFORMATION SOURCES

**DATA SOURCE:** CMBD - Acute hospitals.

**RESPONSIBLE STRUCTURE:** CatSalut.

#### ANALYSIS DIMENSIONS

**GENERIC ANALYSIS:** Public hospitals in Catalonia.

**CODE: 07**

#### IDENTIFICATION

**INDICATOR NAME: Main diagnosis of contacts with HaH.**

#### DESCRIPTION

**DEFINITION:** Describe the main diagnosis according to the ADP grouping of contacts admitted by HaH according to modality (avoidance *vs*. early discharge) and centre.

**OBJECTIVE:** To know the diagnostic activity of HaH admissions to characterize the patients who have used each hospitalisation model.

**CALCULATION FORMULA:** Number and percentage of the main diagnosis of HaH contacts by modality and hospital.

**UNIT OF MEASURE:** Number and percentage of main diagnoses.

**INTERPRETATION AND LIMITATIONS:** HaH admissions diagnoses help to characterize patients who have used each hospitalisation model and to know the most frequent ones.

**Limitations**: It gives us information on the main diagnosis; secondary ones are missing to complement the characterization of patients. It is proposed to do so in a second phase.

**PERIODICITY:** Annual.

#### INFORMATION SOURCES

**DATA SOURCE:** CMBD - Acute hospitals.

**RESPONSIBLE STRUCTURE:** CatSalut.

#### ANALYSIS DIMENSIONS

**GENERIC ANALYSIS:** Public hospitals in Catalonia.

**CODE: 08**

#### IDENTIFICATION

**INDICATOR NAME: Main diagnosis of contacts with HaH by age group.**

#### DESCRIPTION

**DEFINITION:** Describe the main diagnosis according to the ADP grouping (“Agrupació per a diagnòstic i procediments” or "Diagnostic and Procedure Classification) of contacts admitted by HaH by age groups and centre.

**OBJECTIVE:** To have information on the diagnosis of contacts to identify the most common causes of use of HaH by age.

**CALCULATION FORMULA:** Number and percentage of the main diagnosis of HaH contacts by age group and hospital.

**UNIT OF MEASURE:** Name and percentage of the main diagnoses.

**INTERPRETATION AND LIMITATIONS:** Knowing the diagnostic activity of HaH admissions helps to characterize patients according to the age groups described in the patient profile sheet.

**Limitations**: It gives us information on the main diagnosis; secondary diagnoses are missing to complement the characterization of patients. It is proposed to include them in a second phase.

**PERIODICITY:** Annual.

#### INFORMATION SOURCES

**DATA SOURCE:** CMBD - Acute hospitals.

**RESPONSIBLE STRUCTURE:** CatSalut.

#### ANALYSIS DIMENSIONS

**GENERIC ANALYSIS:** Public hospitals in Catalonia.

**CODE: 09**

#### IDENTIFICATION

**INDICATOR NAME: Procedures of contacts with HaH.**

#### DESCRIPTION

**DEFINITION:** Describe the main procedure according to the APP grouping of contacts admitted by HaH according to modality (avoidance *vs*. early discharge) and centre.

**OBJECTIVE:** To know the procedure used by contacts in order to identify the most frequent types of procedures applied to patients admitted to HaH for each modality and centre.

**CALCULATION FORMULA:** Number and percentage of the main procedure of HaH contacts by modality and hospital.

**UNIT OF MEASURE:** Number and percentage of the main procedures.

**INTERPRETATION AND LIMITATIONS:** The type of procedure for HaH admissions helps to characterize patients who used each hospitalisation model and can facilitate the adequacy and adaptation of the procedure at home.

**Limitations**: It gives us information on the main procedure; the secondary ones are missing to complement the characterization of patients. It is proposed to include them in a second phase.

**PERIODICITY:** Annual.

#### INFORMATION SOURCES

**DATA SOURCE:** CMBD - Acute hospitals.

**RESPONSIBLE STRUCTURE:** CatSalut.

#### ANALYSIS DIMENSIONS

**GENERIC ANALYSIS:** Public hospitals in Catalonia.

**CODE: 10**

#### IDENTIFICATION

**INDICATOR NAME: Procedures for contacts with HaH according to age group.**

#### DESCRIPTION

**DEFINITION:** Describe the main procedure according to the ADP grouping of contacts admitted to HaH by age groups and centre.

**OBJECTIVE:** To have information on the procedure used in contacts to identify the most frequent types of procedures applied to patients admitted to HaH by each age group.

**CALCULATION FORMULA:** Number and percentage of the main procedure of contact with HaH by age groups and hospital.

**UNIT OF MEASURE:** Number and percentage of the main procedures of the contacts.

**INTERPRETATION AND LIMITATIONS:** The type of procedure for admissions to HaH helps to characterize patients who have used each type of hospitalisation and can facilitate the adequacy and adaptation of the procedure at home.

**Limitations**: It gives us information on the main procedure; secondary ones are missing to complement the characterization of patients. It is proposed to include them in a second phase.

**PERIODICITY:** Annual.

#### INFORMATION SOURCES

**DATA SOURCE:** CMBD - Acute hospitals.

**RESPONSIBLE STRUCTURE :** CatSalut.

#### ANALYSIS DIMENSIONS

**GENERIC ANALYSIS:** Public hospitals in Catalonia.

**CODE: 11**

#### IDENTIFICATION

**INDICATOR NAME: HaH discharges versus conventional discharges in hospitals.**

#### DESCRIPTION

**DEFINITION:** Describe the number and percentage of HaH discharges in relation to discharges by conventional hospitalisation by hospital.

**OBJECTIVE:** To know the number of HaH discharges at each centre in order to follow up over the years and, at the same time, to be able to analyse the variability between centres.

**CALCULATION FORMULA:** It should be done globally (Catalonia) and per hospital.

**Numerator:** Number of contacts with HaH per centre.

**Denominator:** Number of registrations with conventional hospitalisation per centre.

**UNIT OF MEASURE:** Number and percentage.

**INTERPRETATION AND LIMITATIONS:** The percentage of HaH in relation to conventional hospitalisation is important to assess the application of this alternative hospitalisation that provides a high quality of life for the patient. It helps us to compare with other countries and also among our own centres.

**Limitations**: It must be taken into account, when comparing centres, that this type of admission requires some items, such as isochrones and family/caregiver conditions, among others, which are difficult to achieve in hospitals located in geographically very dispersed territories with a very old population.

**PERIODICITY:** Annual.

#### INFORMATION SOURCES

**DATA SOURCE:** CMBD - Acute hospitals.

**RESPONSIBLE STRUCTURE:** CatSalut.

#### ANALYSIS DIMENSIONS

**GENERIC ANALYSIS:** Public hospitals in Catalonia.

**CODE: 12**

#### IDENTIFICATION

**INDICATOR NAME: HaH discharges *vs*. conventional discharges in territories.**

#### DESCRIPTION

**DEFINITION:** Describe the number and percentage of HaH discharges in relation to discharges by conventional hospitalisation by territory.

**OBJECTIVE:** To know the number of HaH registrations that are made in the territory to be able to monitor it over time and, at the same time, to be able to analyse the variability between territories.

**CALCULATION FORMULA:** It should be done globally (Catalonia) and by territories.

**Numerator:** Number of HaH contacts by territory.

**Denominator:** Number of registrations with conventional hospitalisation by territories.

**UNIT OF MEASURE:** Number and percentage.

**INTERPRETATION AND LIMITATIONS:** The percentage of HaH in relation to conventional hospitalisation is important to assess the application of this hospitalisation alternative that provides a high quality of life for patients. It helps us to compare with other countries and also among territories.

**Limitations**: It must be taken into account, when comparing territories, that this type of admission is conditioned by specific isochrones and family/caregiver conditions, among others, which are difficult to achieve in hospitals located in geographically dispersed territories with a very old population.

**PERIODICITY:** Annual.

#### INFORMATION SOURCES

**DATA SOURCE:** CMBD - Acute hospitals.

**RESPONSIBLE STRUCTURE:** CatSalut.

#### ANALYSIS DIMENSIONS

**GENERIC ANALYSIS:** Public hospitals in Catalonia.

**CODE: 13**

#### IDENTIFICATION

**INDICATOR NAME: Complexity of HaH contacts by centre and age group.**

#### DESCRIPTION

**DEFINITION:** Describe the complexity of the contacts admitted by HAH by centre, according to the diagnosis-related groups (DRG).

**OBJECTIVE:** To assess the degree of complexity of the contacts that have been admitted to HAH, by centres.

**CALCULATION FORMULA**: Number and percentage of the first 10 most frequent DRGs, includes all HaH contacts. It is specified by hospital.

**UNIT OF MEASURE:** Number and percentage.

**INTERPRETATION AND LIMITATIONS:** DRG is a way of knowing the complexity of patients, since it is a variable calculated from a set of items such as age, comorbidity, sex, among others. It brings us closer and helps to characterize HaH admissions with severity criteria.

**Limitations**: It is a synthetic variable that brings us closer to complexity, without refining it in all cases.

**PERIODICITY:** Annual.

#### INFORMATION SOURCES

**DATA SOURCE:** CMBD - Acute hospitals.

**RESPONSIBLE STRUCTURE:** CatSalut.

#### ANALYSIS DIMENSIONS

**GENERIC ANALYSIS:** Public hospitals in Catalonia.

**CODE: 14**

#### IDENTIFICATION

**INDICATOR NAME: Length of stay of contacts with HaH according to centre.**

#### DESCRIPTION

**DEFINITION:** Describe the length of stay of the contacts admitted for both modalities of HaH in different centres.

**OBJECTIVE:** To assess whether the length of stay with HaH is similar to the stay in conventional hospitalisation and see the heterogeneity between centres.

**CALCULATION FORMULA:** Mean, median, mode, variance, standard deviation (SD), range and 25th, 50th and 75th percentiles of the stay calculated in days.

**UNIT OF MEASURE:** Whole number or decimal (two decimal places), if applicable.

**INTERPRETATION AND LIMITATIONS:** The length of stay gives us indirect information on the behaviour and effectiveness of the centres depending on the patient's morbidity. We will be able to assess the centres in relation to those of the rest of Catalonia.

**Limitations**: In order to make more precise comparisons, it is necessary to do so for the same pathologies, at least adjusting by complexity, which is expected to be done at a later stage.

**PERIODICITY:** Annual.

#### INFORMATION SOURCES

**DATA SOURCE:** CMBD - Acute hospitals.

**RESPONSIBLE STRUCTURE:** CatSalut.

#### ANALYSIS DIMENSIONS

**GENERIC ANALYSIS:** Public hospitals in Catalonia.

**CODE: 15**

#### IDENTIFICATION

**INDICATOR NAME: Readmissions of HaH contacts within 30 days.**

#### DESCRIPTION

**DEFINITION:** Describe the number of readmissions in 30 days of HaH contacts in each centre.

**OBJECTIVE:** To assess the number of readmissions in 30 days as an indirect indicator of the effectiveness of HaH considering the limitations, and also to describe if there is a lot of variability between centres.

**CALCULATION FORMULA:** Number of patients who readmit to the same centre in less than 30 days from the date of discharge.

**UNIT OF MEASUREMENT:** Number and percentage of patients.

**INTERPRETATION AND LIMITATIONS:** Assessing readmissions is an indicator of the behaviour and actions of the centre and shows us the differences with the conventional hospitalisation.

**Limitations**: In order to make more appropriate comparisons, it would be necessary to do so for the same pathologies, at least adjusting by complexity, which is expected to be done at a later stage.

**PERIODICITY:** Annual.

#### INFORMATION SOURCES

**DATA SOURCE:** CMBD - Acute hospitals.

**RESPONSIBLE STRUCTURE:** CatSalut.

#### ANALYSIS DIMENSIONS

**GENERIC ANALYSIS:** Public hospitals in Catalonia.

#### Annex 9. Prescription of respiratory therapies

Definition of the resources necessary for an HaH supported by home respiratory therapies.

##### Respiratory therapies during HaH

During the home hospitalisation, the hospital is in charge of the integral treatment of the patient, their pharmacological treatment and, where appropriate, their home oxygen treatment, nebulized therapy, with the possibility of performing arterial blood gasometry at home.

**The service lines included in the respiratory therapies of HaH are:**

| |
|---|
| OB - Oxygen therapy with gas cylinder |
| OC - Oxygen therapy with static concentrator from 0 to 4 lpm |
| OC - Oxygen therapy with static concentrator from 0 to 5 lpm |
| OC - Oxygen therapy with static concentrator from 1 to 10 lpm |
| OL - Oxygen therapy with liquid oxygen |
| AC - Nebulization with conventional jet nebulizers |
| AF - Nebulization with jet nebulizers with high flow jet nebulizers |
| AE - Nebulization with electronic nebulizers with vibrating mesh |
| AP - Nebulization with specific nebulizers according to prescription |
| GM - Portable arterial blood gasometry |

The hospital must formalize the contract for the supply of therapies with one of the supply companies that comply with the gas handling regulations and have authorization for manufacture, storage and marketing by the Agency of Medicines and Health Products (Agència del Medicament i Productes Sanitaris, AEMPS). The hospital's contract with the company must include the supply of specific equipment, accessories, and specific consumables for each therapy, as well as the provision of services at the patient's home, installation and maintenance of the equipment. Training on the use of the equipment must be coordinated with the HaH teams of each territory. The profile of this type of patient, in HaH, is the person with acute respiratory pathology who requires hospitalisation if there is no HaH service available.

The prescription circuit must be established by each hospital in the context of the patient's admission to their home. The prescription of home respiratory therapies, when the patient is admitted to HaH, must be made by the medical staff of the same hospital.

The company must ensure the installation of the equipment before the patients arrive at their home, and, if not possible, always within the first 2 hours from the request.

Under no circumstances can a patient with home admission be prescribed oxygen using the home respiratory therapies prescription circuit of the application portal. This circuit is exclusively aimed at situations of chronic long-term pathology. In case the patient has a previous chronic respiratory therapy prescription, the service can be maintained, but if it needs to be increased or expanded with some additional therapy, it must be guaranteed by the same hospital during admission to HaH.

#### 10. Bibliographical references

Alepuz L, Antón F, Arias J, Espallargues M, Estrada MD, Estrada O, Hermida L, Fernández M, Massa B, Mirón M, Murja A, Muñoz L, Ponce MA, Rio M, Torres A. Hospitalització domiciliària. Barcelona: Agència de Qualitat i Avaluació Sanitàries de Catalunya. Departament de Salut. Generalitat de Catalunya; 2018.

Amblàs-Novellas J, Espaulella J, Rexach L et al. Frailty, severity, progression and shared decision-making: A pragmatic framework for the challenge of clinical complexity at the end of life. Eur Geriatr Med 2015;6(2):189–194.

Amblàs-Novellas, Jordi; Contel, Joan Carles; Kukielka, Deborah; Rico, Lalo; Barbeta, Conxita; Santaeugènia, Sebastià; en nom del grup VIG-express. VIG-express: consenso de un sistema de valoración multidimensional/geriátrica rápida en Cataluña. In press.

Busquet X, Esteban M, Jiménez EM, Tura M, Bosch O, Moragas A et al. Debate: criterios de complejidad utilizados en cuidados paliativos. El hexágono de la complejidad (HexCom®2018). In:.In: Limón E MA (coord.), editor. Cronicidad avanzada. Limón E, Meléndez A (coord.), Madrid: SECPAL; 2018. p. 35–40. Available at: http://www.secpal.com/Documentos/Blog/Monografia%20Cronicidad.pdf

Busquet-Duran X. L'hexàgon de la complexitat (HexCom). Intercanvis/Intercambios Psicoanàlisi. 2017;38:86–106. Available at a: http://www.raco.cat/index.php/Intercanvis/article/view/330764

Carme Hernández, Albert Alonso, Judith Garcia-Aymerich et al. Integrated care services: lessons learned from the deployment of the NEXES project. Int J Integr Care. 2015 Jan-Mar; 15.

Carme Hernández, Jesus Aibar, Nuria Seijas et al. Implementation of Home Hospitalisation and Early Discharge as an Integrated Care Service: A Ten Years Pragmatic Assessment. Int J Integr Care. 2018 Apr-Jun; 18(2): 12.

Catàleg d'indicadors: Avedis Donabedian. Milbank Q. 2005 Dec; 83(4): 691–729.

Coleman EA, Parry C, Chalmers S et al. The care transitions intervention: results of a randomized controlled trial. Arch Intern Med 2006; 166(17):1822-1828.

Conley J, O'Brien CW, Leff BA, Bolen S, Zulman D. Alternative Strategies to Inpatient Hospitalisation for Acute Medical Conditions. A Systematic Review. JAMA Intern Med. 2016;176(11):1693-1702. doi:10.1001/jamainternmed.2016.5974.

Cuxart A, Estrada O. Hospitalización a domicilio: una oportunidad para el cambio. Med Clin (Barc) 2011; 138: 355-60.

Esteban-Pérez M, Fernández-Ballart J, Boira-Senlí R, Martínez- Serrano T, Nadal-Ventura S, Castells-Trilla G. Concordancia entre la complejidad observada desde diferentes niveles asistenciales en pacientes crónicos complejos, con enfermedad avanzada o al final de la vida mediante un modelo de abordaje de la complejidad. Med Paliat. 2018;25. Available at: http://www.elsevier.es/es-revista-medicina-paliativa-337-pdf-S1134248X18300028-S200

Esteban-Pérez M, Grau IC, Castells-Trilla G, Bullich-Marín I, Busquet-Duran X,

Aranzana-Martínez A, Basora Torradeflot M, Picaza Vila J, Tuca Rodríguez A, Valverde Vilabella E. Complejidad asistencial en la atención al final de la vida: criterios y niveles de intervención en atención comunitaria de salud. Med Paliat. 2015;22:69–80.

Estrada O, Massa B, Ponce MA, Mirón M, Torres A, Mujal A et al. Proyecto HaD 2020: una propuesta para consolidar la hospitalización a domicilio en España. Hosp Domic. 2017;1(2):93-117.

González-Ramallo VJ, Valdivieso B, Ruiz V. Hospitalización a domicilio. Med Clin (Barc). 2002;118:659–64.

Grup de Treball d'Hospitalització Domiciliària i Geriatria Comunitària de la Societat Catalana de Geriatria i Gerontologia. Model d'hospitalització domiciliària integral geriàtrica per a l'atenció de persones d'edat avançada amb fragilitat i complexitat.Gener 2020. Available at: http://scgig.cat/grup-hospitalitzacio

Jencks SF, Williams MV, Coleman EA. Rehospitalisations among patients in the Medicare fee-for-service program. N Engl J Med 2009; 360(14):1418-1428.

Leff B, Burton L, Mader SL et al. Hospital at home: feasibility and outcomes of a program to provide hospital-level care at home for acutely ill older patients. Ann Intern Med 2005; 143(11):798-808.

Leff B. Defining and disseminating the hospital-at-home model. CMAJ 2009; 180(2):156-157.

Lu CY, Roughead E. Determinants of patient-reported medication errors: a comparison among seven countries. Int J Clin Pract 2011; 65(7):733-740.

Mas MÀ, Santaeugènia S. Hospital-at-home in older patients: a scoping review on opportunities of developing comprehensive geriatric assessment-based services. Rev Esp Geriatr Gerontol. 2015;50(1):26-34.

Mas MÀ, Santaeugènia S. Hospitalización domiciliaria en el paciente anciano: revisión de la evidencia y oportunidades de la geriatría. Rev Esp Geriatr Gerontol 2015;50(1):26-34.

Mas MÀ, Santaeugènia SJ, Tarazona-Santabalbina FJ, Gámez S, Inzitari M. Effectiveness of a Hospital-at-Home Integrated Care Program as Alternative Resource for Medical Crises Care in Older Adults With Complex Chronic Conditions. J Am Med Dir Assoc 2018;19(10):860-863.

Moore C, Wisnivesky J, Williams S et al. Medical errors related to discontinuity of care from an inpatient to an outpatient setting. J Gen Intern Med 2003; 18(8):646-651.

Naylor MD. Transitional care for older adults: a cost-effective model. LDI Issue Brief 2004; 9(6):1-4.

Pilotto A, Cella A, Pilotto A, Daragjati J, Veronese N, Musacchio C et al. Three Decades of Comprehensive Geriatric Assessment: Evidence Coming From Different Healthcare Settings and Specific Clinical Conditions. J Am Med Dir Assoc 2017;18(2):192.e1-192.e11.

Rennke S, Nguyen OK, Shoeb MH et al. Hospital-initiated transitional care interventions as a patient safety strategy: a systematic review. Ann Intern Med 2013; 158(5 Pt 2):433-440.

Salvador Comino, Rosa; Garrido Torres, Nathalia; Perea Cejudo, Inmaculada; Martin Rosello, Maria Luisa; Regife Garcia, Victor; Fernandez Lopez A. El valor del instrumento diagnóstico de la complejidad en cuidados paliativos para identificar la complejidad en pacientes tributarios de cuidados paliativos. Med Paliat. 2017;24:196–203.

Shepperd S, Doll H, Angus RM et al. Hospital at home admission avoidance (Review). The Cochrane Library 2011, Issue 8, 2011.

Stuck AE, Iliffe S. Comprehensive geriatric assessment for older adults. BMJ 2011;343:d6799.

**References on the legal framework related to the transfusion of blood components**

Comité Científico para la Seguridad Transfusional (CCST). Transfusión extrahospitalaria. Recomendaciones. Ministerio de Sanidad, Servicios Sociales e Igualdad, 2016. [Scientific Committee for Transfusion Safety (CCST). Out-of-hospital transfusion. Recommendations. Ministry of Health, Social Services and Equality, 2016.]

Dispositiu transversal d'hospitalització a domicili i Servei d'Hemoteràpia i Hemostàsia, Transfusió de components sanguinis, Hospital Clínic de Barcelona, 2016. [Cross-disciplinary home hospitalisation device and Haemotherapy and Haemostasis Service, Blood Component Transfusion, Hospital Clínic de Barcelona, 2016.]

Guía sobre la transfusión de componentes sanguíneos y derivados plasmáticos de la SETS. 4ª edición, 2010. [Guide on the transfusion of blood components and plasma derivatives of SETS. 4th edition, 2010.]

Real decreto 1088/2005, de 16 de septiembre (Ref.2005/15514), por el que se establecen los requisitos técnicos y condiciones mínimas de la hemodonación y de los centros y los servicios de transfusión. BOE número 225, de 20 de septiembre de 2005. [Royal decree 1088/2005, of 16 September (Ref.2005/15514), by which establish the technical requirements and minimum conditions of the hemodonation and of the centres and the services of transfusión. BOE number 225, of 20 September 2005.]

Transfusión extrahospitalaria. Acuerdos 17/12/15 y 18/02/16. Comité científico sobre seguridad transfusional. Ministerio de Sanidad, Servicios Sociales e Igualdad. [Out-of-hospital transfusion. Agreements 17/12/15 and 18/02/16. Scientific Committee on Transfusion Safety. Ministry of Health, Social Services and Equality.]

Trasfusión domiciliaria de hemoderivados. Recomandaciones clínicas y procedimientos en Hospitalización Domiciliaria. Sanroma P, Sanpedro I, Gonzalez C Baños MT (Ed) Tomo 2 Cap:3,1293-1316, IFIMAV, 2012.

Villegas Bruguera, EB i Torres Corts, AM. Transfusió a domicili (HADO) en transfusió de components sanguinis. Protocol Clínic “Guia d’hemoteràpia”, Hospital Dos de Maig, Consorci Sanitari Integral, 2013. [Home transfusion of blood products. Clinical recommendations and procedures in Home Hospitalisation. Sanroma P, Sanpedro I, Gonzalez C Baños MT (Ed) Tomo 2 Cap:3,1293-1316, IFIMAV, 2012.]

Villegas Bruguera, EB and Torres Corts, AM. Home transfusion (HADO) in blood component transfusion. Clinical Protocol "Haemotherapy Guide", Hospital Dos de Maig, Consorci Sanitari Integral, 2013.
